# Duration of protection after vaccination against yellow fever - systematic review and meta-analysis

**DOI:** 10.1101/2022.06.21.22276699

**Authors:** Kerstin Kling, Cristina Domingo, Christian Bogdan, Steven Duffy, Thomas Harder, Jeremy Howick, Jos Kleijnen, Kevin McDermott, Ole Wichmann, Annelies Wilder-Smith, Robert Wolff

## Abstract

**Background:** The duration of protection after a single dose of yellow fever vaccine is a matter of debate. To summarize the current knowledge, we performed a systematic literature review and meta-analysis.

**Methods:** Studies on the duration of protection after 1 and ≥2 vaccine doses were reviewed. Data were stratified by time since vaccination. In our meta-analysis, we used random-effects models.

**Results:** We identified 36 studies from 20 countries, comprising over 17,000 participants aged 6 months to 85 years. Among healthy adults and children, pooled seroprotection rates after single vaccination dose were close to 100% by 3 months and remained high in adults for 5 to 10 years. In children vaccinated before age 2, the seroprotection rate was 52% within 5 years after primary vaccination. For immunodeficient persons, data indicate relevant waning.

**Conclusions:** The extent of waning of seroprotection after yellow fever vaccination depends on age at vaccination and immune status.

**Summary:** Systematic Review and meta-analysis of existing studies indicates a relevant waning of seroprotection after a single dose of yellow fever vaccination of different groups including healthy adults, children when vaccinated before the age of 2 years and immunodeficient persons.

## Introduction

Yellow fever (YF) is a vector-borne disease transmitted by mosquitoes of the *Aedes* and *Haemagogus* species. In 2020, 40 countries in Africa and South America were classified as endemic by the World Health Organization (WHO) [1]. The case fatality rate with the severe form of YF can reach 60% or more in persons with underlying diseases (such as diabetes mellitus) [2, 3]. Each year, approximately 200,000 YF cases and 30,000 YF deaths occur worldwide [4]. As no licensed drugs are available to treat YF, reduction of disease burden is exclusively accomplished through vaccination and vector control. Despite effective YF vaccines being available since the 1930s, outbreaks continue to occur and the disease has spread into new areas during recent decades [5-7]. To date, it is unknown to what extent a lack of seroconversion (primary vaccine failure) and waning immunity (secondary vaccine failure) influence the individual risk of YF.

For many years, YF vaccine booster doses were recommended every 10 years for those at risk of exposure, including people living in endemic countries and travellers. In 2013, WHO decided that a single dose of the YF vaccine is usually sufficient to confer lifelong protection against YF, except for certain sub-populations such as persons with immunodeficiencies (e.g. HIV). Accordingly, International Health Regulations (IHR) were adapted in 2016 concerning validity of vaccination certificates. Since then, the sufficiency of a single dose for life-long protection has been questioned for various reasons [8-12]. To provide an up-to-date overview of the currently available data and knowledge, we performed a systematic review (SR) and meta-analysis on the duration of protection after vaccination against YF.

## Methods

### Inclusion and exclusion criteria

We performed a SR in accordance with the methods recommended by Cochrane (formerly The Cochrane Collaboration) and the Centre for Reviews and Dissemination [13, 14]. Studies were selected for inclusion based on the following criteria:

- Population: People living in areas where YF is endemic (list of countries as defined by WHO) as well as travellers from non-endemic areas. Subgroups: children, adults (≥18 years), older adults (≥60 years), those with any form of immunodeficiency, pregnant women, and persons who have received a vaccine against another flavivirus
- Intervention: Any full single dose of a licenced YF vaccine
- Comparators: Placebo, no vaccine, other vaccine, fractional dose or booster dose(s) of YF vaccine alone or with another vaccine against another related viral disease, e.g. dengue fever, Japanese encephalitis
- Outcomes: Proportion (%) of individuals with YF; proportion (%) of individuals with death due to YF; seropositivity rates – e.g. proportion (%) of individuals who are seropositive for neutralising antibodies against YF

We included randomised controlled trials (RCT), non-randomized (observational) studies with control groups and prospective single-armed observational studies with ≥50 participants (non-RCT). Retrospective single arm studies, prospective single arm studies with <50 participants and case reports/series were excluded.

### Literature search

We performed a systematic literature search in 15 databases (date of last search: 12.11.2021).

Search strategies combined relevant search terms comprising indexed keywords (e.g., Medical Subject Headings [MeSH]) and free text terms appearing in the titles and/or abstracts of database records. Search terms were identified through discussion, by scanning background literature and ‘key articles’ already known to the project team, and by browsing database thesauri. Searches were not limited by language, geographic location, publication status or date of publication. The Embase search strategy was peer reviewed by a second information specialist. After removal of duplicates, 4,800 records remained for further screening based on titles and abstracts. Full details of all search strategies are provided in Supplementary Data (Supplementary 1 - Search strategies: Table 1 and 2).

**Table 1:** Characteristics of included studies.

| Study | Country<br>(endemic region<br>Y/N) | Subjects | n | Vaccine-specification | Specific Assay | Serology after YF<br>vaccination up to |
| --- | --- | --- | --- | --- | --- | --- |
| **RCT** |  |  |  |  |  |  |
| Asante et al., 2020 [23] | Ghana (Y) | Children | 709 | 17D (Stamaril) | PRNT50 | 1 month |
| Belmusto-Worm et al., 2005 [24] | Peru (Y) | Children | 1107 | 17D (Arlvax, YF-VAX) | LNI | 1 month |
| Camacho et al., 2004 [21] | Brazil (N) | Adults | 1087 | 17D (Amaril), 17DD | PRNT50 | 1 month |
| Campi-Azevedo et al., 2014 [25] | Brazil (N) | Adults | 590 | 17DD | PRNT50 | 1 year |
| Chan et al., 2016 [26] | Singapur (N) | Adults | 70 | 17D (Stamaril) | PRNT50 | 1 month |
| Collaborative Group, 2015 [22] | Brazil (Y) | Children | 1966 | 17D (Amaril), 17DD | PRNT50 | 1 month |
| Coursaget et al., 1995 [27] | Senegal (Y) | Children | 220 | 17D (Amaril) | PRNT90 | 1 month |
| Edupuganti et al., 2012 [28] | US (N) | Adults | 40 | 17D (YF-VAX) | PRNT90 | 3 months |
| Guirakhoo et al., 2006 [29] | US (N) | Adults | 42 | 17D (YF-VAX) | LNI | 3 years |
| Juan-Giner et al., 2021 [30] | Uganda, Kenya (Y) | Adults | 960 | 17D (different vaccines),<br>17DD | PRNT50,<br>PRNT90; non-<br>inferiority with<br>PRNT50 | 1 year |
| Lang et al., 1999 [31] | UK (N) | Adults | 185 | 17D (Stamaril, Arilvax) | PRNT80 | 1 month |
| Lopez et al., 2016 [32] | Colombia and<br>Peru (Y) | Children | 792 | 17D (Stamaril) | PRNT50 | 1 month |
| Monath et al., 2002 [33] | US (N) | Adults | 1440 | 17D (Arlvax, YF-VAX) | LNI | 1 month |
| Nasveld et al., 2010 [34] | US (N) | Adults | 90 | 17D (Stamaril) | PRNT50 | 6 months |
| NOVARTIS, 2012 [35] | Czech Republic,<br>Germany (N) | Adults | 101 | 17D (Stamaril) | NR | 1 month |
| Osei-Kwasi et al., 2001 [36] | Ghana (Y) | Children | 384 | 17D (Amaril) | microNT | 3 months |
| Roukens et al., 2008 [37] | Netherlands (N) | Adults | 175 | 17D (Stamaril) | PRNT80 | 1 year |
| Stefano et al., 1999 [38] | Brazil (Y) | Children | 294 | 17DD | PRNT50 | 1 month |
| **Non-RCT** |  |  |  |  |  |  |
| Avelino-Silva et al., 2016* [39] | Brazil (N) | Adults with HIV | 63 | NR | PRNT50 | 1 year |
| Avelino-Silva et al., 2016* [40] | Brazil (N) | Adults with HIV | 92 | 17DD | PRNT50 | 11 years |

| Study | Country (endemic region Y/N) | Subjects | n | Vaccine-specification | Specific Assay | Serology after YF vaccination up to |
| --- | --- | --- | --- | --- | --- | --- |
| Burkhard et al., 2020 [41] | Switzerland (N) | Adults with immunosuppressive therapy | 75 | NR | PRNT90/PRNT 80 | 31 years |
| Campi-Azevedo et al., 2016 [9] | Brazil (Y) | Adults | 171 | 17DD | PRNT50 | 13 years |
| Collaborative Group, 2019 [42] | Brazil (Y) | Adults | 323 | 17DD | PRNT50 | > 10 years |
| Collaborative Group, 2019 [43] | Brazil (Y) | Adults | 326 | 17DD | PRNT50 | 30 years |
| De Verdiere et al., 2018 [44] | France (N) | Adults with HIV | 71 | 17D (Stamaril) | PRNT80 | 1 year |
| Kerneis et al., 2013 [45] | France (N) | Adults under corticosteroids | 131 | 17D (Stamaril) | PRNT | 3 months |
| Michel et al., 2015 [46] | Senegal, French Guyana (N) | Children | 284 | 17D (different vaccines) | PRNT90 | 2 months |
| Project RETRO-CI, 1997 [47] | Ivory Coast (Y) | Children with HIV | 75 | 17D (Amaril) | PRNT | > 5 months |
| Roukens et al., 2011** [48] | Netherlands (N) | Adults | 58 | 17D (Stamaril) | PRNT80 | 10 years |
| Valim et al., 2020 [49] | Brazil (Y) | Adults with autoimmune diseases | 278 | 17DD | PRNT50 | 1 month |
| **Single arm studies** |  |  |  |  |  |  |
| Campi-Azevedo et al., 2019 [50] | Brazil (Y) | Children | 673 | 17DD | PRNT50 | 10 years |
| Domingo et al., 2019 [51] | Ghana, Mali (Y) | Children | 1023 | 17D (Stamaril), 17DD | microNT | 6 years |
| Idoko et al., 2020 [52] | Gambia, Mali (Y) | Children | 481 | 17D (Stamaril), 17DD | microNT | 6 years |
| Jia et al., 2019 [53] | China (N) | Adults | 2411 | Based on 17D (produced in China) | PRNT50 | 11 years |
| Kareko et al., 2018 [54] | US (N) | Adults | 92 | 17D (YF-VAX) | PRNT90 | 14 years |
| Veit et al., 2017 [55] | Switzerland (N) | Adults with HIV | 247 | 17D (different vaccines) | PRNT90 | 10 years |

\*Personal communication with Dr. Avelino-Silva confirmed that “the study populations in these two publications have some overlap but were not exactly the same”. Participants in Avelino-Silva 2016 [40] were previously vaccinated and referred for booster YF vaccine while some participants in Avelino-Silva 2016 [39] were previously included in Avelino-Silva 2016 [40] but some were vaccinated for the first time. In this systematic review, the publications were handled as two separate studies.
\*\*Updated information is available in Rosenstein et al. [56]
PRNT = Plaque reduction Neutralization Test; LNI = log<sub>10</sub> Neutralization Index; microNT = microNeutralization Test; HIV = Human Immunodeficiency Virus; NR = not reported

**Table 2:** Protection after single dose of YF vaccine - results of the meta-analysis.

| Forest Plot | Population | N studies | N data-points | N subjects | Effect estimate | 95% CI | I<sup>2</sup> |
| --- | --- | --- | --- | --- | --- | --- | --- |
| **≤3 months** |  |  |  |  |  |  |  |
| **001** | **Adults** | **16** | **27** | **3115** | **0.98** | **0.97 to 0.98** | **64%** |
| 002 | Endemic | 4 | 7 | 587 | 0.97 | 0.95 to 0.99 | 0% |
| 003 | Non-endemic | 12 | 20 | 2528 | 0.98 | 0.97 to 0.99 | 68% |
| **004** | **Children** | **10** | **21** | **5654** | **0.94** | **0.90 to 0.96** | **92%** |
| 005 | Endemic | 10 | 21 | 5654 | 0.94 | 0.90 to 0.96 | 92% |
| 006 | Non-endemic | 0 | 0 | 0 | N/A | N/A | N/A |
| **007** | **Immunodeficient** | **3** | **3** | **208** | **0.92** | **0.65 to 0.98** | **66%** |
| 008 | Endemic | 1 | 1 | 160 | 0.78 | 0.71 to 0.84 | N/A |
| 009 | Non-endemic | 2 | 2 | 48 | 0.98 | 0.84 to 1.00 | 0% |
| **>3 months to ≤5 years** |  |  |  |  |  |  |  |
| **010** | **Adults** | **8** | **13** | **790** | **0.97** | **0.95 to 0.98** | **0%** |
| 011 | Endemic | 3 | 6 | 514 | 0.98 | 0.95 to 0.99 | 0% |
| 012 | Non-endemic | 5 | 7 | 276 | 0.96 | 0.93 to 0.98 | 0% |
| **013** | **Children** | **3** | **4** | **1208** | **0.52** | **0.33 to 0.71** | **96%** |
| 014 | Endemic | 3 | 4 | 1208 | 0.52 | 0.33 to 0.71 | 96% |
| 015 | Non-endemic | 0 | 0 | 0 | N/A | N/A | N/A |
| **016** | **Immunodeficient** | **4** | **4** | **198** | **0.86** | **0.31 to 0.99** | **91%** |
| 017 | Endemic | 1 | 1 | 18 | 0.17 | 0.04 to 0.41 | N/A |
| 018 | Non-endemic | 3 | 3 | 180 | 0.94 | 0.77 to 0.99 | 56% |
| **>5 years to ≤10 years** |  |  |  |  |  |  |  |
| **019** | **Adults** | **6** | **7** | **267** | **0.88** | **0.78 to 0.93** | **53%** |
| 020 | Endemic | 2 | 2 | 45 | 0.88 | 0.75 to 0.95 | 0% |
| 021 | Non-endemic | 4 | 5 | 222 | 0.89 | 0.74 to 0.96 | 63% |
| **022** | **Children** | **3** | **3** | **1044** | **0.54** | **0.37 to 0.70** | **97%** |
| 023 | Endemic | 3 | 3 | 1044 | 0.54 | 0.37 to 0.70 | 97% |
| 024 | Non-endemic | 0 | 0 | 0 | N/A | N/A | N/A |
| **025** | **Immunodeficient** | **2** | **2** | **67** | **0.75** | **0.64 to 0.84** | **0%** |
| 026 | Endemic | 0 | 0 | 0 | N/A | N/A | N/A |
| 027 | Non-endemic | 2 | 2 | 67 | 0.75 | 0.64 to 0.84 | 0% |
| **>10 years to ≤20 years** |  |  |  |  |  |  |  |
| **028** | **Adults** | **4** | **4** | **231** | **0.71** | **0.62 to 0.79** | **36%** |
| 029 | Endemic | 3 | 3 | 193 | 0.74 | 0.63 to 0.82 | 43% |
| 030 | Non-endemic | 1 | 1 | 38 | 0.63 | 0.46 to 0.78 | N/A |
| **031** | **Children\*** | **0** | **0** | **0** | **N/A** | **N/A** | **N/A** |
| **034** | **Immunodeficient** | **1** | **1** | **8** | **0.62** | **0.24 to 0.91** | **N/A** |
| 035 | Endemic | 0 | 0 | 0 | N/A | N/A | N/A |
| 036 | Non-endemic | 1 | 1 | 8 | 0.62 | 0.24 to 0.91 | N/A |
| **>20 years** |  |  |  |  |  |  |  |
| **037** | **Adults** | **1** | **1** | **1** | **0.00** | **0.00 to 0.98** | **N/A** |
| 038 | Endemic | 0 | 0 | 0 | N/A | N/A | N/A |
| 039 | Non-endemic | 1 | 1 | 1 | 0.00 | 0.00 to 0.98 | N/A |
| **040** | **Children\*** | **0** | **0** | **0** | **N/A** | **N/A** | **N/A** |
| **043** | **Immunodeficient** | **1** | **1** | **17** | **0.94** | **0.71 to 1.00** | **N/A** |
| 044 | Endemic | 0 | 0 | 0 | N/A | N/A | N/A |
| 045 | Non-endemic | 1 | 1 | 17 | 0.94 | 0.71 to 1.00 | N/A |

\*No data

Titles and abstracts identified through electronic database and web searching were independently screened by 2 reviewers. Subsequently, full texts were independently examined by 2 reviewers to determine whether they met the criteria for inclusion in the review (see Supplementary Data for studies excluded at this stage) (Supplementary 2 -Excluded studies: Table 3). Any discrepancies between reviewers were resolved through discussion or a 3rd reviewer. The study selection process is detailed in accordance with the Preferred Reporting Items for Systematic Reviews and Meta-Analyses (PRISMA) statement [15].

**Table 3:** Protection after one booster dose of YF vaccine - results of the meta-analysis.

| Forest Plot | Population | N studies | N data-points | N subjects | Effect estimate | 95% CI | I2 |
| --- | --- | --- | --- | --- | --- | --- | --- |
| **≤3 months** |  |  |  |  |  |  |  |
| 046 | Adults | 2 | 2 | 64 | 0.98 | 0.89 to 1.00 | 0% |
| 047 | Endemic | 1 | 1 | 45 | 1.00 | 0.92 to 1.00 | N/A |
| 048 | Non-endemic | 1 | 1 | 19 | 1.00 | 0.82 to 1.00 | N/A |
| 049 | Children* | 0 | 0 | 0 | N/A | N/A | N/A |
| 052 | Immunodeficient | 1 | 1 | 11 | 1.00 | 0.72 to 1.00 | N/A |
| 053 | Endemic | 0 | 0 | 0 | N/A | N/A | N/A |
| 054 | Non-endemic | 1 | 1 | 11 | 1.00 | 0.72 to 1.00 | N/A |
| **>3 months to ≤5 years** |  |  |  |  |  |  |  |
| 055 | Adults | 2 | 2 | 62 | 0.92 | 0.82 to 0.97 | 0% |
| 056 | Endemic | 1 | 1 | 47 | 0.91 | 0.80 to 0.98 | N/A |
| 057 | Non-endemic | 1 | 1 | 15 | 1.00 | 0.78 to 1.00 | N/A |
| 058 | Children* | 0 | 0 | 0 | N/A | N/A | N/A |
| 061 | Immunodeficient* | 0 | 0 | 0 | N/A | N/A | N/A |
| **>5 years to ≤10 years** |  |  |  |  |  |  |  |
| 064 | Adults | 3 | 3 | 258 | 0.88 | 0.84 to 0.92 | 0% |
| 065 | Endemic | 2 | 2 | 249 | 0.88 | 0.83 to 0.91 | 0% |
| 066 | Non-endemic | 1 | 1 | 9 | 1.00 | 0.66 to 1.00 | N/A |
| 067 | Children* | 0 | 0 | 0 | N/A | N/A | N/A |
| 070 | Immunodeficient* | 0 | 0 | 0 | N/A | N/A | N/A |
| **>10 years to ≤20 years** |  |  |  |  |  |  |  |
| 073 | Adults | 2 | 2 | 17 | 0.86 | 0.61 to 0.96 | 0% |
| 074 | Endemic | 1 | 1 | 12 | 0.83 | 0.52 to 0.98 | N/A |
| 075 | Non-endemic | 1 | 1 | 5 | 1.00 | 0.48 to 1.00 | N/A |
| 076 | Children* | 0 | 0 | 0 | N/A | N/A | N/A |
| 079 | Immunodeficient* | 0 | 0 | 0 | N/A | N/A | N/A |
| **>20 years** |  |  |  |  |  |  |  |
| 082 | Adults* | 0 | 0 | 0 | N/A | N/A | N/A |
| 085 | Children* | 0 | 0 | 0 | N/A | N/A | N/A |
| 088 | Immunodeficient | 1 | 1 | 40 | 0.88 | 0.73 to 0.96 | N/A |
| 089 | Endemic | 0 | 0 | 0 | N/A | N/A | N/A |

| Forest Plot | Population | N studies | N data-points | N subjects | Effect estimate | 95% CI | I<sup>2</sup> |
| --- | --- | --- | --- | --- | --- | --- | --- |
| 090 | Non-endemic | 1 | 1 | 40 | 0.88 | 0.73 to 0.96 | N/A |

\*No data

### Data extraction

Data extraction forms were individually designed and piloted using Microsoft Excel. Data extraction was performed by 1 reviewer and checked for accuracy by a 2^nd^ reviewer. Any discrepancies were resolved through discussion or through the intervention of a 3^rd^ reviewer. Where necessary and feasible, we requested additional information from the authors (details available upon request).

### Risk of bias assessment

Risk of bias in randomised trials (RCT) was assessed using the Cochrane risk of bias tool [16]. For assessment of non-randomised studies we used the JBI (formerly Joanna Briggs Institute) checklist for non-randomised experimental studies [17]. Assessments were made by 2 reviewers independently and discrepancies were resolved through discussion or a 3^rd^ reviewer.

### Statistical analyses

A narrative summary of all included studies was prepared in tabular form. Where available, separate data were described for subgroups.

To investigate the duration of vaccine-induced protection against YF, data were stratified according to the follow-up time period after vaccination: ≤ 3 months; > 3 months to ≤ 5 years; > 5 to ≤ 10 years; > 10 to ≤ 20 years; > 20 years.

The outcome of interest was the proportion of people who were seropositive at a given time point post-vaccination. The presence of neutralizing antibody titers ≥ 1:10 in the serum neutralization assay (which is the reference test for the detection of humoral immunity to YF) is an established correlate of protection in vaccinated individuals [18-20] for which equivalent efficacy of the 17D- and its 17DD-substrain has been extensively demonstrated [11, 21, 22].

Meta-analyses were performed using the metaprop command with exact 95% confidence intervals (95% CI) in R version 4.1.0. Study arms not containing a YF vaccine (i.e. placebo) were excluded. If a study reported results at multiple time points for the same groups of participants, only the results for time-points closest to 3 months, 5 years, 10 years, and 20 years, respectively, were included. All analyses have been grouped by the follow-up time period after vaccination. Results for the larger datasets (e.g., for single dose applications and endemic regions) were additionally grouped by study design and person subgroups (adults, children, persons with immunodeficiency).

This review was registered in PROSPERO (CRD42020223939).

## Results

### Characteristics of the included studies

The systematic literature search revealed a total of 4,800 records. Thirty-six studies [9, 21-55] (reported in 61 references) met the inclusion criteria (see Supplementary Data) (Supplementary 3 - Included studies: Table 4). These studies comprised 18 RCT (reported in 32 references), 12 non-randomised comparative studies (non-RCT, reported in 18 references), and 6 single arm studies (reported in 11 references). For the study selection process see PRISMA flow chart in Supplementary Data (Supplementary 4 - PRISMA Flowchart: Figure 1).

**Table 4:** Protection after two or more booster doses of YF vaccine - results of the meta-analysis.

| Forest Plot | Population | N studies | N data-points | N subjects | Effect estimate | 95% CI | I<sup>2</sup> |
| --- | --- | --- | --- | --- | --- | --- | --- |
| **≤3 months** |  |  |  |  |  |  |  |
| 092 | Adults* | 0 | 0 | 0 | N/A | N/A | N/A |
| 095 | Children* | 0 | 0 | 0 | N/A | N/A | N/A |
| 097 | Immunodeficient* | 0 | 0 | 0 | N/A | N/A | N/A |
| **>3 months to ≤5 years** |  |  |  |  |  |  |  |
| 100 | Adults | 2 | 2 | 14 | 0.90 | 0.62 to 0.98 | 0% |
| 101 | Endemic | 2 | 2 | 14 | 0.90 | 0.62 to 0.98 | 0% |
| 102 | Non-endemic | 0 | 0 | 0 | N/A | N/A | N/A |
| 103 | Children* | 0 | 0 | 0 | N/A | N/A | N/A |
| 106 | Immunodeficient* | 0 | 0 | 0 | N/A | N/A | N/A |
| **>5 years to ≤10 years** |  |  |  |  |  |  |  |
| 109 | Adults* | 0 | 0 | 0 | N/A | N/A | N/A |
| 112 | Children* | 0 | 0 | 0 | N/A | N/A | N/A |
| 115 | Immunodeficient* | 0 | 0 | 0 | N/A | N/A | N/A |
| **>10 years to ≤20 years** |  |  |  |  |  |  |  |
| 118 | Adults* | 0 | 0 | 0 | N/A | N/A | N/A |
| 121 | Children* | 0 | 0 | 0 | N/A | N/A | N/A |
| 124 | Immunodeficient* | 1 | 1 | 2 | 1.00 | 0.16 to 1.00 | N/A |
| 125 | Endemic | 0 | 0 | 0 | N/A | N/A | N/A |
| 126 | Non-endemic | 1 | 1 | 2 | 1.00 | 0.16 to 1.00 | N/A |
| **>20 years** |  |  |  |  |  |  |  |
| 127 | Adults* | 0 | 0 | 0 | N/A | N/A | N/A |
| 130 | Children* | 0 | 0 | 0 | N/A | N/A | N/A |
| 133 | Immunodeficient | 1 | 1 | 3 | 1.00 | 0.29 to 1.00 | N/A |
| 134 | Endemic | 0 | 0 | 0 | N/A | N/A | N/A |
| 135 | Non-endemic | 1 | 1 | 3 | 1.00 | 0.29 to 1.00 | N/A |

\*No data

The included studies were conducted during 1993-2019 and entailed about 10,000 participants in 11 endemic and about 7,000 participants in 9 non-endemic countries, aged 6 months to 85 years (see Table 1 for study characteristics). All but one study in children examined children who received their YF vaccination before the age of 2. Eight studies included participants with immunodeficiencies (including HIV, autoimmune diseases, organ transplantation recipients and patients under immunosuppressive therapy for various reasons). The duration of protection beyond 3 months after YF vaccination was analysed in 20 studies. All included studies reported the titers of detectable neutralizing antibodies as surrogate markers for protection. None of the studies reported the proportion of individuals with clinical endpoints such as YF or death due to YF.

### Risk of Bias (RoB) assessment

Six of the 36 included studies (4 RCT and 2 non-randomized studies) had a low RoB, while RoB was high in 23 studies (10 RCT and 13 non-randomized studies). The remaining 7 studies were of unclear RoB (Supplementary Data) (Supplementary 5 - RoB Table: Table 5 and 6).

### Protection after single dose of YF vaccine

Up to 3 months: A total of 29 studies investigated the protection up to 3 months after a single dose of YF vaccine. In all groups with healthy individuals (adults [16 studies], children [10 studies]), pooled seroprotection rates were close to 100%. In persons with immunodeficiency (3 studies), pooled seroprotection rate was 92% (Table 2 and Supplementary Forest Plots 001-009) (Supplementary 6 - Forest Plots: Figures 001-009).

#### >3 months up to 5 years

Protection up to 5 years was addressed in 15 studies. In adults (8 studies), the pooled seroprotection rate remained as high as 97%. In contrast, the seroprotection rate was 52% in children (3 studies), with all 3 studies being conducted in endemic countries. In persons with immunodeficiency, the pooled seroprotection rate was slightly lower than for the preceding time interval (86%; 4 studies; Table 2 and Supplementary Forest Plots 010-014 and 016-018).

#### >5 years up to 10 years

Eleven studies addressed protection up to 10 years. In adults from both endemic and non-endemic countries, seroprotection rates were 88% (6 studies). In children, the respective value was 54% (3 studies in endemic settings). Two studies in persons with immunodeficiency in non-endemic settings showed a pooled seroprotection rate of 75% (Table 2 and Supplementary Forest Plots 019-023, 025 and 027).

#### >10 years up to 20 years

Protection up to 20 years was evaluated in 5 studies. In 4 studies (3 from endemic countries, 1 from a non-endemic country), the pooled seroprotection rate was 71% for healthy adults. No studies were conducted in children. The only study in immunodeficient persons in a non-endemic country showed a seroprotection rate of 62% (Table 2 and Supplementary Forest Plots 028-030, 034 and 036).

#### >20 years

The only study for this time period was performed in immunodeficient adults in a non-endemic setting and showed a seroprotection rate of 94% in 16 of 17 persons who had been vaccinated prior to immunosuppressive therapy (Table 2 and Supplementary Forest Plots 037, 039, 043 and 045).

### Protection after one booster dose of YF vaccine

Up to 3 months, seroprotection rates were 98% in adults and 100% in patients with immunodeficiency (Table 3 and Supplementary Forest Plots 046-048, 052 and 054). Between 3 months and 5 years after the booster dose, one study in an endemic and one in a non-endemic setting reported a pooled seroprotection rate of 92%. Up to 10 years after the booster dose, 3 studies in adults resulted in a pooled seroprotection rate of 88% (Table 3 and Supplementary Forest Plots 055-057 and 064-066).

Two studies in adults, one of which was performed in an endemic country, reported a pooled seroprotection rate of 86% for 10 to 20 years after booster dose (Table 3 and Supplementary Forest Plots 073-075).

For protection >20 years after booster dose, a study in immunodeficient persons receiving corticosteroid therapy showed a seroprotection rate of 88% (Table 3 and Supplementary Forest Plots 088 and 090).

In children, none of the studies met the inclusion criteria.

### Protection after two or more booster doses of YF vaccine

In two studies which investigated the protection from 3 months to 5 years after multiple booster doses in adults a pooled seroprotection rate of 90% was reported. For >10 years, one study which was performed in a non-endemic setting with patients receiving corticosteroid therapy, demonstrated a seroprotection rate of 100%. The participants had their first YF vaccination before the onset of immunosuppression (Table 4 and Supplementary Forest Plots 100, 101, 124, 126, 133 and 135).

### Subgroup analysis: Protection in immunocompromised persons

In persons with HIV, reduced antibody levels and a faster waning of YF immunity within 10 years were found compared to healthy controls, especially in patients with unsuppressed HIV RNA [44, 47, 55]. In persons with autoimmune diseases, seroprotection rates ranged between 73-85% one month after primary vaccination, depending on the underlying disease [49].

Another study in patients with autoimmune diseases examined the duration of immunity after 1 or more vaccine doses. All seronegative participants had only received a single YF vaccine dose. Those with 2 doses (or more) were seropositive for up to 33 years after the last dose [41] (Table 2, 3 and 4).

### Subgroup analysis: Protection in persons aged >60 years

Two studies reported on persons >60 years [48, 56]. A group of 28 persons was followed up 10 years after having received their first dose between 60-80 years of age. All of the 22 participants that could be contacted at the end of the observation period maintained a protective titer 10 years after primary vaccination [56].

### Subgroup analysis: Cross-reactive antibodies against other flavivirus

In one study with cross-reactive antibodies from a prior vaccination against Japanese encephalitis, an enhanced YF immunogenicity was detected after YF vaccination [26]. Crossreactive antibodies facilitated immune cell interactions and provoked greater pro-inflammatory responses.

## Discussion

This systematic review shows that the YF vaccine confers high rates of seroprotection within 3 months after primary vaccination. After a single vaccine dose, reduced seroprotection rates were observed 5 and 10 years after vaccination of healthy adults and 3 months to 5 years after vaccination of children. There is only scarce data on the persistence of humoral immunity beyond 10 years after a single YF vaccine dose. Beyond 20 years, no studies have been published in healthy adults. In immunodeficient persons, only limited data for different groups are available, which make general statements difficult. However, our subgroup analysis allows the conclusion that waning occurs in all groups examined. In the majority of studies, waning was more pronounced in immunodeficient persons than in healthy adults.

Only very limited data exist on the effects of booster vaccinations. For the time span beyond 20 years, either after a single YF vaccine dose or after booster doses, the only available data are derived from one study with immunodeficient persons. Due to the small sample size and a wide 95% CI, the results are difficult to interpret [41].

Our data revealed no epidemiologically relevant differences between endemic and non-endemic settings, suggesting that natural boosters in endemic settings are either rare or do not play a major role in maintaining protection.

The Strategic Advisory Group of Experts on Immunization, the principal advisory group to the WHO for vaccines and immunization, concluded in 2013 that YF booster doses are not needed for lifelong protection against YF in immunocompetent persons. The conclusion was based on a SR published in 2013 by Gotuzzo et al. [57]. However, this SR has been criticized for its methodological weaknesses. Other experts questioned the development of long-term protective immunity in a considerable proportion of those vaccinated with only one dose [8, 10, 12, 58]. The SR by Gotuzzo et al. mainly relies on retrospective cohort and small observational studies including case reports.

Since the data cut of the SR by Gotuzzo et al. [57], 23 additional studies have been published that were incorporated in our meta-analysis. To ascertain high quality evidence, we excluded retrospective studies, case reports and case series. Prospective single-arm studies were only accepted if they comprised ≥50 participants. Moreover, we included studies with different entities of immunodeficiency, such as patients seropositive for HIV (adults and children), patients with autoimmune diseases and organ transplantation recipients.

In persons with immunodeficiency, seroprotection after primary vaccination was only slightly lower than in healthy persons, but appeared to wane faster. Some countries recommend booster doses for patients with some but not all conditions leading to immunodeficiency [59]. The analysis of the various diseases associated with immunodeficiency supports to extend this recommendation to other patient groups with immunodeficiencies, provided that there are no contra-indications for a YF vaccination of the individual.

Persons aged 60 years and older often exhibit immunosenescence, which increases with age but also depends on other factors such as comorbidities. Although in one study all analysed participants aged >60 years still showed a protective YF titer 10 years after primary vaccination, the studied cohort was too small to draw firm conclusions [56].

The only study that measured antibodies against Japanese encephalitis [26] indicated a potential impact of antibodies cross-reactive with the YF virus, but the duration and relevance of this cross-reactivity remains uncertain. In endemic settings with high dengue seropositivity, pre-existing antibodies against dengue might lead to an overestimation of the seroprotection against YF.

One reason for no longer recommending routine booster doses was the extremely low number of reported YF vaccine failures. This rationale can be misleading, since vaccine failures can only be detected upon exposure of the vaccinated person to the YF virus as it occurs during outbreaks or in highly endemic sylvatic areas in Latin America. In addition, an underestimation of vaccine failures can result from insufficient local surveillance, case detection and reporting, especially in endemic countries in Africa, where 90% of all YF cases occur [4].

In Africa, reliable laboratory diagnostics are developing and recent examples, such as the YF outbreak in Uganda, might also show secondary vaccine failures [60].

For control and elimination of YF, it is crucial to improve the epidemiological surveillance not only for vaccine failures, but also for outbreaks [61].

### Strengths and limitations

Our review has several strengths. It adheres to rigorous methods recommended by relevant bodies such as Cochrane and the Centre for Reviews and Dissemination. With the last literature search performed in November 2021, our review reflects the current state of the evidence. With our meta-analysis we provide numerical estimates for protection at different time points after vaccination. High risk of bias and statistical heterogeneity of some of the included studies limits the ability to generate firm implications. The limitations associated with a functional assay and the variability among protocols represent a challenge for the comparison of results. As serum neutralization assays are usually only carried out in reference or specialized laboratories for the assessment of the immune response in vaccinated individuals, they certainly can provide a robust estimation on the presence of protective immunity against YF. Although we excluded prospective studies with less than 50 participants, for some study arms the number of subjects was too small to draw firm conclusions, which particularly applies to the studies with immunodeficient persons. Based on our data, it is not possible to make definitive statements on the necessity and impact of one or several YF booster doses in immunodeficient patients.

## Conclusions

A single dose of YF vaccine confers high levels of immunity (as measured by seroprotection rates) in healthy adults for up to 10 years, after which waning occurs, thereby increasing the risk for secondary vaccine failures. The extent to which immunity wanes depends on the age at primary vaccination and immune status.

## Supporting information

Supplementary Data 1 - Search strategies - Table 1 and 2

Supplementary Data 2 - Excluded studies - Table 3

Supplementary Data 3 - Included studies - Table 4

Supplementary Data 4 - PRISMA - Figure 1

Supplementary Data 5 - Risk of bias - Table 5 and 6

Supplementary Data 6 - Forest plots - Figures 001-135

## Data Availability

All data produced in the present work are contained in the manuscript and in the supplementary data

## Notes

### Author contributions

KK, TH, JK, and RW developed the protocol and SD developed the search strategy. KK, TH, CD and OW conceptualized the study, developed the PICO question and finalized the manuscript. KK, SD, JH, JK, KM, and RW reviewed full-text articles and abstracted the data. RW and KK performed the analysis. KK, CD and TH checked data extraction. KK wrote the first draft of the manuscript. CB provided immunological expertise and contributed to the writing of the paper. AWS contributed to the interpretation of data and writing of the final manuscript. TH supervised study conduct and all authors reviewed the manuscript for final revisions before submission.

## Acknowledgments

The authors thank Gerd Burchard, Annette Chalker, Annika Falman, Debra Fayter, Carol Forbes, Thomas Ledig, Thomas Mertens, Marianne Röbl-Mathieu, Jonas Schmidt-Chanasit, Ursula Wiedermann-Schmidt, and Gill Worthy for their valuable contribution to content.

