## Supplementary Data 1 - Search strategies - Table 1 and 2 for "Duration of protection after vaccination against yellow fever - systematic review and meta-analysis"

**Search strategies**

**Supplementary Table 1. Databases/resources searched 2020**

| **Database/**  **Resource** | **Host** | **Date range** | **Results** | **Date Searched** |
| --- | --- | --- | --- | --- |
| MEDLINE; MEDLINE In-Process Citations, Medline Daily Update, and Epub Ahead of Print | Ovid | 1946 to October 30, 2020 | 1654 | 2.11.20 |
| PubMed | NLM | up to 2 November 2020 | 33 | 2.11.20 |
| Embase | Ovid | 1974 to 2020 Week 43 | 3562 | 2.11.20 |
| Cochrane Central Register of Controlled Trials (CENTRAL) | Cochrane Library: Wiley | Issue 11 of 12, November 2020 | 262 | 2.11.20 |
| Cochrane Database of Systematic Reviews (CDSR) | Cochrane Library: Wiley | Issue 11 of 12, November 2020 | 2 | 2.11.20 |
| KSR Evidence | www.ksrevidence.com | Database last updated 2 Nov 2020 | 21 | 2.11.20 |
| Database of Abstracts of Reviews of Effects (DARE) | https://www.crd.york.ac.uk/  CRDWeb/ | up to 31 March 2015 | 2 | 2.11.20 |
| Health Technology Assessment Database (HTA) | https://www.crd.york.ac.uk/  CRDWeb/ | up to 31 March 2018 | 0 | 2.11.20 |
| PROSPERO | https://www.crd.york.ac.uk/  PROSPERO/ | up to 2 November 2020 | 49 | 2.11.20 |
| WHO Global Index Medicus | https://www.globalindexmedicus.net/ | up to 2 November 2020 | 351 | 2.11.20 |
| Northern Light Life Sciences Conference Abstracts database | Ovid | 2010 *-* 2020 Week 42 | 236 | 2.11.20 |
| ClinicalTrials.gov | http://clinicaltrials.gov | up to 2 November 2020 | 63 | 2.11.20 |
| WHO International Clinical Trials Register Portfolio (ICTRP) | http://www.who.int/ictrp/  search/en/ | up to 2 November 2020 | 76 | 2.11.20 |
| **Total records retrieved** | | | **6311** | |
| **Duplicate records removed** | | | **1823** | |
| **Total records to screen** | | | **4488** | |

**Search strategies. November 2020**

**MEDLINE and Epub Ahead of Print, In-Process & Other Non-Indexed Citations and Daily (Ovid): 1946 to October 30, 2020**

**Searched: 2.11.20**

1	(Yellow Fever/ or Yellow fever virus/) and (exp Vaccination/ or exp Immunization/) (678)

2	(yellow fever$ adj3 (jab$ or vaccin$ or revaccin$ or immuniz$ or immunis$ or reimmuniz$ or reimmunis$ or inoculat$ or shot$ or booster$)).ti,ab. (1302)

3	yellow fever vaccine/ (784)

4	((17D adj3 vaccin$) or (17DD adj3 vaccin$) or yf-vax or "yf vax" or stamaril or ap-yf or "ap yf" or BERNA-YF or flavimun or rki-yf or "rki yf" or "xrx 001" or xrx001).ti,ab,rn. (446)

5	or/1-4 (1854)

6	exp animals/ not humans/ (4751253)

**7	5 not 6 (1654)**

**PubMed (NLM): up to 2 November 2020**

<https://pubmed.ncbi.nlm.nih.gov/>

**Searched: 2.11.20**

**5	#3 AND #4	33**

4	pubstatusaheadofprint OR publisher[sb] OR pubmednotmedline[sb]	3,915,736

3	#1 AND #2	589

2	"17D vaccine" OR "17D vaccines" OR "17D vaccination" OR "17D vaccinations" OR "17DD vaccine" OR "17DD vaccines" or "17DD vaccination" or "17DD vaccinations" OR yf-vax OR "yf vax" OR stamaril OR ap-yf OR "ap yf" OR BERNA-YF OR flavimun OR rki-yf OR "rki yf" OR "xrx 001" OR xrx001	599

1	"yellow fever" AND (jab* or vaccin* or revaccin* or immuniz* or immunis* or reimmuniz* or reimmunis* or inoculat* or shot* or booster*)	2,786

**Embase (Ovid): 1974 to 2020 Week 43**

**Searched: 2.11.20**

1	(yellow fever/ or yellow fever virus/) and (exp vaccination/ or immunization/) (1631)

2	(yellow fever$ adj3 (jab$ or vaccin$ or revaccin$ or immuniz$ or immunis$ or reimmuniz$ or reimmunis$ or inoculat$ or shot$ or booster$)).ti,ab. (1456)

3	Yellow Fever Vaccine/ (2677)

4	((17D adj3 vaccin$) or (17DD adj3 vaccin$) or yf-vax or "yf vax" or stamaril or ap-yf or "ap yf" or BERNA-YF or flavimun or rki-yf or "rki yf" or "xrx 001" or xrx001).ti,ab,rn. (505)

5	or/1-4 (3711)

6	Animal experiment/ not (human experiment/ or human/) (2288426)

7	(rat or rats or mouse or mice or swine or porcine or murine or sheep or lambs or pigs or piglets or rabbit or rabbits or cat or cats or dog or dogs or cattle or bovine or monkey or monkeys or trout or marmoset$1).ti. and animal experiment/ (1086791)

**8	5 not (6 or 7) (3562)**

**Cochrane Database of Systematic Reviews (CDSR) (Wiley): Issue 11 of 12, November 2020**

**Cochrane Central Register of Controlled Trials (CENTRAL) (Wiley): Issue 11 of 12, November 2020**

**Searched: 2.11.20**

#1	MeSH descriptor: [Yellow Fever] this term only	58

#2	MeSH descriptor: [Yellow fever virus] this term only	28

#3	(yellow NEAR/3 fever*):ti,ab,kw	198

#4	#1 or #2 or #3	198

#5	MeSH descriptor: [Vaccination] explode all trees	2514

#6	MeSH descriptor: [Immunization] this term only	650

#7	(jab* or vaccin* or revaccin* or immuniz* or immunis* or reimmuniz* or reimmunis* or inoculat* or shot* or booster*):ti,ab,kw	31908

#8	#5 or #6 or #7	31908

#9	#4 and #8	179

#10	MeSH descriptor: [Yellow Fever Vaccine] this term only	37

#11	(17D* or yf-vax or "yf vax" or stamaril or ap-yf or "ap yf" or BERNA-YF or flavimun or rki-yf or "rki yf" or "xrx 001" or xrx001):ti,ab,kw	148

#12	#9 or #10 or #11	264

**CDSR	2 (2 reviews; 0 protocol)**

**CENTRAL	262**

**KSR Evidence (Internet): Database last updated 2 November 2020**

**www.ksrevidence.com**

**Searched: 2.11.20**

1	"yellow fever*" in All text	**21 results**

Database last updated Mon Nov 02 2020

**Database of Abstracts of Reviews of Effects (DARE) (CRD): up to 31 March 2015**

**Health Technology Assessment Database (HTA) (CRD): to 31 March 2018**

**http://www.crd.york.ac.uk/CRDWeb/**

**Searched: 2.11.20**

1	MeSH DESCRIPTOR Yellow Fever EXPLODE ALL TREES	1

2	MeSH DESCRIPTOR Yellow fever virus EXPLODE ALL TREES	1

3	(yellow NEAR fever*)	3

4	(17D* or yf-vax or "yf vax" or stamaril or ap-yf or "ap yf" or BERNA-YF or flavimun or rki-yf or "rki yf" or "xrx 001" or xrx001)	1

5	#1 OR #2 OR #3 OR #4	3

6	* IN DARE	4	5418

**7	#5 AND #6	2**

8	* IN HTA	17351

**9	#5 AND #8	0**

**PROSPERO (International prospective register of systematic reviews): up to 2 November 2020**

**https://www.crd.york.ac.uk/PROSPERO/**

**Searched: 2.11.20**

yellow fever	**49**

**WHO Global Index Medicus (GIM): up to 2 November 2020**

**https://www.globalindexmedicus.net/**

**Searched: 2.11.20**

(mh:(("Yellow Fever" OR "Yellow fever virus") AND ("Vaccination" OR "Immunization"))) OR (tw:("yellow fever" AND (jab* or vaccin* or revaccin* or immuniz* or immunis* or reimmuniz* or reimmunis* or inoculat* or shot* or booster*))) OR (mh:("Yellow Fever Vaccine")) OR (tw:("17D vaccine" OR "17D vaccines" OR "17D vaccination" OR "17D vaccinations" OR "17DD vaccine" OR "17DD vaccines" or "17DD vaccination" or "17DD vaccinations" OR yf-vax OR "yf vax" OR stamaril OR ap-yf OR "ap yf" OR BERNA-YF OR flavimun OR rki-yf OR "rki yf" OR "xrx 001" OR xrx001))

**Results 351**

**Northern Light Life Sciences Conference Abstracts (Ovid): 2010 – 2020 Week 42**

**Searched: 2.11.20**

1	Yellow Fever/ and Vaccines/ (156)

2	(yellow fever$ adj3 (jab$ or vaccin$ or revaccin$ or immuniz$ or immunis$ or reimmuniz$ or reimmunis$ or inoculat$ or shot$ or booster$)).ti,ab. (125)

3	Yellow Fever Vaccine/ (5)

4	((17D adj3 vaccin$) or (17DD adj3 vaccin$) or yf-vax or "yf vax" or stamaril or ap-yf or "ap yf" or BERNA-YF or flavimun or rki-yf or "rki yf" or "xrx 001" or xrx001).ti,ab. (34)

**5	or/1-4 (236)**

**ClinicalTrials.gov (Internet): up to 2 November 2020**

**http://clinicaltrials.gov/ct2/search/advanced**

**Searched: 2.11.20**

*Expert search option*

(("yellow fever" AND (vaccine OR vaccines OR vaccinate OR vaccination OR vaccinations OR revaccinate OR revaccination OR revaccinations OR immunization OR immunisation OR immunizations OR immunisations OR immunize OR immunise OR reimmunization OR reimmunisation OR reimmunizations OR reimmunisations OR reimmunize OR reimmunise OR shot OR shots OR booster OR boosters)) OR ("17D vaccine" OR "17D vaccines" OR "17D vaccination" OR "17D vaccinations" OR "17DD vaccine" OR "17DD vaccines" or "17DD vaccination" or "17DD vaccinations" OR yf-vax OR "yf vax" OR stamaril OR ap-yf OR "ap yf" OR BERNA-YF OR flavimun OR rki-yf OR "rki yf" OR "xrx 001" OR xrx001))

**63 Studies found**

**WHO International Clinical Trials Register Platform (ICTRP) (Internet): up to 2 Nov. 2020**

**https://apps.who.int/trialsearch/**

**Searched: 2.11.20**

yellow fever

**(83 records for) 76 trials found**

**Supplementary Table 2. Databases/resources searched. Update 2021**

| **Database/**  **Resource** | **Host** | **Date range** | **Results** | **Date Searched** |
| --- | --- | --- | --- | --- |
| MEDLINE; MEDLINE In-Process Citations, Medline Daily Update, and Epub Ahead of Print | Ovid | 1946 to November 11, 2021 | 107 | 12.11.21 |
| PubMed | NLM | up to 12 November 2021 | 11 | 12.11.21 |
| Embase | Ovid | 1974 to 2021 Week 44 | 233 | 12.11.21 |
| Cochrane Central Register of Controlled Trials (CENTRAL) | Cochrane Library: Wiley | Issue 11 of 12, November 2021 | 28 | 12.11.21 |
| Cochrane Database of Systematic Reviews (CDSR) | Cochrane Library: Wiley | Issue 11 of 12, November 2021 | 0 | 12.11.21 |
| KSR Evidence | www.ksrevidence.com | Database last updated 12 Nov 2021 | 6 | 12.11.21 |
| Database of Abstracts of Reviews of Effects (DARE)* | https://www.crd.york.ac.uk/  CRDWeb/ | up to 31 March 2015* | - | - |
| Health Technology Assessment Database (HTA)* | https://www.crd.york.ac.uk/  CRDWeb/ | up to 31 March 2018* | - | - |
| PROSPERO | https://www.crd.york.ac.uk/  PROSPERO/ | up to 12 November 2021 | 17 | 12.11.21 |
| WHO Global Index Medicus | https://www.globalindexmedicus.net/ | up to 12 November 2021 | 15 | 12.11.21 |
| Northern Light Life Sciences Conference Abstracts database | Ovid | 2010 *-* 2021 Week 44 | 17 | 12.11.21 |
| ClinicalTrials.gov | http://clinicaltrials.gov | up to 12 November 2021 | 8 | 12.11.21 |
| WHO International Clinical Trials Register Portfolio (ICTRP) | http://www.who.int/ictrp/  search/en/ | up to 12 November 2021 | 5 | 12.11.21 |
| **Total records retrieved** | | | **447** | |
| **Duplicate records removed** | | | **139** | |
| **Total records to screen** | | | **308** | |

*New records have not been added to DARE since March 2015 and HTA since March 2018.

**Search strategies. November 2021 update**

**MEDLINE and Epub Ahead of Print, In-Process & Other Non-Indexed Citations and Daily (Ovid): 1946 to November 11, 2021**

**Searched: 12.11.21**

1	(Yellow Fever/ or Yellow fever virus/) and (exp Vaccination/ or exp Immunization/) (721)

2	(yellow fever$ adj3 (jab$ or vaccin$ or revaccin$ or immuniz$ or immunis$ or reimmuniz$ or reimmunis$ or inoculat$ or shot$ or booster$)).ti,ab. (1388)

3	yellow fever vaccine/ (862)

4	((17D adj3 vaccin$) or (17DD adj3 vaccin$) or yf-vax or "yf vax" or stamaril or ap-yf or "ap yf" or BERNA-YF or flavimun or rki-yf or "rki yf" or "xrx 001" or xrx001).ti,ab,rn. (470)

5	or/1-4 (1963)

6	exp animals/ not humans/ (4913651)

**7	5 not 6 (1759)**

**PubMed (NLM): up to 12 November 2021**

<https://pubmed.ncbi.nlm.nih.gov/>

**Searched: 12.11.21**

**5	#3 AND #4	41**

4	pubstatusaheadofprint OR publisher[sb] OR pubmednotmedline[sb]	4,418,762

3	#1 AND #2	620

2	"17D vaccine" OR "17D vaccines" OR "17D vaccination" OR "17D vaccinations" OR "17DD vaccine" OR "17DD vaccines" or "17DD vaccination" or "17DD vaccinations" OR yf-vax OR "yf vax" OR stamaril OR ap-yf OR "ap yf" OR BERNA-YF OR flavimun OR rki-yf OR "rki yf" OR "xrx 001" OR xrx001	631

1	"yellow fever" AND (jab* or vaccin* or revaccin* or immuniz* or immunis* or reimmuniz* or reimmunis* or inoculat* or shot* or booster*)	2,963

**Embase (Ovid): 1974 to 2021 Week 44**

**Searched: 12.11.21**

1	(yellow fever/ or yellow fever virus/) and (exp vaccination/ or immunization/) (1754)

2	(yellow fever$ adj3 (jab$ or vaccin$ or revaccin$ or immuniz$ or immunis$ or reimmuniz$ or reimmunis$ or inoculat$ or shot$ or booster$)).ti,ab. (1545)

3	Yellow Fever Vaccine/ (2812)

4	((17D adj3 vaccin$) or (17DD adj3 vaccin$) or yf-vax or "yf vax" or stamaril or ap-yf or "ap yf" or BERNA-YF or flavimun or rki-yf or "rki yf" or "xrx 001" or xrx001).ti,ab,rn. (525)

5	or/1-4 (3928)

6	Animal experiment/ not (human experiment/ or human/) (2365551)

7	(rat or rats or mouse or mice or swine or porcine or murine or sheep or lambs or pigs or piglets or rabbit or rabbits or cat or cats or dog or dogs or cattle or bovine or monkey or monkeys or trout or marmoset$1).ti. and animal experiment/ (1126979)

**8	5 not (6 or 7) (3775)**

**Cochrane Database of Systematic Reviews (CDSR) (Wiley): Issue 11 of 12, November 2021**

**Cochrane Central Register of Controlled Trials (CENTRAL) (Wiley): Issue 11 of 12, November 2021**

**Searched: 12.11.21**

#1	MeSH descriptor: [Yellow Fever] this term only	64

#2	MeSH descriptor: [Yellow fever virus] this term only	29

#3	(yellow NEAR/3 fever*):ti,ab,kw	223

#4	#1 or #2 or #3	223

#5	MeSH descriptor: [Vaccination] explode all trees	2696

#6	MeSH descriptor: [Immunization] this term only	671

#7	(jab* or vaccin* or revaccin* or immuniz* or immunis* or reimmuniz* or reimmunis* or inoculat* or shot* or booster*):ti,ab,kw	34772

#8	#5 or #6 or #7	34772

#9	#4 and #8	201

#10	MeSH descriptor: [Yellow Fever Vaccine] this term only	43

#11	(17D* or yf-vax or "yf vax" or stamaril or ap-yf or "ap yf" or BERNA-YF or flavimun or rki-yf or "rki yf" or "xrx 001" or xrx001):ti,ab,kw	162

#12	#9 or #10 or #11	293

**CDSR	2 (2 reviews; 0 protocol)**

**CENTRAL	291**

**KSR Evidence (Internet): Database last updated 12 November 2021**

[www.ksrevidence.com](http://www.ksrevidence.com)

**Searched: 12.11.21**

1	"yellow fever*" in All text	**27 results**

Database last updated Fri Nov 12 2021

**Database of Abstracts of Reviews of Effects (DARE) (CRD): up to 31 March 2015**

**Health Technology Assessment Database (HTA) (CRD): to 31 March 2018**

**http://www.crd.york.ac.uk/CRDWeb/**

**Searched: 2.11.20**

1	MeSH DESCRIPTOR Yellow Fever EXPLODE ALL TREES	1

2	MeSH DESCRIPTOR Yellow fever virus EXPLODE ALL TREES	1

3	(yellow NEAR fever*)	3

4	(17D* or yf-vax or "yf vax" or stamaril or ap-yf or "ap yf" or BERNA-YF or flavimun or rki-yf or "rki yf" or "xrx 001" or xrx001)	1

5	#1 OR #2 OR #3 OR #4	3

6	* IN DARE	4	5418

**7	#5 AND #6	2**

8	* IN HTA	17351

**9	#5 AND #8	0**

**PROSPERO (International prospective register of systematic reviews): up to 12 November 2021**

<https://www.crd.york.ac.uk/PROSPERO/>

**Searched: 12.11.21**

yellow fever	65

**WHO Global Index Medicus (GIM): up to 12 November 2021**

**https://www.globalindexmedicus.net/**

**Searched: 12.11.21**

(mh:(("Yellow Fever" OR "Yellow fever virus") AND ("Vaccination" OR "Immunization"))) OR (tw:("yellow fever" AND (jab* or vaccin* or revaccin* or immuniz* or immunis* or reimmuniz* or reimmunis* or inoculat* or shot* or booster*))) OR (mh:("Yellow Fever Vaccine")) OR (tw:("17D vaccine" OR "17D vaccines" OR "17D vaccination" OR "17D vaccinations" OR "17DD vaccine" OR "17DD vaccines" or "17DD vaccination" or "17DD vaccinations" OR yf-vax OR "yf vax" OR stamaril OR ap-yf OR "ap yf" OR BERNA-YF OR flavimun OR rki-yf OR "rki yf" OR "xrx 001" OR xrx001))

**Results 377**

**Northern Light Life Sciences Conference Abstracts (Ovid): 2010 – 2021 Week 44**

**Searched: 12.11.21**

1	Yellow Fever/ and Vaccines/ (0)

2	(yellow fever$ adj3 (jab$ or vaccin$ or revaccin$ or immuniz$ or immunis$ or reimmuniz$ or reimmunis$ or inoculat$ or shot$ or booster$)).ti,ab. (153)

3	Yellow Fever Vaccine/ (8)

4	((17D adj3 vaccin$) or (17DD adj3 vaccin$) or yf-vax or "yf vax" or stamaril or ap-yf or "ap yf" or BERNA-YF or flavimun or rki-yf or "rki yf" or "xrx 001" or xrx001).ti,ab. (37)

**5	or/1-4 (163)**

**ClinicalTrials.gov (Internet): up to 12 November 2021**

**http://clinicaltrials.gov/ct2/search/advanced**

**Searched: 12.11.21**

*Expert search option*

(("yellow fever" AND (vaccine OR vaccines OR vaccinate OR vaccination OR vaccinations OR revaccinate OR revaccination OR revaccinations OR immunization OR immunisation OR immunizations OR immunisations OR immunize OR immunise OR reimmunization OR reimmunisation OR reimmunizations OR reimmunisations OR reimmunize OR reimmunise OR shot OR shots OR booster OR boosters)) OR ("17D vaccine" OR "17D vaccines" OR "17D vaccination" OR "17D vaccinations" OR "17DD vaccine" OR "17DD vaccines" or "17DD vaccination" or "17DD vaccinations" OR yf-vax OR "yf vax" OR stamaril OR ap-yf OR "ap yf" OR BERNA-YF OR flavimun OR rki-yf OR "rki yf" OR "xrx 001" OR xrx001))

**68 Studies found**

**WHO International Clinical Trials Register Platform (ICTRP) (Internet): up to 12 Nov. 2021**

**https://apps.who.int/trialsearch/**

**Searched: 12.11.21**

yellow fever

(91 records for) **85 trials found**
