## Supplementary Data 2 - Excluded studies - Table 3 for "Duration of protection after vaccination against yellow fever - systematic review and meta-analysis"

**Supplementary Table 3: Excluded studies with reasons for exclusion**

| **Citation** | **Comment** |
| --- | --- |
| Aarhus University Hospital. Immune response following vaccination against yellow fever. EUCTR2019-001731-31-DK. In: WHO International Clinical Trials Registry Platform (ICTRP) [Internet]. Geneva: World Health Organization (WHO). 2019 [accessed 02.11.20]. Available from: https://www.clinicaltrialsregister.eu/ctr-search/search?query=eudract_number:2019-001731-31 | No relevant outcome |
| Abreu A, Braga JU, Wigg L, Caetano R. Safety of yellow fever vaccine in elderly: systematic review. PROSPERO 2020 CRD42020160430. Available from: https://www.crd.york.ac.uk/prospero/display_record.php?ID=CRD42020160430 | No relevant study design |
| Academic Medical Center, Amsterdam, the Netherlands. Protection after yellow fever vaccination in patients using medication suppressing the immune system. NTR3581. In: WHO International Clinical Trials Registry Platform (ICTRP) [Internet]. Geneva: World Health Organization (WHO). 2012 [accessed 02.11.20]. Available from: https://trialregister.nl/trial/3430 | No relevant outcome |
| Adetokunboh O, Ndwandwe D, Awotiwon A, Uthman O. Evaluating the efficacy and effectiveness of vaccines among HIV–infected and HIV-exposed uninfected children: a systematic review and meta-analysis. PROSPERO 2018 CRD42018095334. Available from: https://www.crd.york.ac.uk/prospero/display_record.php?ID=CRD42018095334 | No relevant study design |
| Adler M, Lapierre V, Castilla-Llorente C, Bourhis J-H, Gachot B, Wyplosz B. Persistence of Yellow Fever Vaccine-Induced Antibodies After Allogeneic Haematopoietic Stem-Cell Transplantation. In: European Congress of Clinical Microbiology and Infectious Diseases 2017; 22-Apr-2017, 2017. Available from: European Society of Clinical Microbiology and Infectious Diseases (ESCMID) | No relevant study design |
| Adler M, Lapierre V, Sakr R, Bourhis JH, Gachot B, Castilla-Llorente C, et al. Persistence of yellow fever immunization-induced antibodies in allogeneic hematopoietic stem cell transplant recipients. J Infect Dis 2018;217(11):1844-5. | No relevant study design |
| Alberer M, Burchard G, Jelinek T, Reisinger E, Beran J, Hlavata LC, et al. Safety and immunogenicity of typhoid fever and yellow fever vaccines when administered concomitantly with quadrivalent meningococcal ACWY glycoconjugate vaccine in healthy adults. J Travel Med 2015;22(1):48-56. | No relevant information |
| Anderson CR, Gast-Galvis A. Immunity to yellow fever five years after vaccination. Am J Epidemiol 1947;45(3):302-4. | Un­obtainable |
| Avelino-Silva VI, Freire Mda S, Rocha V, Rodrigues CA, Novis YS, Sabino EC, et al. Persistence of yellow fever vaccine-induced antibodies after cord blood stem cell transplant. Hum Vaccin Immunother 2016;12(4):937-8. | No relevant study design |
| Avelino-Silva VI, Miyaji KT, Simoes M, Freire M, Sartori A, Hunt PW, et al. Immune activation impairs yellow fever vaccine efficacy in HIV-infected patients. Paper presented at 2015 Conference on Retroviruses and Opportunistic Infections, CROI 2015; 23-26 Feb 2015; Seattle: United States. Top Antivir Med 2015;23(E-1):133. | No relevant study design |
| Azamor T, da Silva AMV, Melgaco JG, Dos Santos AP, Xavier-Carvalho C, Alvarado-Arnez LE, et al. Activation of an effective immune response after yellow fever vaccination is associated with the genetic background and early response of IFN-γ and CLEC5A. Viruses 2021;13(1):96. | No relevant outcome |
| Baudon D, Robert V, Roux J. [The yellow fever epidemic in Burkina Faso in 1983]. Bull World Health Organ 1986;64(6):873-82. | No relevant study design |
| Bio-Manguinhos/Fiocruz (Brazil). Yellow fever vaccine dose-response study on children. ISRCTN36905484. In: WHO International Clinical Trials Registry Platform (ICTRP) [Internet]. Geneva: World Health Organization (WHO). 2011 [accessed 02.11.20]. Available from: http://isrctn.com/ISRCTN36905484 | No relevant outcome |
| Bouree P, Bisaro F. Yellow fever immunization for the solid organ recipients travellers. Paper presented at 12th conference of the International Society of Travel Medicine; 8-12 May 2011; Boston: United States. 2011. | No relevant outcome |
| Bovay A, Nassiri S, Maby-El Hajjami H, Marcos Mondejar P, Akondy RS, Ahmed R, et al. Minimal immune response to booster vaccination against yellow fever associated with pre-existing antibodies. Vaccine 2020;38(9):2172-82. | No relevant study design |
| Boyd AT, Dombaxe D, Moreira R, Oliveira MS, Manuel E, Colorado CN, et al. Notes from the field: investigation of patients testing positive for yellow fever viral RNA after vaccination during a mass yellow fever vaccination campaign - Angola, 2016. MMWR Morb Mortal Wkly Rep 2017;66(10):282-3. | No relevant study design |
| Brick IB. Residuals of yellow fever vaccine after ten years; a medical and legal problem. AMA Arch Intern Med 1953;92(2):221-7. | Un­obtainable |
| Buhler S, Jaeger VK, Eperon G, Furrer H, Fux CA, Jansen S, et al. Safety and immunogenicity of a primary yellow fever vaccination under low-dose methotrexate therapy - a prospective multi-centre pilot study. J Travel Med 2020;27(6):taaa126. | No relevant study design |
| Camacho LAB. Further Data On the Need For Booster Doses of Yellow Fever Vaccine (YFV). In: World Congress of Epidemiology 2014; 17-Aug-2014, 2014. Available from: World Congress of Epidemiology (WCE) https://discovery.northernlight.com/document.php?datasource=PHE&docid=PE20140811110001070&context=WK%40northernlight.com | No relevant study design |
| Campi-Azevedo A, Luiza-Silva M, Pacheco LP, Martins MA, Camacho LB, Homma A, et al. The 17D-213/77 seed-lot and the 17DD sub-strain of yellow fever vaccine trigger comparable overall cytokine signatures in vaccinated children. Paper presented at 15th Annual Conference on Vaccine Research; 7-9 May 2012; Baltimore: United States. 2012. | Un­obtainable |
| Caparroz ALMA, Trevisani VFM, Pileggi GCS. A systematic review of the safety and effectiveness of immunization on children and adolescents with chronic immune-mediated inflammatory diseases undergoing treatment with biologic and targeted synthetic disease-modifying antirheumatic drugs. PROSPERO 2019 CRD42019140927 Available from: https://www.crd.york.ac.uk/prospero/display_record.php?ID=CRD42019140927 | No relevant study design |
| Casey RM, Harris JB, Ahuka-Mundeke S, Dixon MG, Kizito GM, Nsele PM, et al. Immunogenicity of fractional-dose vaccine during a yellow fever outbreak - final report. N Engl J Med 2019;381(5):444-454. | No relevant study design |
| Centers for Disease Control and Prevention, Infectious Disease Institute, Kampala, Uganda, MRC/UVRI Uganda Research Unit on Aids, Ministry of Health, Uganda. Immunogenicity of fractional one-fifth and one-half doses of yellow fever vaccine compared to full dose in children 9-23 months old. In: ClinicalTrials.gov [Internet]. Bethesda (MD): National Library of Medicine (US). 2018-2019 [cited 2020 Nov 2]. Available from: http://clinicaltrials.gov/show/NCT03725618. NLM Identifier: NCT03725618 | Ongoing |
| Centre Hospitalier Universitaire Saint Pierre. "Persistence of neutralizing antibodies against yellow fever (YF) in HIV-infected patients". In: ClinicalTrials.gov [Internet]. Bethesda (MD): National Library of Medicine (US). 2015-2020 [cited 2020 Nov 2]. Available from: http://clinicaltrials.gov/show/NCT03591003. NLM Identifier: NCT03591003 | No relevant outcome |
| Centro de Pesquisas René Rachou- FIOCRUZ - Belo Horizonte, MG, Brazil. Immunity of the vaccine against yellow fever in patients taking medications that affect the immune system. RBR-946bv5. In: WHO International Clinical Trials Registry Platform (ICTRP) [Internet]. Geneva: World Health Organization (WHO). 2018 [accessed 02.11.20]. Available from: http://www.ensaiosclinicos.gov.br/rg/RBR-946bv5/ | Ongoing |
| Chan RC, Penney DJ, Little D, Carter IW, Roberts JA, Rawlinson WD. Hepatitis and death following vaccination with 17D-204 yellow fever vaccine. Lancet 2001;358(9276):121-2. | No relevant outcome |
| Chen LH, Wilson ME. Yellow fever control: current epidemiology and vaccination strategies. Trop Dis Travel Med Vaccines 2020;6:1. | No relevant study design |
| Collaborative Group for Studies on Yellow Fever Vaccines. Duration of post-vaccination immunity against yellow fever in adults. Vaccine 2014;32(39):4977-84. | No relevant study design |
| Coulange Bodilis H, Benabdelmoumen G, Gergely A, Goujon C, Pelicot M, Poujol P, et al. [Long term persistence of yellow fever neutralising antibodies in elderly persons]. Bull Soc Pathol Exot 2011;104(4):260-5. | No relevant study design |
| Courtois G. [Duration of immunity after yellow fever vaccination]. Ann Soc Belg Med Trop 1954;34(1):9-12. | Un­obtainable |
| Croce E, Hatz C, Jonker EF, Visser LG, Jaeger VK, Buhler S. Safety of live vaccinations on immunosuppressive therapy in patients with immune-mediated inflammatory diseases, solid organ transplantation or after bone-marrow transplantation - a systematic review of randomized trials, observational studies and case reports. Vaccine 2017;35(9):1216-26. | No relevant study design |
| da Silva VHIA. Avaliação da imunogenicidade e reatogenicidade da vacina contra febre amarela em pessoas que vivem com HIV [Internet]. São Paulo: Faculdade de Medicina da Universidade de São Paulo; 2015 [accessed 2.11.20]. Available from: http://www.teses.usp.br/teses/disponiveis/5/5134/tde-06012016-112024/publico/VivianHelenaIidaAvelinodaSilva.pdf | No relevant outcome |
| Dabrowska MM, Flisiak R. [Efficacy and safety of vaccination against yellow fever of persons traveling to endemic areas]. Przegl Epidemiol 2010;64(2):319-22. | No relevant study design |
| de Noronha TG, de Lourdes de Sousa Maia M, Geraldo Leite Ribeiro J, Campos Lemos JA, Maria Barbosa de Lima S, Martins-Filho OA, et al. Duration of post-vaccination humoral immunity against yellow fever in children. Vaccine 2019;37(48):7147-54. | No relevant study design |
| de Roever-Bonnet, Hoekstra J, van DJ. A follow-up of immunity after inoculation with 17D yellow fever vaccine. Tropical & Geographical Medicine 1962;14:361-74. | Un­obtainable |
| de Sousa MV, Zollner RL, Stucchi RSB, Boin IFSF, de Ataide EC, Mazzali M. Yellow fever disease in a renal transplant recipient: case report and literature review. Transpl Infect Dis 2019;21(5):e13151. | No relevant study design |
| De Verdiere NC, Durier C, Samri A, Launay O, Matheron S, Mercier-Delarue S, et al. Safety and immunogenicity of yellow fever vaccine in HIV-1-infected patients: ANRS EP46 NOVAA. Paper presented at 10th European Congress on Tropical Medicine and International Health; 16‐20 Oct 2017; Antwerp: Belgium. Trop Med Int Health 2017;22(Suppl 1):355-6. | No relevant study design |
| Dick GW, Gee FL. Immunity to yellow fever nine years after vaccination with 17D vaccine. Trans R Soc Trop Med Hyg 1952;46(4):449-58. | No relevant study design |
| Dick GW, Smithburn KC. Immunity to yellow fever 6 years after vaccination. Am J Trop Med Hyg 1949;29(1):57-61. | Un­obtainable |
| Diniz LMO, Romanelli RMDC, Bentes AA, Silva NLCD, Soares Cruzeiro FR, Marcial TM, et al. Yellow fever in children and adolescents previously immunized in Minas Gerais State, Brazil. Vaccine 2020;38(44):6954-6958. | No relevant study design |
| dos Santos AP. [Study of the immune response after vaccination against the yellow fever] [Internet]. Rio de Janeiro: Instituto Oswaldo Cruz; 2006 [accessed 2.11.20]. Available from: https://pesquisa.bvsalud.org/gim/resource/en/lil-453440 | Un­obtainable |
| Duclos P. Yellow fever vaccination: doing away with the ten yearly booster. Paper presented at International Society of Travel Medicine Biennial Conference 2015; 24-28 May 2015; Quebec City: Canada. 2015. | No relevant study design |
| Emory University, National Institutes of Health (NIH), National Institute of Allergy and Infectious Diseases (NIAID). Human immune responses to the yellow fever virus vaccine. In: ClinicalTrials.gov [Internet]. Bethesda (MD): National Library of Medicine (US). 2008- [cited 2020 Nov 2]. Available from: http://clinicaltrials.gov/show/NCT00694655. NLM Identifier: NCT00694655 | Ongoing |
| Epicentre, Kenya Medical Research Institute. Immunogenicity and safety of fractional doses of yellow fever vaccines (YEFE). In: ClinicalTrials.gov [Internet]. Bethesda (MD): National Library of Medicine (US). 2017-2018 [cited 2020 Nov 2]. Available from: http://clinicaltrials.gov/show/NCT02991495. NLM Identifier: NCT02991495 | Ongoing |
| Fantinato FFST, Duarte EC, Peixoto HM. Factors associated with vaccine failure with the yellow fever vaccine. A systematic review. PROSPERO 2020 CRD42020165079. Available from: https://www.crd.york.ac.uk/prospero/display_record.php?ID=CRD42020165079 | No relevant study design |
| Farnon EC, Gould LH, Griffith KS, Osman MS, El Kholy A, Brair ME, et al. Household-based sero-epidemiologic survey after a yellow fever epidemic, Sudan, 2005. Am J Trop Med Hyg 2010;82(6):1146-52. | No relevant study design |
| Ferreira CC, Campi-Azevedo AC, Peruhype-Magalhaes V, Coelho-Dos-Reis JG, Antonelli L, Torres K, et al. Impact of synthetic and biological immunomodulatory therapy on the duration of 17DD yellow fever vaccine-induced immunity in rheumatoid arthritis. Arthritis Res Ther 2019;21(1):75. | No relevant study design |
| Ferreira CC, Campi-Azevedo AC, Peruhype-Magalhaes V, Costa-Pereira C, Albuquerque CP, Muniz LF, et al. The 17D-204 and 17DD yellow fever vaccines: an overview of major similarities and subtle differences. Expert Rev Vaccines 2018;17(1):79-90. | No relevant study design |
| Figueiredo J, Moreira J, Brasil P, Siqueira A. Global risk assessment of travel-related yellow fever spread: a systematic review. Paper presented at 67th Annual Meeting of the American Society of Tropical Medicine and Hygiene, ASTMH 2018; 28 Oct-1 Nov 2018; New Orleans: United States. Am J Trop Med Hyg 2018;99(4 Suppl):66. | No relevant study design |
| Gibney KB, Edupuganti S, Panella AJ, Delorey MJ, Weaver B, Lanciotti RS, et al. Detection of yellow fever immunoglobulin m antibodies at 3-4 years following yellow fever vaccination. Paper presented at 12th conference of the International Society of Travel Medicine; 8-12 May 2011; Boston: United States. 2011. | No relevant study design |
| Gibney KB, Edupuganti S, Panella AJ, Kosoy OI, Delorey MJ, Lanciotti RS, et al. Detection of anti-yellow fever virus immunoglobulin m antibodies at 3-4 years following yellow fever vaccination. Am J Trop Med Hyg 2012;87(6):1112-5. | No relevant study design |
| Gomez SY, Ocazionez RE. [Yellow fever virus 17D neutralising antibodies in vaccinated Colombian people and unvaccinated ones having immunity against dengue]. Rev Salud Publica (Bogota) 2008;10(5):796-807. | No relevant outcome |
| Gotuzzo E, Yactayo S, Córdova E. Efficacy and duration of immunity after yellow fever vaccination: systematic review on the need for a booster every 10 years. Am J Trop Med Hyg 2013;89(3):434-44. | No relevant study design |
| Gowda R, Cartwright K, Bremner JA, Green ST. Yellow fever vaccine: a successful vaccination of an immunocompromised patient. Eur J Haematol 2004;72(4):299-301. | No relevant study design |
| Gowda R, Cartwright K, Bremner JA, Green ST. Yellow fever vaccine: a successful vaccination of an immunocompromised patient. Eur J Haematol 2004;72(4):299-301. | No relevant study design |
| Groot H, Riberiro RB. Neutralizing and haemagglutination-inhibiting antibodies to yellow fever 17 years after vaccination with 17D vaccine. Bull World Health Organ 1962;27:699-707. | No relevant study design |
| Hayakawa K, Takasaki T, Tsunemine H, Kanagawa S, Kutsuna S, Takeshita N, et al. Persistent seropositivity for yellow fever in a previously vaccinated autologous hematopoietic stem cell transplantation recipient. Int J Infect Dis 2015;37:9-10. | No relevant study design |
| Hospital Universitário Cassiano Antônio de Moraes - Vitória, ES, Brazil. Effectiveness and safety of yellow fever vaccination in patients with rheumatic diseases. RBR-3875DD. In: WHO International Clinical Trials Registry Platform (ICTRP) [Internet]. Geneva: World Health Organization (WHO). 2020 [accessed 02.11.20]. Available from: http://www.ensaiosclinicos.gov.br/rg/RBR-3875dd/ | Ongoing |
| Idoko OT, Mohammed N, Ansah P, Hodgson A, Tapia MD, Sow SO, et al. Antibody responses to yellow fever vaccine in 9 to 11-month-old Malian and Ghanaian children. Expert Rev Vaccines 2019;18(8):867-875. | No relevant study design |
| The Immunobiological Technology Institute (Bio-Manguinhos) / Oswaldo Cruz Foundation (Fiocruz). Immunity after two doses of yellow fever vaccine. In: ClinicalTrials.gov [Internet]. Bethesda (MD): National Library of Medicine (US). 2014-2015 [cited 2020 Nov 2]. Available from: http://clinicaltrials.gov/show/NCT02572518. NLM Identifier: NCT02572518 | No relevant outcome |
| The Immunobiological Technology Institute (Bio-Manguinhos) / Oswaldo Cruz Foundation (Fiocruz), Ministry of Health, Brazil. Immunity period after one dose of yellow fever vaccine in adults and children (Paraiba study). In: ClinicalTrials.gov [Internet]. Bethesda (MD): National Library of Medicine (US). 2016- [cited 2020 Nov 2]. Available from: http://clinicaltrials.gov/show/NCT02555072. NLM Identifier: NCT02555072 | Ongoing |
| The Immunobiological Technology Institute (Bio-Manguinhos) / Oswaldo Cruz Foundation (Fiocruz), Wellcome Trust. Duration of immunity 10 years after a dose-response study with yellow fever vaccine - complementary study. In: ClinicalTrials.gov [Internet]. Bethesda (MD): National Library of Medicine (US). 2019- [cited 2020 Nov 2]. Available from: http://clinicaltrials.gov/show/NCT04416477. NLM Identifier: NCT04416477 | Ongoing |
| Instituto de Investigacion de Enfermedades Tropicales de la Marina de Los E.E.U.U (NMRCD). Randomized, double-blind, pivotal study of phase III of the immunogenicity, security and comparative tolerance of two vaccines 17D against yellow fever (ARILVAX tm and YF-VAX tm) in infants and healthy children in Peru. PER-036-02. In: WHO International Clinical Trials Registry Platform (ICTRP) [Internet]. Geneva: World Health Organization (WHO). 2002 [accessed 02.11.20]. Available from: https://www.ins.gob.pe/ensayosclinicos/rpec/recuperarECPBNuevoEN.asp?numec=036-02 | No relevant outcome |
| Jean K, Donnelly CA, Ferguson NM, Garske T. A meta-analysis of serological response associated with yellow fever vaccination. Am J Trop Med Hyg 2016;95(6):1435-39. | No relevant study design |
| Jean K, Raad H, Gaythorpe KAM, Hamlet A, Mueller JE, Hogan D, et al. Assessing the impact of preventive mass vaccination campaigns on yellow fever outbreaks in Africa: a population-level self-controlled case series study. PLoS Med 2021;18(2):e1003523. | No relevant outcome |
| Kay A, Chen LH, Sisti M, Monath TP. Yellow fever vaccine seroconversion in travelers. Am J Trop Med Hyg 2011;85(4):748-9. | No relevant study design |
| Kernéis S, Launay O, Turbelin C, Batteux F, Hanslik T, Boelle PY. Long-term immune responses to vaccination in HIV-infected patients: a systematic review and meta-analysis. Clin Infect Dis 2014;58(8):1130-9. | No relevant study design |
| Kimathi D, Juan A, Bejon P, Grais RF, Warimwe GM, YEFE and NIFTY vaccine trials teams. Randomized, double-blinded, controlled non-inferiority trials evaluating the immunogenicity and safety of fractional doses of yellow fever vaccines in Kenya and Uganda. Wellcome Open Res 2019;4:182. | Ongoing |
| Kongsgaard M, Bassi MR, Rasmussen M, Skjodt K, Thybo S, Gabriel M, et al. Adaptive immune responses to booster vaccination against yellow fever virus are much reduced compared to those after primary vaccination. Sci Rep 2017;7(1):662. | No relevant study design |
| Lagos LWdA, Caetano R, Braga JU, Abreu A. Evaluation of yellow fever vaccine safety in immune depressed individuals: systematic review. PROSPERO 2020 CRD42020158807. Available from: https://www.crd.york.ac.uk/prospero/display_record.php?ID=CRD42020158807 | No relevant study design |
| Leiden University Medical Centre (LUMC) (Netherlands). Comparison between immune response to different modes of vaccination: intradermal and subcutaneous yellow fever vaccination. ISRCTN46326316. In: WHO International Clinical Trials Registry Platform (ICTRP) [Internet]. Geneva: World Health Organization (WHO). 2005 [accessed 02.11.20]. Available from: http://isrctn.com/ISRCTN46326316 | No relevant outcome |
| Leiden University Medical Center, Department of Infectious Diseases. Comparison between immune response to different modes of vaccination; intradermal and subcutaneous yellow fever vaccination. NTR231. In: WHO International Clinical Trials Registry Platform (ICTRP) [Internet]. Geneva: World Health Organization (WHO). 2005 [accessed 02.11.20]. Available from: https://trialregister.nl/trial/194 | No relevant outcome |
| Leiden University Medical Center. Seroprotection ten years after fractional dose yellow fever vaccination. NTR7094. In: WHO International Clinical Trials Registry Platform (ICTRP) [Internet]. Geneva: World Health Organization (WHO). 2018 [accessed 02.11.20]. Available from: https://trialregister.nl/trial/5528 | No relevant outcome |
| Licari A, Gertosio C, Silvestri AD, Rebuffi C, Marseglia GL, Chiappini E. What is the efficacy, immunogenicity, and safety of available vaccines in children with chronic conditions treated with biologic drugs? A systematic review and meta-analysis. PROSPERO 2020 CRD42020176227. Available from: https://www.crd.york.ac.uk/prospero/display_record.php?ID=CRD42020176227 | No relevant study design |
| Lindsey NP, Perry L, Fischer M, Woolpert T, Biggerstaff BJ, Brice G, et al. Duration of seropositivity following yellow fever vaccination in U.S. military service members. *Vaccine* 2020;38(52):8286-91. | No relevant study design |
| Machado VW, Vasconcelos PF, Silva EV, Santos JB. Serologic assessment of yellow fever immunity in the rural population of a yellow fever-endemic area in Central Brazil. Rev Soc Bras Med Trop 2013;46(2):166-71. | No relevant study design |
| Martin C, Domingo C, Bottieau E, Buonfrate D, De Wit S, Van Laethem Y, et al. Immunogenicity and duration of protection after yellow fever vaccine in people living with human immunodeficiency virus: a systematic review. Clin Microbiol Infect 2021;27(7):958-67. | No relevant study design |
| Martin C, Florence E, Delforge M, De Wit S, Domingo Carrasco C. Seroconversion rate after yellow fever vaccine in HIV positive patients. Paper presented at 17th European AIDS Conference; 6-9 Nov 2019; Basel: Switzerland. HIV Med 2019;20(Suppl 9):213. | No relevant study design |
| Medical Research Council. The impact of BCG vaccination on the response to other vaccines among Ugandan adolescents (POPVAC C). ISRCTN10482904. In: ISRCTN registry [Internet]. 2019 [accessed 2.11.20]. Available from: https://doi.org/10.1186/ISRCTN10482904 | No relevant outcome |
| Melo AK, Trevisani V, Pileggi G. A systematic review of the safety and effectiveness of immunization on patients with chronic immune-mediated inflammatory diseases undergoing treatment with biologic and targeted synthetic disease-modifying antirheumatic drugs. PROSPERO 2019 CRD4201913991. Available from: https://www.crd.york.ac.uk/prospero/display_record.php?ID=CRD42019139915 | No relevant study design |
| Miyaji KT, Avelino-Silva VI, Simoes M, Freire MD, Medeiros CR, Braga PE, et al. Prevalence and titers of yellow fever virus neutralizing antibodies in previously vaccinated adults. Rev Inst Med Trop Sao Paulo 2017;59:e2. | No relevant study design |
| Monath TP, Cetron MS, McCarthy K, Nichols R, Archambault WT, Weld L, et al. Yellow fever 17D vaccine safety and immunogenicity in the elderly. Hum Vaccin 2005;1(5):207-14. | No relevant study design |
| Monath TP, Fowler E, Johnson CT, Balser J, Morin MJ, Sisti M, et al. An inactivated cell-culture vaccine against yellow fever. *N Engl J Med* 2011;364(14):1326-33. | No relevant intervention |
| Monath TP, McCarthy K, Bedford P, Johnson CT, Nichols R, Yoksan S, et al. Clinical proof of principle for ChimeriVax: recombinant live, attenuated vaccines against flavivirus infections. Vaccine 2002;20(7-8):1004-18. | No relevant study design |
| Nash ER, Brand M, Chalkias S. Yellow fever vaccination of a primary vaccinee during adalimumab therapy. J Travel Med 2015;22(4):279-81. | No relevant study design |
| Nasidi A, Monath TP, Vandenberg J, Tomori O, Calisher CH, Hurtgen X, et al. Yellow fever vaccination and pregnancy: a four-year prospective study. Trans R Soc Trop Med Hyg 1993;87(3):337-9. | No relevant study design |
| Niedrig M, Lademann M, Emmerich P, Lafrenz M. Assessment of IgG antibodies against yellow fever virus after vaccination with 17D by different assays: neutralization test, haemagglutination inhibition test, immunofluorescence assay and ELISA. Trop Med Int Health 1999;4(12):867-71. | No relevant study design |
| Nnaji C, Adetokunboh O, Wiysonge C. A systematic review of the effects of fractional-dose yellow fever vaccine. PROSPERO 2018 CRD42018084214. Available from: https://www.crd.york.ac.uk/prospero/display_record.php?ID=CRD42018084214 | No relevant study design |
| Nnaji CA, Shey MS, Adetokunboh OO, Wiysonge CS. Immunogenicity and safety of fractional dose yellow fever vaccination: a systematic review and meta-analysis. Vaccine 2020;38(6):1291-1301. | No relevant study design |
| Odutola A, Ota MOC, Antonio M, Ogundare EO, Saidu Y, Owiafe PK, et al. Immunogenicity of pneumococcal conjugate vaccine formulations containing pneumococcal proteins, and immunogenicity and reactogenicity of co-administered routine vaccines - a phase II, randomised, observer-blind study in Gambian infants. Vaccine 2019;37(19):2586-99. | No relevant information |
| Ohmagari Norio. Stamaril research. JPRN-jRCTs031180027. In: WHO International Clinical Trials Registry Platform (ICTRP) [Internet]. Geneva: World Health Organization (WHO). 2018 [accessed 02.11.20]. Available from: https://jrct.niph.go.jp/latest-detail/jRCTs031180027 | No relevant outcome |
| Oliveira AC, Mota LM, Santos-Neto LL, Simoes M, Martins-Filho OA, Tauil PL. Seroconversion in patients with rheumatic diseases treated with immunomodulators or immunosuppressants, who were inadvertently revaccinated against yellow fever. Arthritis Rheumatol 2015;67(2):582-3. | No relevant study design |
| Omilabu SA, Adejumo JO, Olaleye OD, Fagbami AH, Baba SS. Yellow fever haemagglutination-inhibiting, neutralising and IgM antibodies in vaccinated and unvaccinated residents of Ibadan, Nigeria. Comp Immunol Microbiol Infect Dis 1990;13(2):95-100. | No relevant study design |
| Osinusi K, Akinkugbe FM, Akinwolere OA, Fabiyi A. Safety and efficacy of yellow fever vaccine in children less than one-year-old. West Afr J Med 1990;9(3):200-3. | Un­obtainable |
| Oswaldo Cruz Foundation. Immunogenicity and safety of the yellow fever vaccine in hiv infected individuals. In: ClinicalTrials.gov [Internet]. Bethesda (MD): National Library of Medicine (US). 2017-2018 [cited 2020 Nov 2]. Available from: http://clinicaltrials.gov/show/NCT03132311. NLM Identifier: NCT03132311 | Ongoing |
| Pacanowski J, Lacombe K, Campa P, Dabrowska M, Poveda JD, Meynard JL, et al. Plasma HIV-RNA is the key determinant of long-term antibody persistence after yellow fever immunization in a cohort of 364 HIV-infected patients. J Acquir Immune Defic Syndr 2012;59(4):360-7. | No relevant study design |
| Pfister M, Kursteiner O, Hilfiker H, Favre D, Durrer P, Ennaji A, et al. Immunogenicity and safety of BERNA-YF compared with two other 17D yellow fever vaccines in a phase 3 clinical trial. Am J Trop Med Hyg 2005;72(3):339-46. | No relevant information |
| Poland JD, Calisher CH, Monath TP, Downs WG, Murphy K. Persistence of neutralizing antibody 30-35 years after immunization with 17D yellow fever vaccine. Bull World Health Organ 1981;59(6):895-900. | No relevant study design |
| Receveur MC, Thiebaut R, Vedy S, Malvy D, Mercie P, Bras ML. Yellow fever vaccination of human immunodeficiency virus-infected patients: report of 2 cases. Clin Infect Dis 2000;31(3):E7-8. | No relevant study design |
| Alba Maria Ropero, Centers for Disease Control and Prevention, Ministry of Public Health, Argentina. Immunogenicity of co-administered yellow fever and measles, mumps, and rubella (MMR) vaccines. In: ClinicalTrials.gov [Internet]. Bethesda (MD): National Library of Medicine (US). 2015-2018 [cited 2020 Nov 2]. Available from: http://clinicaltrials.gov/show/NCT03368495. NLM Identifier: NCT03368495 | No relevant outcome |
| AHE Roukens and LG Visser. Immune response ten years after yellow fever vaccination in the elderly traveller. NL8079. In: WHO International Clinical Trials Registry Platform (ICTRP) [Internet]. Geneva: World Health Organization (WHO). 2019 [accessed 02.11.20]. Available from: https://trialregister.nl/trial/8079 | Ongoing |
| Roukens AHE, Visser LG. Fractional-dose yellow fever vaccination: an expert review. J Travel Med 2019;26(6):taz024. | No relevant study design |
| Saad S, Bark D, Kitchin V, Sadarangani M. Efficacy, effectiveness, immunogenicity, and safety of vaccination in hematopoietic stem cell transplant recipients: a systematic review. PROSPERO 2020 CRD42020182137. Available from: https://www.crd.york.ac.uk/prospero/display_record.php?ID=CRD42020182137 | No relevant study design |
| Sanofi Pasteur, a Sanofi Company. Dose-ranging study of an investigational yellow fever candidate vaccine in adults. In: ClinicalTrials.gov [Internet]. Bethesda (MD): National Library of Medicine (US). 2020- [cited 2020 Nov 2]. Available from: http://clinicaltrials.gov/show/NCT04142086. NLM Identifier: NCT04142086 | Ongoing |
| Sanofi Pasteur, a Sanofi Company. Immune response to different schedules of a tetravalent dengue vaccine given with or without yellow fever vaccine. In: ClinicalTrials.gov [Internet]. Bethesda (MD): National Library of Medicine (US). 2011-2013 [cited 2020 Nov 2]. Available from: http://clinicaltrials.gov/show/NCT01488890. NLM Identifier: NCT01488890 | No relevant outcome |
| Sanofi Pasteur Inc. Immunogenicity and safety of yellow fever vaccine (Stamaril®) administered concomitantly with tetravalent dengue vaccine in healthy toddlers at 12-13 months of age in Colombia and Peru. PER-037-11. In: WHO International Clinical Trials Registry Platform (ICTRP) [Internet]. Geneva: World Health Organization (WHO). 2011 [accessed 02.11.20]. Available from: https://www.ins.gob.pe/ensayosclinicos/rpec/recuperarECPBNuevoEN.asp?numec=037-11 | No relevant outcome |
| Santos AP, Bertho AL, Dias DC, Santos JR, Marcovistz R. Lymphocyte subset analyses in healthy adults vaccinated with yellow fever 17DD virus. Mem Inst Oswaldo Cruz 2005;100(3):331-7. | No relevant study design |
| Schnyder JL, De Pijper CA, Garcia Garrido HM, Daams JG, Goorhuis A, Stijnis C, et al. Fractional dose of intradermal compared to intramuscular and subcutaneous vaccination - a systematic review and meta-analysis. Travel Med Infect Dis 2020;37:101868. | No relevant study design |
| Schnyder J, Pijper CD, Stijnis C, Schaumburg F, Grobusch M. Fractional dose of intradermal compared to intramuscular and subcutaneous vaccination: a systematic review. PROSPERO 2020 CRD42020151725. Available from: https://www.crd.york.ac.uk/prospero/display_record.php?ID=CRD42020151725 | No relevant study design |
| Sicre de Fontbrune F, Arnaud C, Cheminant M, Boulay A, Konopacki J, Lapusan S, et al. Immunogenicity and safety of yellow fever vaccine in allogeneic hematopoietic stem cell transplant recipients after withdrawal of immunosuppressive therapy. J Infect Dis 2018;217(3):494-7. | No relevant study design |
| Sicre De Fontbrune F, Arnaud C, Cheminant M, Konopacki J, Lapusan S, Boulay A, et al. Efficacy and safety of yellow fever vaccine in allogeneic SCT recipients after withdrawal of immunosuppression. Paper presented at 42nd Annual Meeting of the European Society for Blood and Marrow Transplantation, EBMT 2016; 3-6 Apr 2016; Valencia: Spain. Bone Marrow Transplant 2016;51(Suppl):S204-S205. | No relevant study design |
| Sidibe M, Yactayo S, Kalle A, Sall AA, Sow S, Ndoutabe M, et al. Immunogenicity and safety of yellow fever vaccine among 115 HIV-infected patients after a preventive immunisation campaign in Mali. Trans R Soc Trop Med Hyg 2012;106(7):437-44. | No relevant study design |
| Silva JVJ, Jr., Lopes TRR, Oliveira-Filho EF, Oliveira RAS, Duraes-Carvalho R, Gil LHVG. Current status, challenges and perspectives in the development of vaccines against yellow fever, dengue, Zika and chikungunya viruses. Acta Trop 2018;182:257-63. | No relevant study design |
| Silva ML. Caracterização da resposta vacinal antiamarílica em crianças e adultos, utilizando o modelo panorâmico de análise imunofenotípica [Internet]. Belo Horizonte: Saúde do Centro de Pesquisas René Rachou; 2011 [accessed 2.11.20]. Available from: http://www.cpqrr.fiocruz.br/texto-completo/T_33.pdf | No relevant outcome |
| Simoes M, Camacho LAB, Yamamura AMY, Miranda EH, Cajaraville ACRA, da Silva Freire M. Evaluation of accuracy and reliability of the plaque reduction neutralization test (micro-PRNT) in detection of yellow fever virus antibodies. Biologicals 2012;40(6):399-404. | No relevant study design |
| Smith CE, McMahon DA, Turner LH. Yellow fever vaccination in malaya by subcutaneous injection and multiple puncture. Haemagglutinin-inhibiting antibody responses in persons with and without pre-existing antibody. Bull World Health Organ 1963;29:75-80. | No relevant information |
| Song R, Guan S, Lee SS, Chen Z, Chen C, Han L, et al. Late or lack of vaccination linked to importation of yellow fever from Angola to China. Emerg Infect Dis 2018;24(7):1383-6. | No relevant study design |
| Sow A, Faye O, Diallo M, Ndiaye Y, Ba IO, Yactayo S, et al. Yellow fever immunity assessment in Kedougou, South Eastern Senegal, in 2012. Paper presented at 16th International Congress on Infectious Diseases, ICID 2014; 2-5 Apr 2014; Cape Town: South Africa. Int J Infect Dis 2014;21(Suppl 1):253. | No relevant information |
| Staples JE, Barrett ADT, Wilder-Smith A, Hombach J. Review of data and knowledge gaps regarding yellow fever vaccine-induced immunity and duration of protection. NPJ Vaccines 2020;5:54. | No relevant study design |
| Staples JE, Bocchini JA, Jr., Rubin L, Fischer M, Centers for Disease and Control Prevention. Yellow fever vaccine booster doses: recommendations of the Advisory Committee on Immunization Practices, 2015. MMWR Morb Mortal Wkly Rep 2015;64(23):647-50. | No relevant study design |
| Stoffella-Dutra AG, Silva de Oliveira J, Barbosa Costa G, Geessien Kroon E, Santos Abrahao J, Desiree LaBeaud A, et al. Absence of YF-neutralizing antibodies in vulnerable populations of Brazil: a warning for epidemiological surveillance and the potential risks for future outbreaks. Vaccine 2020;38(42):6592-9. | No relevant study design |
| Stuhec M. Yellow fever vaccine used in a psoriatic arthritis patient treated with methotrexate: a case report. Acta dermatovenerolog 2014;23(3):63-4. | No relevant outcome |
| Suzano CE, Amaral E, Sato HK, Papaiordanou PM, Campinas Group on Yellow Fever Immunization during Pregnancy. The effects of yellow fever immunization (17DD) inadvertently used in early pregnancy during a mass campaign in Brazil. Vaccine 2006;24(9):1421-6. | No relevant study design |
| Takeda. Immunogenicity and safety of tetravalent dengue vaccine (TDV) administered with a yellow fever vaccine in adults. In: ClinicalTrials.gov [Internet]. Bethesda (MD): National Library of Medicine (US). 2018-2019 [cited 2020 Nov 2]. Available from: http://clinicaltrials.gov/show/NCT03342898. NLM Identifier: NCT03342898 | Ongoing |
| Takey PRG, Brasil P, Guaraldo L, Pedro RS. Effectiveness and safety of the yellow fever vaccine (attenuated): systematic review and meta-analysis. PROSPERO 2020 CRD42020157929. Available from: https://www.crd.york.ac.uk/prospero/display_record.php?ID=CRD42020157929 | No relevant study design |
| Tattevin P, Depatureaux AG, Chapplain JM, Dupont M, Souala F, Arvieux C, et al. Yellow fever vaccine is safe and effective in HIV-infected patients. AIDS 2004;18(5):825-7. | No relevant study design |
| Tokyo Medical University Hospital. Long-term immunity after Yellow fever vaccination. JPRN-UMIN000040526. In: WHO International Clinical Trials Registry Platform (ICTRP) [Internet]. Geneva: World Health Organization (WHO). 2020 [accessed 02.11.20]. Available from: https://upload.umin.ac.jp/cgi-open-bin/ctr_e/ctr_view.cgi?recptno=R000046155 | Ongoing |
| University of Oxford, KEMRI-Wellcome Trust Collaborative Research Program, Institut Pasteur, MRC/UVRI Uganda Research Unit on Aids, Epicentre, Paris, France. Non-inferiority fractional-doses trial for yellow fever vaccine. In: ClinicalTrials.gov [Internet]. Bethesda (MD): National Library of Medicine (US). 2019- [cited 2020 Nov 2]. Available from: http://clinicaltrials.gov/show/NCT04059471. NLM Identifier: NCT04059471 | Ongoing |
| University of Sao Paulo General Hospital. Yellow fever vaccine in patients with rheumatic diseases. In: ClinicalTrials.gov [Internet]. Bethesda (MD): National Library of Medicine (US). 2018-2019 [cited 2020 Nov 2]. Available from: http://clinicaltrials.gov/show/NCT03430388. NLM Identifier: NCT03430388 | No relevant outcome |
| University of Zurich, Swiss Tropical & Public Health Institute, Kantonsspital Aarau, University Hospital Inselspital, Berne, University Hospital, Geneva, Centre Hospitalier Universitaire Vaudois. Yellow fever vaccination under low dose methotrexate therapy. In: ClinicalTrials.gov [Internet]. Bethesda (MD): National Library of Medicine (US). 2014-2016 [cited 2020 Nov 2]. Available from: http://clinicaltrials.gov/show/NCT02383680. NLM Identifier: NCT02383680 | No relevant study design |
| Valim V, Gouvea SA, Lima SMB, Azevedo ACC, Carvalho AT, Pascoal VPM, et al. Effectiveness and safety of yellow fever vaccine in patients with primary Sjogren's syndrome. Paper presented at 14th International Symposium on Sjogren's Syndrome; 17-20 Apr 2018; Washington DC: United States. Clin Exp Rheumatol 2018;36(3 Suppl 112):S326. | No relevant outcome |
| Vasconcelos PFC, Barrett ADT. Are booster doses of yellow fever vaccine needed? Lancet Infect Dis 2019;19(12):1275-6. | No relevant study design |
| Wieten RW, Goorhuis A, Jonker EFF, de Bree GJ, de Visser AW, van Genderen PJJ, et al. 17D yellow fever vaccine elicits comparable long-term immune responses in healthy individuals and immune-compromised patients. J Infect 2016;72(6):713-22. | No relevant study design |
| Wieten RW, Jonker E, De Bree G, Goorhuis A, Visser LG, Van Leeuwen E, et al. Yellow fever vaccination immune responses are measurable up to 38 years after vaccination. Paper presented at 16th International Congress on Infectious Diseases, ICID 2014; 2-5 Apr 2014; Cape Town: South Africa. Int J Infect Dis 2014;21(Suppl 1):437. | No relevant outcome |
| Wieten RW, Jonker EF, Pieren DK, Hodiamont CJ, van Thiel PP, van Gorp EC, et al. Comparison of the PRNT and an immune fluorescence assay in yellow fever vaccinees receiving immunosuppressive medication. Vaccine 2016;34(10):1247-51. | No relevant study design |
| Wieten RW, Jonker EF, van Leeuwen EM, Remmerswaal EB, Ten Berge IJ, de Visser AW, et al. A single 17D yellow fever vaccination provides lifelong immunity; characterization of yellow-fever-specific neutralizing antibody and T-cell responses after vaccination. PLoS One 2016;11(3):e0149871. | No relevant study design |
| Willcox AC, Collins MH, Jadi R, Keeler C, Parr JB, Mumba D, et al. Seroepidemiology of dengue, Zika, and yellow fever viruses among children in the Democratic Republic of the Congo. Am J Trop Med Hyg 2018;99(3):756-63. | No relevant study design |
| Wilder-Smith A, Barrett A, Vannice K, Hombach J. Long-term protection after fractional-dose yellow fever vaccination. Ann Intern Med 2019;171(2):145-6. | No relevant study design |
| Wisseman CL, Jr., Sweet BH. Immunological studies with group B arthropod-borne viruses. III. Response of human subjects to revaccination with 17D strain yellow fever vaccine. Am J Trop Med Hyg 1962;11(4):570-5. | Un­obtainable |
| Wyplosz B, Burdet C, Francois H, Durrbach A, Duclos-Vallee JC, Mamzer-Bruneel MF, et al. Persistence of yellow fever vaccine-induced antibodies after solid organ transplantation. Am J Transplant 2013;13(9):2458-61. | No relevant study design |
| Wyplosz B, Burdet C, Francois H, Durrbach A, Duclos-Vallee JC, Mamzer-Bruneel MF, et al. Persistence of yellow fever vaccine induced antibodies after solid organ transplantation. Paper presented at European Congress of Clinical Microbiology and Infectious Diseases 2014; 10-13 May 2014; Barcelona: Spain. 2014. | No relevant study design |
| Yamoah P, Bangalee V, Oosthuizen F. A review of the safety of vaccines used in routine immunization in Africa. Afr Health Sci 2020;20(1):227-37. | No relevant study design |
| Zhao, L., Miao, F., Chen, T., Du, H. & Zhao, J. Stablity of yellow fever virus neutralising antibody titres. Lancet Infect Dis 20, 166-167, doi:http://dx.doi.org/10.1016/S1473-3099%2819%2930703-0 (2020). | No relevant study design |
