## Supplementary Data 3 - Included studies - Table 4 for "Duration of protection after vaccination against yellow fever - systematic review and meta-analysis"

**Supplementary Table 4: Included studies**

| **Study** | **Reference** |
| --- | --- |
| **Randomised controlled trials (18 studies, 32 references)** | |
| Asante 2020  (NCT02699099) | Asante KP, Ansong D, Kaali S, Adjei S, Lievens M, Nana Badu L, et al. Immunogenicity and safety of the RTS,S/AS01 malaria vaccine co-administered with measles, rubella and yellow fever vaccines in Ghanaian children: a phase IIIb, multi-center, non-inferiority, randomized, open, controlled trial. Vaccine 2020;38(18):3411-21. |
|  | GlaxoSmithKline. Immunogenicity and safety study of GSK biologicals' candidate malaria vaccine given at 6, 7.5 and 9 months of age in co-administration with measles, rubella and yellow fever (YF) vaccines followed by a booster of the malaria vaccine. In: ClinicalTrials.gov [Internet]. Bethesda (MD): National Library of Medicine (US). 2017-2020 [cited 2020 Nov 2]. Available from: https://ClinicalTrials.gov/show/NCT02699099. NLM Identifier: NCT02699099 |
| Belmusto-Worn 2005 | Belmusto-Worn VE, Sanchez JL, McCarthy K, Nichols R, Bautista CT, Magill AJ, et al. Randomized, double-blind, phase III, pivotal field trial of the comparative immunogenicity, safety, and tolerability of two yellow fever 17D vaccines (Arilvax and YF-VAX) in healthy infants and children in Peru. Am J Trop Med Hyg 2005;72(2):189-97. |
| Camacho 2004 | Camacho LA, de Aguiar SG, Freire Mda S, Leal Mda L, do Nascimento JP, Iguchi T, et al. Reactogenicity of yellow fever vaccines in a randomized, placebo-controlled trial. Rev Saude Publica 2005;39(3):413-20. |
|  | Camacho LAB, Freire Mda S, Leal Mda L, de Aguiar SG, do Nascimento JP, Iguchi T, et al. Immunogenicity of WHO-17D and Brazilian 17DD yellow fever vaccines: a randomized trial. Rev Saude Publica 2004;38(5):671-8. |
| Campi-Azevedo 2014  (NCT0338231; ISRCTN38082350) | de Menezes Martins R, Maia MLS, de Lima SMB, de Noronha TG, Xavier JR, Camacho LAB, et al. Duration of post-vaccination immunity to yellow fever in volunteers eight years after a dose-response study. Vaccine 2018;36(28):4112-7. |
|  | da Costa-Rocha IA, Campi-Azevedo AC, Peruhype-Magalhaes V, Coelho-Dos-Reis JG, Fradico JRB, Souza-Lopes T, et al. Duration of humoral and cellular immunity 8 years after administration of reduced doses of the 17DD-yellow fever vaccine. Front Immunol 2019;10:1211. |
|  | Campi-Azevedo AC, de Almeida Estevam P, Coelho-Dos-Reis JG, Peruhype-Magalhaes V, Villela-Rezende G, Quaresma PF, et al. Subdoses of 17DD yellow fever vaccine elicit equivalent virological/immunological kinetics timeline. BMC Infect Dis 2014;14:391. |
|  | Martins RM, Maia Mde L, Farias RH, Camacho LA, Freire MS, Galler R, et al. 17DD yellow fever vaccine: a double blind, randomized clinical trial of immunogenicity and safety on a dose-response study. Hum Vaccin Immunother 2013;9(4):879-88. |
|  | The Immunobiological Technology Institute (Bio-Manguinhos) / Oswaldo Cruz Foundation (Fiocruz). Clinical study of immunity duration of yellow fever vaccine in military. In: ClinicalTrials.gov [Internet]. Bethesda (MD): National Library of Medicine (US). 2017-2017 [cited 2020 Nov 2]. Available from: http://clinicaltrials.gov/show/NCT03338231. NLM Identifier: NCT03338231 |
|  | Bio-Manguinhos/Fiocruz (Brazil). Yellow fever vaccine dose-response study. ISRCTN38082350. In: WHO International Clinical Trials Registry Platform (ICTRP) [Internet]. Geneva: World Health Organization (WHO). 2008 [accessed 02.11.20]. Available from: http://isrctn.com/ISRCTN38082350 |
| Chan 2016  (NCT01943305) | Chan KR, Wang X, Saron WAA, Gan ES, Tan HC, Mok DZL, et al. Cross-reactive antibodies enhance live attenuated virus infection for increased immunogenicity. Nat Microbiol 2016;1:16164. |
|  | Low JG, Wijaya L, Li GK, Lim EY, Shum AK, Cheung YB, et al. The role of pre-existing cross-reactive antibodies in determining the efficacy of vaccination in humans: study protocol for a randomized controlled trial. Trials 2015;16:147. |
| Collaborative Group for Studies of Yellow Fever Vaccine 2015  (ISRCTN72367932) | Collaborative Group for Studies of Yellow Fever Vaccine. A randomised double-blind clinical trial of two yellow fever vaccines prepared with substrains 17DD and 17D-213/77 in children nine-23 months old. Mem Inst Oswaldo Cruz 2015;110(6):771-80. |
|  | Campi-Azevedo AC, de Araujo-Porto LP, Luiza-Silva M, Batista MA, Martins MA, Sathler-Avelar R, et al. 17DD and 17D-213/77 yellow fever substrains trigger a balanced cytokine profile in primary vaccinated children. PLoS One 2012;7(12):e49828. |
|  | Collaborative Group for Studies with Yellow Fever Vaccine. Randomized, double-blind, multicenter study of the immunogenicity and reactogenicity of 17DD and WHO 17D-213/77 yellow fever vaccines in children: implications for the Brazilian National Immunization Program. Vaccine 2007;25(16):3118-23. |
|  | Bio-Manguinhos (Brazil). Multicentre, randomised, double-blind trial comparing yellow fever vaccines from 17D and WHO 17DD-213/77 substrains in children. ISRCTN72367932. In: WHO International Clinical Trials Registry Platform (ICTRP) [Internet]. Geneva: World Health Organization (WHO). 2006 [accessed 02.11.20]. Available from: http://isrctn.com/ISRCTN72367932 |
| Coursaget 1995 | Coursaget P, Fritzell B, Blondeau C, Saliou P, Diop-Mar I. Simultaneous injection of plasma-derived or recombinant hepatitis B vaccines with yellow fever and killed polio vaccines. Vaccine 1995;13(1):109-11. |
| Edupuganti 2013 | Edupuganti S, Eidex RB, Keyserling H, Akondy RS, Lanciotti R, Orenstein W, et al. A randomized, double-blind, controlled trial of the 17D yellow fever virus vaccine given in combination with immune globulin or placebo: comparative viremia and immunogenicity. Am J Trop Med Hyg 2013;88(1):172-7. |
| Guirakhoo 2006 | Guirakhoo F, Kitchener S, Morrison D, Forrat R, McCarthy K, Nichols R, et al. Live attenuated chimeric yellow fever dengue type 2 (ChimeriVax-DEN2) vaccine: phase I clinical trial for safety and immunogenicity: effect of yellow fever pre-immunity in induction of cross neutralizing antibody responses to all 4 dengue serotypes. Hum Vaccin 2006;2(2):60-7. |
| Juan-Giner 2021 | Juan-Giner A, Kimathi D, Grantz KH, Hamaluba M, Kazooba P, Njuguna P, et al. Immunogenicity and safety of fractional doses of yellow fever vaccines: a randomised, double-blind, non-inferiority trial. Lancet 2021;397(10269):119-27. |
| Lang 1999 | Lang J, Zuckerman J, Clarke P, Barrett P, Kirkpatrick C, Blondeau C. Comparison of the immunogenicity and safety of two 17D yellow fever vaccines. Am J Trop Med Hyg 1999;60(6):1045-50. |
| Lopez 2016  (NCT01436396; EudraCT 2014-001714-26) | Lopez P, Lanata CF, Zambrano B, Cortes M, Andrade T, Amemiya I, et al. Immunogenicity and safety of yellow fever vaccine (Stamaril) when administered concomitantly with a tetravalent dengue vaccine candidate in healthy toddlers at 12-13 months of age in Colombia and Peru: a randomized trial. Pediatr Infect Dis J 2016;35(10):1140-7. |
|  | Sanofi Pasteur, a Sanofi Company. Study of yellow fever vaccine administered with tetravalent dengue vaccine in healthy toddlers. In: ClinicalTrials.gov [Internet]. Bethesda (MD): National Library of Medicine (US). 2011-2013 [cited 2020 Nov 2]. Available from: http://clinicaltrials.gov/show/NCT01436396. NLM Identifier: NCT01436396 |
| Monath 2002 | Monath TP, Nichols R, Archambault WT, Moore L, Marchesani R, Tian J, et al. Comparative safety and immunogenicity of two yellow fever 17D vaccines (ARILVAX and YF-VAX) in a phase III multicenter, double-blind clinical trial. Am J Trop Med Hyg 2002;66(5):533-41 |
| Nasveld 2010  (NCT00982137) | Nasveld PE, Marjason J, Bennett S, Aaskov J, Elliott S, McCarthy K, et al. Concomitant or sequential administration of live attenuated Japanese encephalitis chimeric virus vaccine and yellow fever 17D vaccine: randomized double-blind phase II evaluation of safety and immunogenicity. Hum Vaccin 2010;6(11):906-14. |
|  | Sanofi. Study of live attenuated Japanese encephalitis vaccine (ChimeriVax™-JE) and yellow fever vaccine (STAMARIL®). In: ClinicalTrials.gov [Internet]. Bethesda (MD): National Library of Medicine (US). 2004-2007 [cited 2020 Nov 2]. Available from: http://clinicaltrials.gov/show/NCT00982137. NLM Identifier: NCT00982137 |
| Novartis 2011 (EUCTR2011-000475-14-DE; NCT01466387) | Novartis Vaccines and Diagnostics S.r.l. Study to evaluate the safety and immunogenicity of travel vaccines when administered concomitantly with meningococcal ACWY conjugate vaccine in healthy adults. EUCTR2011-000475-14-DE. In: WHO International Clinical Trials Registry Platform (ICTRP) [Internet]. Geneva: World Health Organization (WHO). 2011 [accessed 02.11.20]. Available from: https://www.clinicaltrialsregister.eu/ctr-search/search?query=eudract_number:2011-000475-14 |
| Osei-Kwasi 2001 | Osei-Kwasi M, Dunyo SK, Koram KA, Afari EA, Odoom JK, Nkrumah FK. Antibody response to 17D yellow fever vaccine in Ghanaian infants. Bull World Health Organ 2001;79(11):1056-9. |
| Roukens 2008  (ISRCTN46326316) | Roukens AHE, van Halem K, de Visser AW, Visser LG. Long-term protection after fractional-dose yellow fever vaccination: follow-up study of a randomized, controlled, noninferiority trial. Ann Intern Med 2018;169(11):761-5. |
|  | Roukens AH, Vossen AC, Bredenbeek PJ, van Dissel JT, Visser LG. Intradermally administered yellow fever vaccine at reduced dose induces a protective immune response: a randomized controlled non-inferiority trial. PLoS One 2008;3(4):e1993. |
| Stefano 1999 | Stefano I, Sato HK, Pannuti CS, Omoto TM, Mann G, Freire MS, et al. Recent immunization against measles does not interfere with the sero-response to yellow fever vaccine. Vaccine 1999;17(9-10):1042-6. |
| **Non-randomised comparative studies (12 studies, 18 references)** | |
| Avelino-Silva 2016a | Avelino-Silva VI, Miyaji KT, Hunt PW, Huang Y, Simoes M, Lima SB, et al. CD4/CD8 ratio and KT ratio predict yellow fever vaccine immunogenicity in HIV-infected patients. PLoS Negl Trop Dis 2016;10(12):e0005219. |
| Avelino-Silva 2016b | Avelino-Silva VI, Miyaji KT, Mathias A, Costa DA, de Carvalho Dias JZ, Lima SB, et al. CD4/CD8 ratio predicts yellow fever vaccine-induced antibody titers in virologically suppressed HIV-infected patients. J Acquir Immune Defic Syndr 2016;71(2):189-95. |
| Burkhard 2020 | Burkhard J, Ciurea A, Gabay C, Hasler P, Muller R, Niedrig M, et al. Long-term immunogenicity after yellow fever vaccination in immunosuppressed and healthy individuals. Vaccine 2020;38(19):3610-7. |
| Campi-Azevedo 2016 | Campi-Azevedo AC, Costa-Pereira C, Antonelli LR, Fonseca CT, Teixeira-Carvalho A, Villela-Rezende G, et al. Booster dose after 10 years is recommended following 17DD-YF primary vaccination. Hum Vaccin Immunother 2016;12(2):491-502. |
| Collaborative Group for Studies of Yellow Fever Vaccine 2019a | Collaborative Group for Studies on Yellow Fever Vaccines. Duration of immunity in recipients of two doses of 17DD yellow fever vaccine. Vaccine 2019;37(35):5135. |
| Collaborative Group for Studies of Yellow Fever Vaccine 2019b | Campi-Azevedo AC, Peruhype-Magalhaes V, Coelho-Dos-Reis JG, Antonelli LR, Costa-Pereira C, Speziali E, et al. 17DD yellow fever revaccination and heightened long-term immunity in populations of disease-endemic areas, Brazil. Emerg Infect Dis 2019;25(8):1511-21. |
| de Verdiere 2018  (NCT01426243) | Colin de Verdiere N, Durier C, Samri A, Meiffredy V, Launay O, Matheron S, et al. Immunogenicity and safety of yellow fever vaccine in HIV-1-infected patients. AIDS 2018;32(16):2291-9. |
|  | Durier C, Mercier-Delarue S, De Verdiere NC, Meiffredy V, Matheron S, Samri A, et al. Long-term immune response to yellow fever vaccination in HIV-infected and non-infected adults: ANRS EP46 study. Paper presented at European Congress of Clinical Microbiology and Infectious Diseases; 18-21 Apr 2020; Paris: France. 2020. |
|  | Colin de Verdiere N. Safety and immunogenicity of yellow fever vaccine in HIV+ patients: ANRS EP46 NOVAA. Paper presented at International AIDS Conference, IAS 2015; 19-22 Jul 2015; Vancouver: Canada. 2015. |
| Kernéis 2013 | Kernéis S, Launay O, Ancelle T, Iordache L, Naneix-Laroche V, Mechai F, et al. Safety and immunogenicity of yellow fever 17D vaccine in adults receiving systemic corticosteroid therapy: an observational cohort study. Arthritis Care Res 2013;65(9):1522-8. |
| Michel 2015 | Michel R, Berger F, Ravelonarivo J, Dussart P, Dia M, Nacher M, et al. Observational study on immune response to yellow fever and measles vaccines in 9 to 15-month old children. Is it necessary to wait 4 weeks between two live attenuated vaccines? Vaccine 2015;33(20):2301-6. |
| Project RETRO-CI | Sibailly TS, Wiktor SZ, Tsai TF, Cropp BC, Ekpini ER, Adjorlolo-Johnson G, et al. Poor antibody response to yellow fever vaccination in children infected with human immunodeficiency virus type 1. Pediatr Infect Dis J 1997;16(12):1177-9. |
| Roukens 2011 | Roukens AH, Soonawala D, Joosten SA, de Visser AW, Jiang X, Dirksen K, et al. Elderly subjects have a delayed antibody response and prolonged viraemia following yellow fever vaccination: a prospective controlled cohort study. PLoS One 2011;6(12):e27753. |
|  | Roukens AH, Soonawala D, Joosten SA, Visser AWD, Jiang X, Bredenbeek PJ, et al. Delayed antibody response to yellow fever vaccination in elderly coincides with prolonged viraemia. Paper presented at 12th conference of the International Society of Travel Medicine; 8-12 May 2011; Boston: United States. 2011. |
|  | AHE Roukens and LG Visser. Immunity induced by yellow fever vaccination in the elderly (60 yrs or older) traveller. NTR1040. In: WHO International Clinical Trials Registry Platform (ICTRP) [Internet]. Geneva: World Health Organization (WHO). 2007 [accessed 02.11.20]. Available from: https://trialregister.nl/trial/1011 |
|  | Rosenstein MD, de Visser AW, Visser LG, Roukens AHE. Long-term immunity after a single yellow fever vaccination in travelers vaccinated at 60 years or older: a 10-year follow-up study. J Travel Med 2021;Online ahead of print. |
| Valim 2020 | Valim V, Machado KLLL, Miyamoto ST, Pinto AD, Rocha PCM, Serrano EV, et al. Planned yellow fever primary vaccination is safe and immunogenic in patients with autoimmune diseases: a prospective non-interventional study. Front Immunol 2020;11:1382. |
|  | Valim V, Pinto AD, Rocha PCM, Miyamoto ST, Serrano EV, Duque RH, et al. Safety and efficacy of primary yellow fever vaccination in autoimmune disease: a prospective controlled study. Ann Rheum Dis 2019;78(Suppl 2):2110. |
| **Single arm studies (6 studies, 11 references)** | |
| Campi-Azevedo 2019  (NCT02990182) | Campi-Azevedo AC, Reis LR, Peruhype-Magalhaes V, Coelho-Dos-Reis JG, Antonelli LR, Fonseca CT, et al. Short-lived immunity after 17DD yellow fever single dose indicates that booster vaccination may be required to guarantee protective immunity in children. Front Immunol 2019;10:2192. |
|  | Oswaldo Cruz Foundation. Complementary study of the duration of post-vaccination against yellow fever immunity in children. In: ClinicalTrials.gov [Internet]. Bethesda (MD): National Library of Medicine (US). 2015-2017 [cited 2020 Nov 2]. Available from: http://clinicaltrials.gov/show/NCT02990182. NLM Identifier: NCT02990182 |
| Domingo 2019  (ISRCTN82484612) | Domingo C, Fraissinet J, Ansah PO, Kelly C, Bhat N, Sow SO, et al. Long-term immunity against yellow fever in children vaccinated during infancy: a longitudinal cohort study. Lancet Infect Dis 2019;19(12):1363-70. |
|  | Roy Chowdhury P, Meier C, Laraway H, Tang Y, Hodgson A, Sow SO, et al. Immunogenicity of yellow fever vaccine coadministered with MenAfriVac in healthy infants in Ghana and Mali. Clin Infect Dis 2015;61 Suppl 5:S586-93. |
|  | Serum Institute of India Limited. A phase II, double-blind, randomised, controlled, dose ranging study to evaluate the safety, immunogenicity, dose response and schedule response of a meningococcal A conjugate vaccine administered concomitantly with local expanded program on immunisation (EPI) vaccines in healthy infants. ISRCTN82484612. In: WHO International Clinical Trials Registry Platform (ICTRP) [Internet]. Geneva: World Health Organization (WHO). 2008 [dated accessed 02.11.20]. Available from: http://isrctn.com/ISRCTN82484612 |
| Idoko 2020 | Idoko OT, Domingo C, Tapia MD, Sow SO, Geldmacher C, Saathoff E, et al. Serological protection 5-6 years post vaccination against yellow fever in African infants vaccinated in routine programmes. Front Immunol 2020;11:577751. |
| Jia 2019  (ChCTR1800017024) | Jia Q, Jia C, Liu Y, Yang Y, Qi J, Tong L, et al. Clinical evidence for the immunogenicity and immune persistence of vaccination with yellow fever virus strain 17D in Chinese peacekeepers deployed to Africa. Antiviral Res 2019;162:1-4. |
|  | Jinan Military Region Center for Disease Control and Prevention. Study of immunogenicity and persistence of yellow fever attenuated live vaccine. ChiCTR1800017024. In: WHO International Clinical Trials Registry Platform (ICTRP) [Internet]. Geneva: World Health Organization (WHO). 2018 [accessed 02.11.20]. Available from: http://www.chictr.org.cn/showproj.aspx?proj=28675 |
| Kareko 2020 | Kareko BW, Booty BL, Nix CD, Lyski ZL, Slifka MK, Amanna IJ, et al. Persistence of neutralizing antibody responses among yellow fever virus 17D vaccinees living in a nonendemic setting. J Infect Dis 2020;221(12):2018-25. |
| Veit 2018 | Veit O, Domingo C, Niedrig M, Staehelin C, Sonderegger B, Hequet D, et al. Long-term immune response to yellow fever vaccination in human immunodeficiency virus (HIV)-infected individuals depends on HIV RNA suppression status: implications for vaccination schedule. Clin Infect Dis 2018;66(7):1099-108. |
|  | Veit O, Niedrig M, Chapuis-Taillard C, Cavassini M, Mossdorf E, Schmid P, et al. Immunogenicity and safety of yellow fever vaccination for 102 HIV-infected patients. Clin Infect Dis 2009;48(5):659-66. |
