## Supplementary figures and images for "Duration of protection after vaccination against yellow fever - systematic review and meta-analysis"

### Supplementary Data 4 - PRISMA - Figure 1

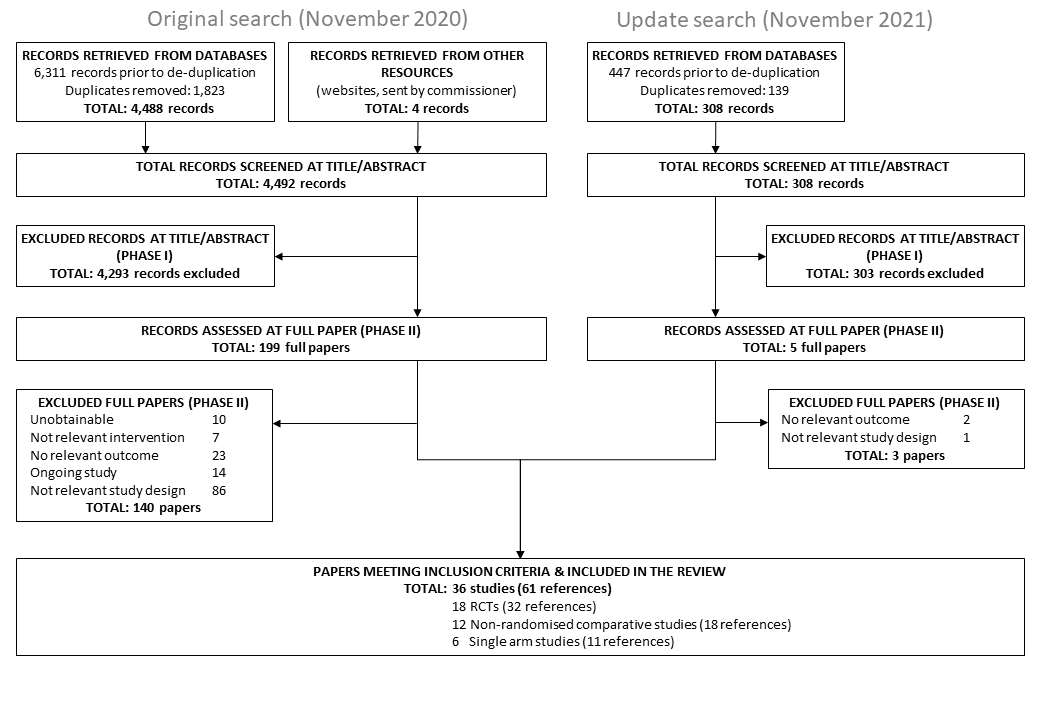
