## Supplementary Data 5 - Risk of bias - Table 5 and 6 for "Duration of protection after vaccination against yellow fever - systematic review and meta-analysis"

**Supplementary Table 5**: **Risk of bias of randomised controlled trials (RCT)**

| **Study** | **Domain 1** | **Domain 2** | **Domain 3** | **Domain 4** | **Domain 5** | **Domain 6** | **Domain 7** | **Domain 8** | **Overall ROB** |
| --- | --- | --- | --- | --- | --- | --- | --- | --- | --- |
| Asante 2020^1^ | low | low | high | high | high | high | low | low | **High** |
| Belmusto-Worn 2005^2^ | unclear | low | low | low | low | high | unclear | low | **Unclear** |
| Camacho 2004^3^ | low | low | low | low | high | low | low | low | **Low** |
| Campi-Azevedo 2014^4^ | unclear | unclear | unclear | unclear | unclear | low | low | low | **Unclear** |
| Chan 2016^5^ | unclear | unclear | high | high | high | low | low | low | **High** |
| Collaborative Group for Studies of Yellow Fever Vaccine 2015^6^ | low | low | low | low | low | low | low | low | **Low** |
| Coursaget 1995^7^ | unclear | unclear | unclear | unclear | unclear | high | unclear | unclear | **High** |
| Edupuganti 2013^8^ | unclear | high | unclear | unclear | high | low | low | unclear | **High** |
| Guirakhoo 2006^9^ | unclear | high | low | unclear | high | low | low | low | **High** |
| Juan-Giner 2021^10^ | low | low | low | high | low | low | low | low | **Low** |
| Lang 1999^11^ | unclear | unclear | low | low | low | high | low | low | **Unclear** |
| Lopez 2016^12^ | low | low | low | low | low | high | low | low | **High** |
| Monath 2002^13^ | unclear | unclear | unclear | unclear | unclear | high | unclear | low | **Unclear** |
| Nasveld 2010^14^ | low | high | high | high | high | high | unclear | low | **High** |
| Novartis 2011^15^ | unclear | unclear | high | high | low | low | low | low | **High** |
| Osei-Kwasi 2001^16^ | unclear | low | low | unclear | high | high | low | low | **High** |
| Roukens 2008^17^ | low | low | unclear | high | unclear | low | unclear | low | **Low** |
| Stefano 1999^18^ | unclear | high | high | high | high | low | low | low | **High** |
| See Supplementary XX for details of ROB tool. Briefly, domain 1 (randomisation), domain 2 (allocation concealment), domain 3 (participant blinding), domain 4 (personnel blinding), domain 5 (assessor blinding), domain 6 (data completeness), domain 7 (selective reporting), domain 8 (other biases).  RCT = randomised controlled trial; ROB = risk of bias | | | | | | | | | |

**Supplementary Table 6: Risk of bias of non-randomised studies (Non-RCT and single arm studies)**

| **Study** | **Domain 1** | **Domain 2** | **Domain 3** | **Domain 4** | **Domain 5** | **Domain 6** | **Domain 7** | **Domain 8** | **Domain 9** | **Overall ROB** |
| --- | --- | --- | --- | --- | --- | --- | --- | --- | --- | --- |
| **Non-randomised comparative studies** | | | | | | | | | | |
| Avelino-Silva 2016a^19^ | yes | unclear | unclear | yes | yes | yes | yes | yes | yes | **Unclear** |
| Avelino-Silva 2016b^20^ | yes | no | unclear | yes | no | unclear | yes | yes | NA | **High** |
| Burkhard 2010^21^ | yes | unclear | unclear | yes | no | yes | yes | yes | yes | **High** |
| Campi-Azevado 2016^22^ | yes | unclear | yes | no | yes | yes | yes | yes | yes | **High** |
| Collaborative Group for Studies of Yellow Fever Vaccine 2019a^23^ | yes | yes | yes | unclear | unclear | yes | yes | yes | unclear | **Unclear** |
| Collaborative Group for Studies of Yellow Fever Vaccine 2019b^24^ | yes | yes | yes | no | yes | yes | yes | yes | yes | **High** |
| De Verdiere 2018^25^ | yes | no | yes | yes | yes | yes | yes | yes | yes | **High** |
| Kernéis 2013^26^ | yes | yes | yes | yes | yes | yes | yes | yes | yes | **Low** |
| Michel 2015^27^ | yes | yes | yes | yes | no | yes | yes | yes | yes | **High** |
| Project RETRO-CI^28^ | yes | NA | NA | no | yes | no | yes | no | NA | **High** |
| Roukens 2011^29^ | yes | unclear | yes | yes | yes | yes | yes | yes | yes | **Unclear** |
| Valim 2020^30^ | yes | yes | yes | yes | yes | yes | yes | yes | yes | **Low** |
| **Single arm studies** | | | | | | | | | | |
| Campi-Azevado 2019^31^ | yes | NA | NA | no | yes | unclear | yes | yes | yes | **High** |
| Domingo 2019^32^ | yes | yes | unclear | no | no | no | yes | yes | yes | **High** |
| Idoko 2020^33^ | yes | yes | NA | no | no | unclear | yes | yes | NA | **High** |
| Jia 2019^34^ | yes | NA | NA | no | yes | yes | yes | yes | yes | **High** |
| Kareko 2018^35^ | yes | NA | NA | no | no | unclear | yes | yes | NA | **High** |
| Veit 2018^36^ | yes | yes | yes | NA | NA | yes | yes | yes | no | **High** |
| See Supplementary XX for details of ROB tool for non-randomised studies. Briefly, domain 1 (clarity of ‘cause’ and ‘effect’), domain 2 (comparison similarities), domain 3 (comparisons of similar treatments), domain 4 (control group), domain 5 (multiple measurements pre and post), domain 6 (follow up), domain 7 (similarity of outcome measures), domain 8 (measurement reliability), domain 9 appropriate statistical analysis).  NA = not applicable; ROB = risk of bias | | | | | | | | | | |
