## Supplementary Data 6 - Forest plots - Figures 001-135 for "Duration of protection after vaccination against yellow fever - systematic review and meta-analysis"

**Supplementary Data - Forest plots - Figures 001-135**

001

| Design / Study | Events | Total | Proportion | 95%-CI | Weight (common) | Weight (random) |
|---|---|---|---|---|---|---|
| **RCT** | | | | | | |
| Camacho 2005 | 744 | 815 | 0.91 | [0.89; 0.93] | 70.0% | 18.5% |
| Campi−Azevedo 2014 | 154 | 157 | 0.98 | [0.95; 1.00] | 3.2% | 7.3% |
| Edupuganti 2013 | 40 | 40 | 1.00 | [0.91; 1.00] | 0.5% | 1.8% |
| Edupuganti 2013 | 40 | 40 | 1.00 | [0.91; 1.00] | 0.5% | 1.8% |
| Guirakhoo 2006 | 13 | 13 | 1.00 | [0.75; 1.00] | 0.5% | 1.7% |
| Guirakhoo 2006 | 13 | 13 | 1.00 | [0.75; 1.00] | 0.5% | 1.7% |
| Guirakhoo 2006 | 12 | 13 | 0.92 | [0.64; 1.00] | 1.0% | 3.1% |
| Juan−Giner 2021 | 116 | 118 | 0.98 | [0.94; 1.00] | 2.1% | 5.6% |
| Juan−Giner 2021 | 118 | 118 | 1.00 | [0.97; 1.00] | 0.5% | 1.8% |
| Juan−Giner 2021 | 116 | 119 | 0.97 | [0.93; 0.99] | 3.2% | 7.3% |
| Juan−Giner 2021 | 120 | 120 | 1.00 | [0.97; 1.00] | 0.5% | 1.8% |
| Lang 1999 | 93 | 93 | 1.00 | [0.96; 1.00] | 0.5% | 1.8% |
| Lang 1999 | 92 | 92 | 1.00 | [0.96; 1.00] | 0.5% | 1.8% |
| Monath 2002 | 279 | 283 | 0.99 | [0.96; 1.00] | 4.3% | 8.7% |
| Monath 2002 | 289 | 291 | 0.99 | [0.98; 1.00] | 2.1% | 5.6% |
| Nasveld 2010 | 17 | 17 | 1.00 | [0.80; 1.00] | 0.5% | 1.7% |
| Nasveld 2010 | 23 | 23 | 1.00 | [0.85; 1.00] | 0.5% | 1.8% |
| Nasveld 2010 | 23 | 23 | 1.00 | [0.85; 1.00] | 0.5% | 1.8% |
| Novartis 2011 | 100 | 100 | 1.00 | [0.96; 1.00] | 0.5% | 1.8% |
| Roukens 2008 | 78 | 78 | 1.00 | [0.95; 1.00] | 0.5% | 1.8% |
| **Common effect model** | | **2566** | **0.94** | **[0.93; 0.95]** | **92.7%** | **--** |
| **Random effects model** | | | **0.98** | **[0.97; 0.99]** | **--** | **78.9%** |
| Heterogeneity: $I^2 = 70\%$, $\tau^2 = 0.2286$, $p < 0.01$ | | | | | | |
| **Non−randomised comparative study** | | | | | | |
| Campi−Azevedo 2016 | 37 | 39 | 0.95 | [0.83; 0.99] | 2.0% | 5.4% |
| Collaborative Group 2019b | 48 | 50 | 0.96 | [0.86; 1.00] | 2.1% | 5.5% |
| de Verdiere 2018 | 30 | 30 | 1.00 | [0.88; 1.00] | 0.5% | 1.8% |
| Roukens 2011 | 30 | 30 | 1.00 | [0.88; 1.00] | 0.5% | 1.8% |
| Roukens 2011 | 28 | 28 | 1.00 | [0.88; 1.00] | 0.5% | 1.8% |
| Valim 2020 | 22 | 23 | 0.96 | [0.78; 1.00] | 1.0% | 3.2% |
| **Common effect model** | | **200** | **0.96** | **[0.93; 0.98]** | **6.7%** | **--** |
| **Random effects model** | | | **0.96** | **[0.93; 0.98]** | **--** | **19.3%** |
| Heterogeneity: $I^2 = 0\%$, $\tau^2 = 0$, $p = 0.94$ | | | | | | |
| **Single arm** | | | | | | |
| Jia 2019 | 349 | 349 | 1.00 | [0.99; 1.00] | 0.5% | 1.8% |
| **Common effect model** | | **349** | **1.00** | **[0.98; 1.00]** | **0.5%** | **--** |
| **Random effects model** | | | **1.00** | **[0.98; 1.00]** | **--** | **1.8%** |
| Heterogeneity: not applicable | | | | | | |
| **Common effect model** | | **3115** | **0.95** | **[0.93; 0.95]** | **100.0%** | **--** |
| **Random effects model** | | | **0.98** | **[0.97; 0.98]** | **--** | **100.0%** |

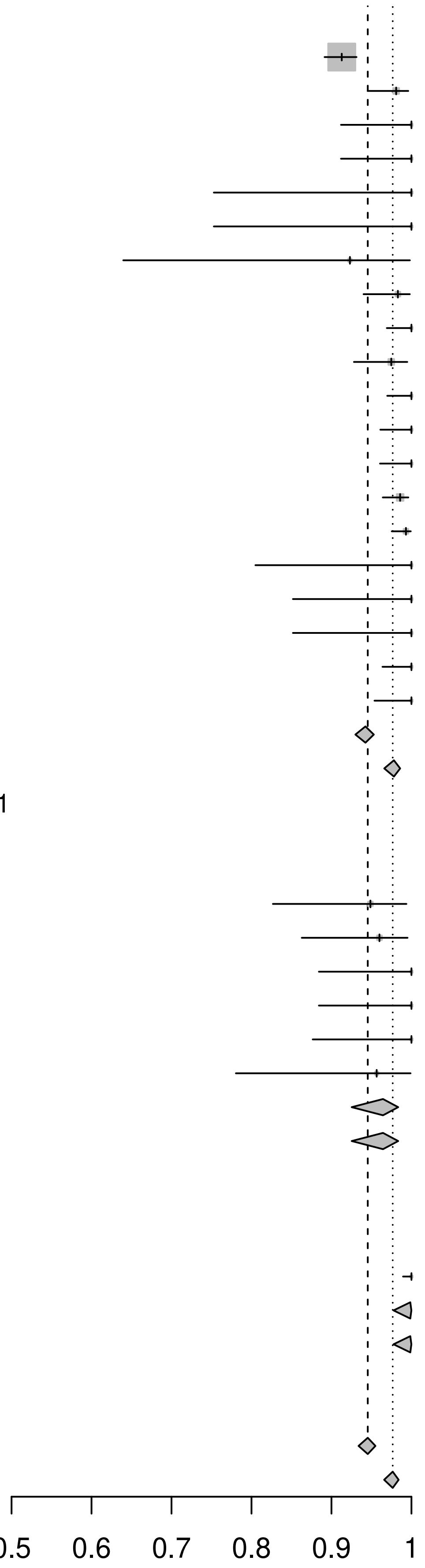

Heterogeneity: $I^2 = 64\%$, $\tau^2 = 0.1966$, $p < 0.01$
Test for subgroup differences (fixed effect): $\chi^2_2 = 8.34$, df = 2 ($p = 0.02$)
Test for subgroup differences (random effects): $\chi^2_2 = 5.21$, df = 2 ($p = 0.07$)

002

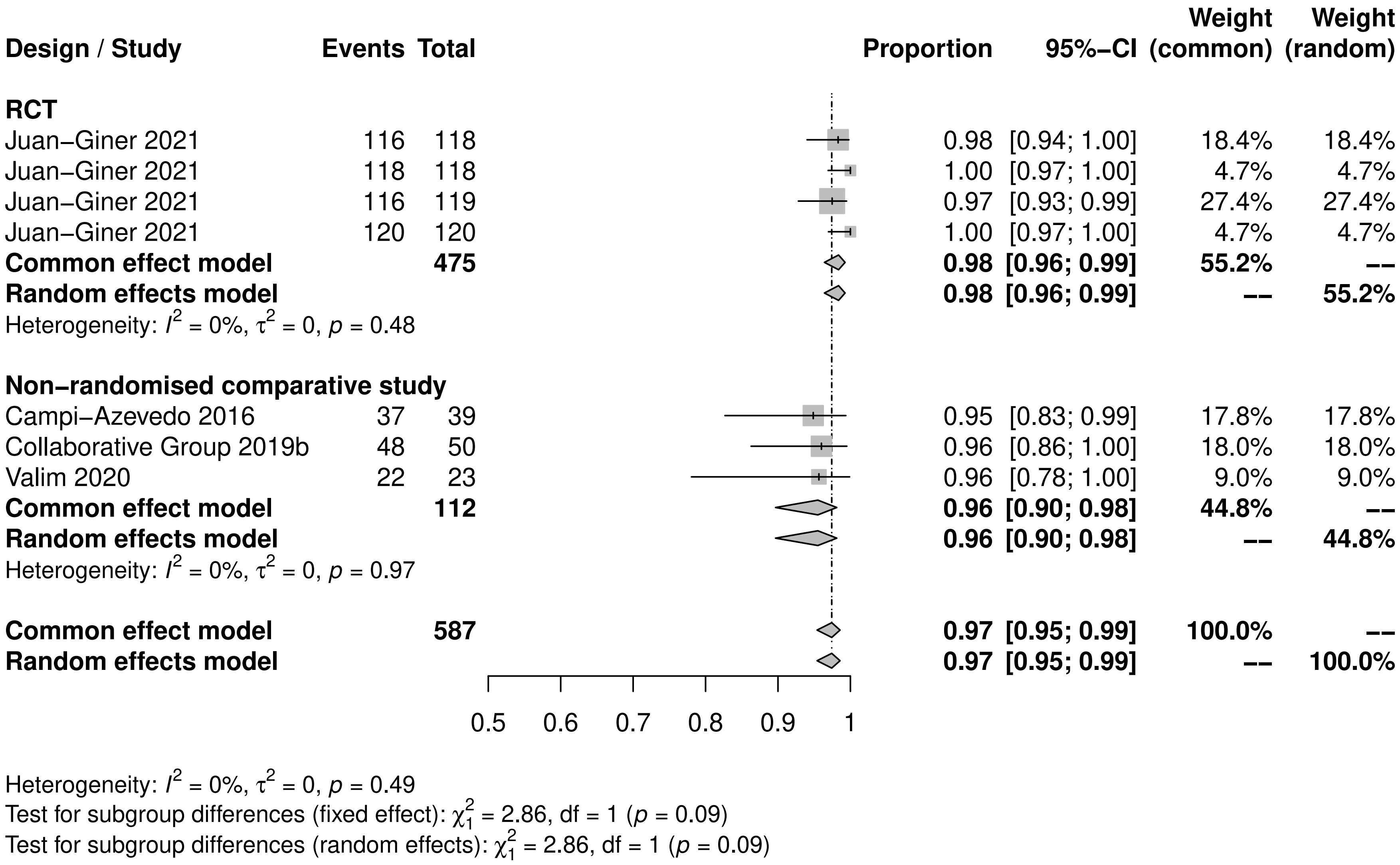

Design / Study Events Total Proportion 95%−CI Weight (common) Weight (random)
RCT
Juan−Giner 2021 116 118 0.98 [0.94; 1.00] 18.4% 18.4%
Juan−Giner 2021 118 118 1.00 [0.97; 1.00] 4.7% 4.7%
Juan−Giner 2021 116 119 0.97 [0.93; 0.99] 27.4% 27.4%
Juan−Giner 2021 120 120 1.00 [0.97; 1.00] 4.7% 4.7%
Common effect model 475 0.98 [0.96; 0.99] 55.2% −−
Random effects model 0.98 [0.96; 0.99] −− 55.2%
Heterogeneity: $I^2 = 0\%$, $\tau^2 = 0$, $p = 0.48$
Non−randomised comparative study
Campi−Azevedo 2016 37 39 0.95 [0.83; 0.99] 17.8% 17.8%
Collaborative Group 2019b 48 50 0.96 [0.86; 1.00] 18.0% 18.0%
Valim 2020 22 23 0.96 [0.78; 1.00] 9.0% 9.0%
Common effect model 112 0.96 [0.90; 0.98] 44.8% −−
Random effects model 0.96 [0.90; 0.98] −− 44.8%
Heterogeneity: $I^2 = 0\%$, $\tau^2 = 0$, $p = 0.97$
Common effect model 587 0.97 [0.95; 0.99] 100.0% −−
Random effects model 0.97 [0.95; 0.99] −− 100.0%
0.5 0.6 0.7 0.8 0.9 1
Heterogeneity: $I^2 = 0\%$, $\tau^2 = 0$, $p = 0.49$
Test for subgroup differences (fixed effect): $\chi^2_1 = 2.86$, df = 1 ($p = 0.09$)
Test for subgroup differences (random effects): $\chi^2_1 = 2.86$, df = 1 ($p = 0.09$)

003

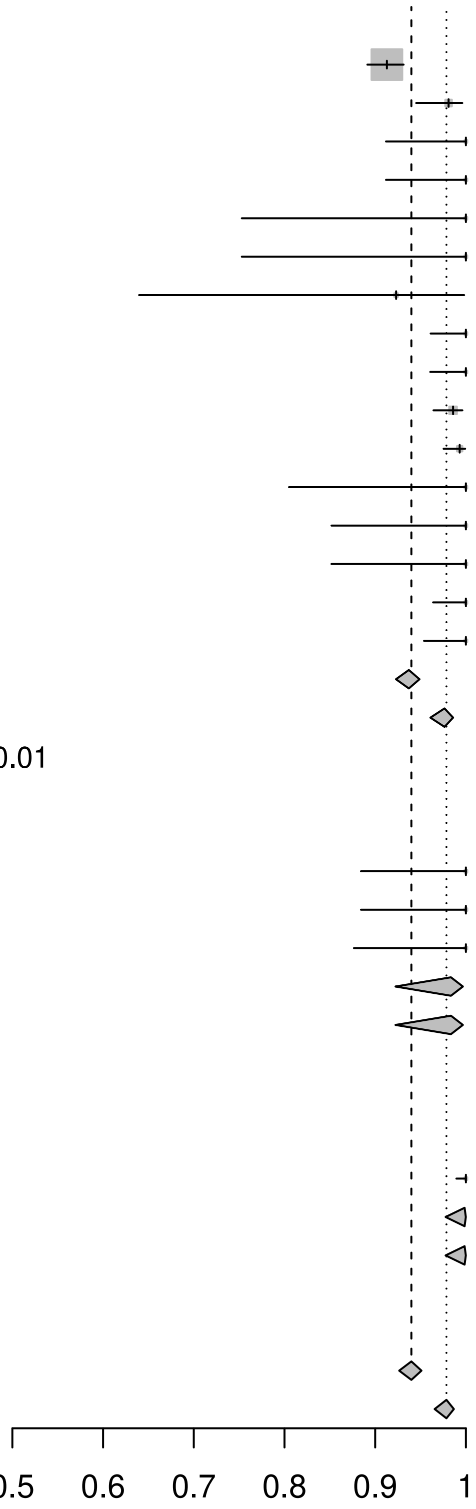

| Design / Study | Events | Total | Proportion | 95%−CI | Weight (common) | Weight (random) |
|---|---|---|---|---|---|---|
| **RCT** | | | | | | |
| Camacho 2005 | 744 | 815 | 0.91 | [0.89; 0.93] | 79.1% | 23.4% |
| Campi−Azevedo 2014 | 154 | 157 | 0.98 | [0.95; 1.00] | 3.6% | 10.5% |
| Edupuganti 2013 | 40 | 40 | 1.00 | [0.91; 1.00] | 0.6% | 2.7% |
| Edupuganti 2013 | 40 | 40 | 1.00 | [0.91; 1.00] | 0.6% | 2.7% |
| Guirakhoo 2006 | 13 | 13 | 1.00 | [0.75; 1.00] | 0.6% | 2.7% |
| Guirakhoo 2006 | 13 | 13 | 1.00 | [0.75; 1.00] | 0.6% | 2.7% |
| Guirakhoo 2006 | 12 | 13 | 0.92 | [0.64; 1.00] | 1.1% | 4.7% |
| Lang 1999 | 93 | 93 | 1.00 | [0.96; 1.00] | 0.6% | 2.7% |
| Lang 1999 | 92 | 92 | 1.00 | [0.96; 1.00] | 0.6% | 2.7% |
| Monath 2002 | 279 | 283 | 0.99 | [0.96; 1.00] | 4.8% | 12.3% |
| Monath 2002 | 289 | 291 | 0.99 | [0.98; 1.00] | 2.4% | 8.2% |
| Nasveld 2010 | 17 | 17 | 1.00 | [0.80; 1.00] | 0.6% | 2.7% |
| Nasveld 2010 | 23 | 23 | 1.00 | [0.85; 1.00] | 0.6% | 2.7% |
| Nasveld 2010 | 23 | 23 | 1.00 | [0.85; 1.00] | 0.6% | 2.7% |
| Novartis 2011 | 100 | 100 | 1.00 | [0.96; 1.00] | 0.6% | 2.7% |
| Roukens 2008 | 78 | 78 | 1.00 | [0.95; 1.00] | 0.6% | 2.7% |
| **Common effect model** | | **2091** | **0.94** | **[0.92; 0.95]** | **97.6%** | **--** |
| **Random effects model** | | | **0.98** | **[0.96; 0.99]** | **--** | **89.1%** |
| Heterogeneity: $I^2 = 70\%$, $\tau^2 = 0.2631$, $p < 0.01$ | | | | | | |
| **Non−randomised comparative study** | | | | | | |
| de Verdiere 2018 | 30 | 30 | 1.00 | [0.88; 1.00] | 0.6% | 2.7% |
| Roukens 2011 | 30 | 30 | 1.00 | [0.88; 1.00] | 0.6% | 2.7% |
| Roukens 2011 | 28 | 28 | 1.00 | [0.88; 1.00] | 0.6% | 2.7% |
| **Common effect model** | | **88** | **0.98** | **[0.92; 1.00]** | **1.8%** | **--** |
| **Random effects model** | | | **0.98** | **[0.92; 1.00]** | **--** | **8.2%** |
| Heterogeneity: $I^2 = 0\%$, $\tau^2 = 0$, $p = 1.00$ | | | | | | |
| **Single arm** | | | | | | |
| Jia 2019 | 349 | 349 | 1.00 | [0.99; 1.00] | 0.6% | 2.8% |
| **Common effect model** | | **349** | **1.00** | **[0.98; 1.00]** | **0.6%** | **--** |
| **Random effects model** | | | **1.00** | **[0.98; 1.00]** | **--** | **2.8%** |
| Heterogeneity: not applicable | | | | | | |
| **Common effect model** | | **2528** | **0.94** | **[0.93; 0.95]** | **100.0%** | **--** |
| **Random effects model** | | | **0.98** | **[0.97; 0.99]** | **--** | **100.0%** |

Heterogeneity: $I^2 = 68\%$, $\tau^2 = 0.2498$, $p < 0.01$
Test for subgroup differences (fixed effect): $\chi^2_2 = 10.04$, df = 2 ($p < 0.01$)
Test for subgroup differences (random effects): $\chi^2_2 = 3.95$, df = 2 ($p = 0.14$)

004

| Design / Study | Events | Total | | Proportion | 95%-CI | Weight (common) | Weight (random) |
|---|---|---|---|---|---|---|---|
| **RCT** | | | | | | | |
| Asante 2020 | 202 | 206 | | 0.98 | [0.95; 0.99] | 0.8% | 4.7% |
| Asante 2020 | 213 | 216 | | 0.99 | [0.96; 1.00] | 0.6% | 4.5% |
| Belmusto−Worn 2005 | 619 | 652 | | 0.95 | [0.93; 0.96] | 6.1% | 5.5% |
| Belmusto−Worn 2005 | 298 | 329 | | 0.91 | [0.87; 0.94] | 5.5% | 5.4% |
| Collaborative Group 2015 | 803 | 981 | | 0.82 | [0.79; 0.84] | 28.4% | 5.6% |
| Collaborative Group 2015 | 820 | 985 | | 0.83 | [0.81; 0.86] | 26.8% | 5.6% |
| Coursaget 1995 | 53 | 55 | | 0.96 | [0.87; 1.00] | 0.4% | 4.0% |
| Coursaget 1995 | 53 | 55 | | 0.96 | [0.87; 1.00] | 0.4% | 4.0% |
| Coursaget 1995 | 59 | 62 | | 0.95 | [0.87; 0.99] | 0.6% | 4.4% |
| Lopez 2016 | 373 | 378 | | 0.99 | [0.97; 1.00] | 1.0% | 4.9% |
| Lopez 2016 | 375 | 376 | | 1.00 | [0.99; 1.00] | 0.2% | 3.2% |
| Osei−Kwasi 2001 | 137 | 139 | | 0.99 | [0.95; 1.00] | 0.4% | 4.1% |
| Osei−Kwasi 2001 | 147 | 150 | | 0.98 | [0.94; 1.00] | 0.6% | 4.5% |
| **Common effect model** | | **4584** | | **0.87** | **[0.86; 0.88]** | **71.6%** | **--** |
| **Random effects model** | | | | **0.96** | **[0.93; 0.98]** | **--** | **60.4%** |
| Heterogeneity: $I^2 = 93\%$, $\tau^2 = 1.1445$, $p < 0.01$ | | | | | | | |
| **Non−randomised comparative study** | | | | | | | |
| Michel 2015 | 49 | 54 | | 0.91 | [0.80; 0.97] | 0.9% | 4.8% |
| Michel 2015 | 209 | 225 | | 0.93 | [0.89; 0.96] | 2.9% | 5.3% |
| **Common effect model** | | **279** | | **0.92** | **[0.89; 0.95]** | **3.8%** | **--** |
| **Random effects model** | | | | **0.92** | **[0.89; 0.95]** | **--** | **10.1%** |
| Heterogeneity: $I^2 = 0\%$, $\tau^2 = 0$, $p = 0.59$ | | | | | | | |
| **Single arm** | | | | | | | |
| Campi−Azevedo 2019 | 40 | 47 | | 0.85 | [0.72; 0.94] | 1.2% | 5.0% |
| Domingo 2019 | 121 | 162 | | 0.75 | [0.67; 0.81] | 6.0% | 5.5% |
| Domingo 2019 | 120 | 168 | | 0.71 | [0.64; 0.78] | 6.7% | 5.5% |
| Domingo 2019 | 58 | 60 | | 0.97 | [0.88; 1.00] | 0.4% | 4.0% |
| Domingo 2019 | 58 | 60 | | 0.97 | [0.88; 1.00] | 0.4% | 4.0% |
| Stefano 1999 | 228 | 294 | | 0.78 | [0.72; 0.82] | 10.0% | 5.5% |
| **Common effect model** | | **791** | | **0.77** | **[0.74; 0.80]** | **24.6%** | **--** |
| **Random effects model** | | | | **0.86** | **[0.72; 0.93]** | **--** | **29.5%** |
| Heterogeneity: $I^2 = 78\%$, $\tau^2 = 0.9485$, $p < 0.01$ | | | | | | | |
| **Common effect model** | | **5654** | | **0.85** | **[0.84; 0.86]** | **100.0%** | **--** |
| **Random effects model** | | | | **0.94** | **[0.90; 0.96]** | **--** | **100.0%** |

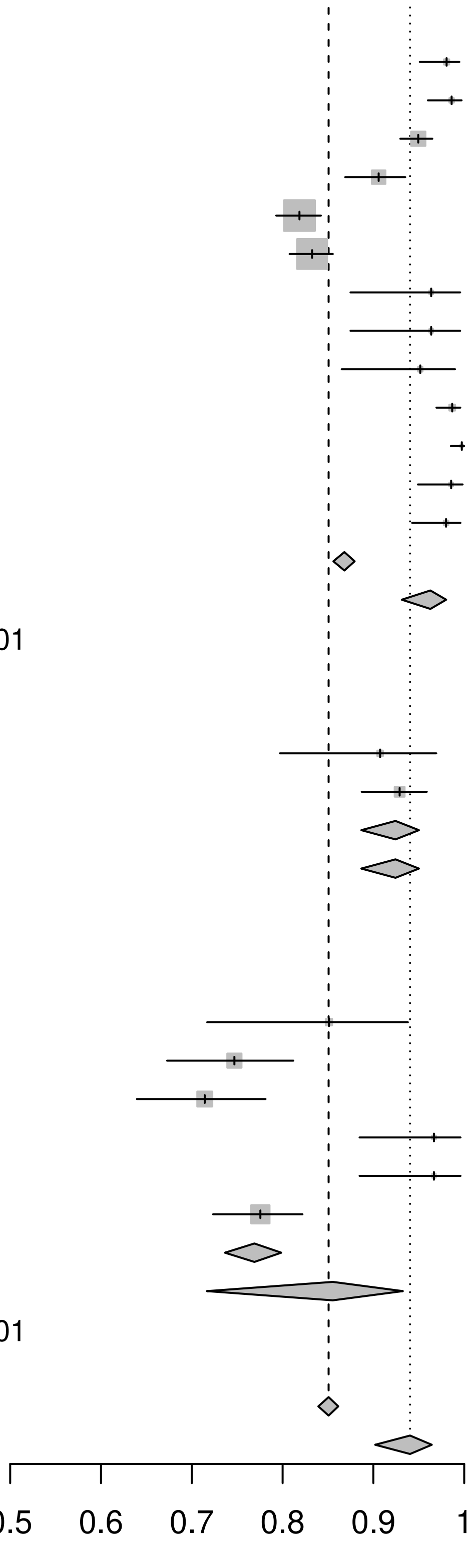

Heterogeneity: $I^2 = 92\%$, $\tau^2 = 1.3502$, $p < 0.01$
Test for subgroup differences (fixed effect): $\chi^2_2 = 55.26$, df = 2 ($p < 0.01$)
Test for subgroup differences (random effects): $\chi^2_2 = 7.71$, df = 2 ($p = 0.02$)

005

| Design / Study | Events | Total | Proportion | 95%-CI | Weight (common) | Weight (random) |
|---|---|---|---|---|---|---|
| **RCT** | | | | | | |
| Asante 2020 | 202 | 206 | 0.98 | [0.95; 0.99] | 0.8% | 4.7% |
| Asante 2020 | 213 | 216 | 0.99 | [0.96; 1.00] | 0.6% | 4.5% |
| Belmusto−Worn 2005 | 619 | 652 | 0.95 | [0.93; 0.96] | 6.1% | 5.5% |
| Belmusto−Worn 2005 | 298 | 329 | 0.91 | [0.87; 0.94] | 5.5% | 5.4% |
| Collaborative Group 2015 | 803 | 981 | 0.82 | [0.79; 0.84] | 28.4% | 5.6% |
| Collaborative Group 2015 | 820 | 985 | 0.83 | [0.81; 0.86] | 26.8% | 5.6% |
| Coursaget 1995 | 53 | 55 | 0.96 | [0.87; 1.00] | 0.4% | 4.0% |
| Coursaget 1995 | 53 | 55 | 0.96 | [0.87; 1.00] | 0.4% | 4.0% |
| Coursaget 1995 | 59 | 62 | 0.95 | [0.87; 0.99] | 0.6% | 4.4% |
| Lopez 2016 | 373 | 378 | 0.99 | [0.97; 1.00] | 1.0% | 4.9% |
| Lopez 2016 | 375 | 376 | 1.00 | [0.99; 1.00] | 0.2% | 3.2% |
| Osei−Kwasi 2001 | 137 | 139 | 0.99 | [0.95; 1.00] | 0.4% | 4.1% |
| Osei−Kwasi 2001 | 147 | 150 | 0.98 | [0.94; 1.00] | 0.6% | 4.5% |
| **Common effect model** | | **4584** | **0.87** | **[0.86; 0.88]** | **71.6%** | **--** |
| **Random effects model** | | | **0.96** | **[0.93; 0.98]** | **--** | **60.4%** |
| Heterogeneity: $I^2 = 93\%$, $\tau^2 = 1.1445$, $p < 0.01$ | | | | | | |
| **Non−randomised comparative study** | | | | | | |
| Michel 2015 | 49 | 54 | 0.91 | [0.80; 0.97] | 0.9% | 4.8% |
| Michel 2015 | 209 | 225 | 0.93 | [0.89; 0.96] | 2.9% | 5.3% |
| **Common effect model** | | **279** | **0.92** | **[0.89; 0.95]** | **3.8%** | **--** |
| **Random effects model** | | | **0.92** | **[0.89; 0.95]** | **--** | **10.1%** |
| Heterogeneity: $I^2 = 0\%$, $\tau^2 = 0$, $p = 0.59$ | | | | | | |
| **Single arm** | | | | | | |
| Campi−Azevedo 2019 | 40 | 47 | 0.85 | [0.72; 0.94] | 1.2% | 5.0% |
| Domingo 2019 | 121 | 162 | 0.75 | [0.67; 0.81] | 6.0% | 5.5% |
| Domingo 2019 | 120 | 168 | 0.71 | [0.64; 0.78] | 6.7% | 5.5% |
| Domingo 2019 | 58 | 60 | 0.97 | [0.88; 1.00] | 0.4% | 4.0% |
| Domingo 2019 | 58 | 60 | 0.97 | [0.88; 1.00] | 0.4% | 4.0% |
| Stefano 1999 | 228 | 294 | 0.78 | [0.72; 0.82] | 10.0% | 5.5% |
| **Common effect model** | | **791** | **0.77** | **[0.74; 0.80]** | **24.6%** | **--** |
| **Random effects model** | | | **0.86** | **[0.72; 0.93]** | **--** | **29.5%** |
| Heterogeneity: $I^2 = 78\%$, $\tau^2 = 0.9485$, $p < 0.01$ | | | | | | |
| **Common effect model** | | **5654** | **0.85** | **[0.84; 0.86]** | **100.0%** | **--** |
| **Random effects model** | | | **0.94** | **[0.90; 0.96]** | **--** | **100.0%** |

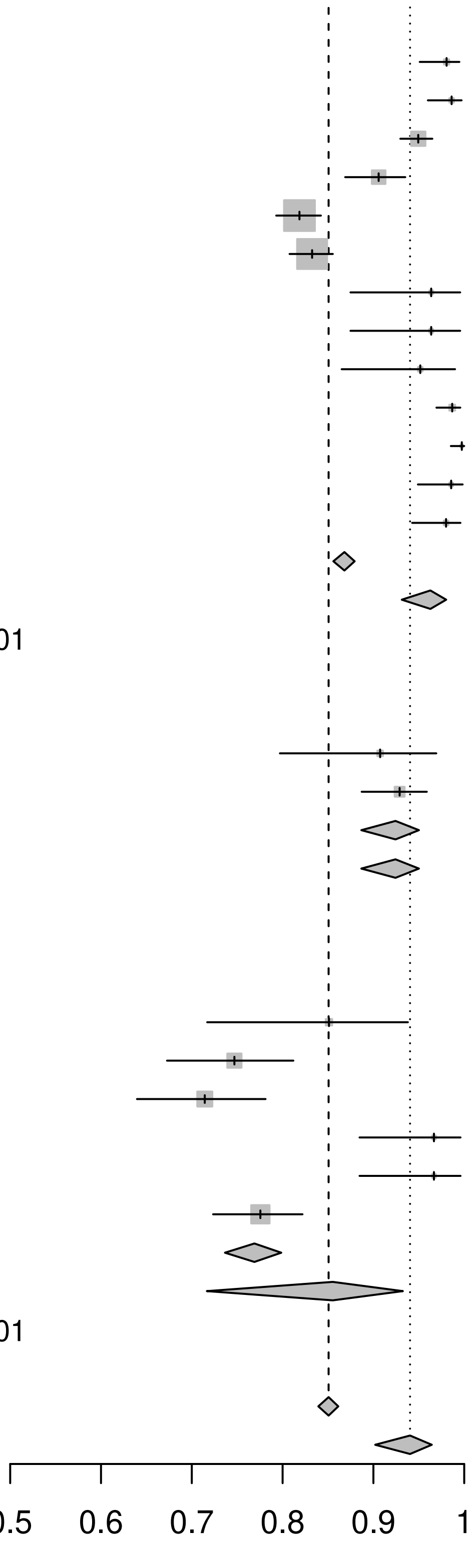

Heterogeneity: $I^2 = 92\%$, $\tau^2 = 1.3502$, $p < 0.01$
Test for subgroup differences (fixed effect): $\chi^2_2 = 55.26$, df = 2 ($p < 0.01$)
Test for subgroup differences (random effects): $\chi^2_2 = 7.71$, df = 2 ($p = 0.02$)

007

| Design / Study | Events | Total | Proportion | 95%−CI | Weight (common) | Weight (random) |
|---|---|---|---|---|---|---|
| **Non−randomised comparative study** | | | | | | |
| de Verdiere 2018 | 39 | 39 | 1.00 | [0.91; 1.00] | 1.7% | 23.5% |
| Kerneis 2013 | 9 | 9 | 1.00 | [0.66; 1.00] | 1.7% | 22.9% |
| Valim 2020 | 125 | 160 | 0.78 | [0.71; 0.84] | 96.6% | 53.6% |
| **Common effect model** | | **208** | **0.79** | **[0.73; 0.85]** | **100.0%** | **--** |
| **Random effects model** | | | **0.92** | **[0.65; 0.98]** | **--** | **100.0%** |
| Heterogeneity: $I^2$ = 66%, $\tau^2$ = 1.5101, $p$ = 0.05 | | | | | | |
| **Common effect model** | | **208** | **0.79** | **[0.73; 0.85]** | **100.0%** | **--** |
| **Random effects model** | | | **0.92** | **[0.65; 0.98]** | **--** | **100.0%** |

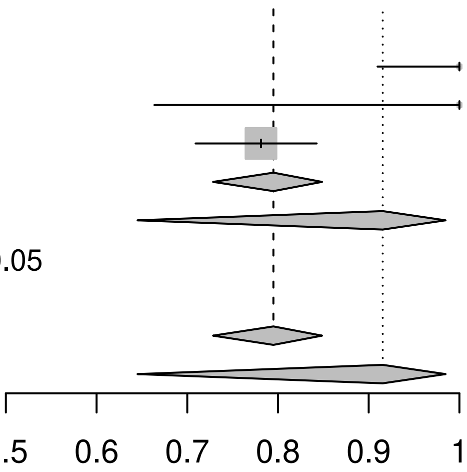

Heterogeneity: $I^2$ = 66%, $\tau^2$ = 1.5101, $p$ = 0.05
Test for subgroup differences (fixed effect): $\chi^2_0$ = 0.00, df = 0 ($p$ = NA)
Test for subgroup differences (random effects): $\chi^2_0$ = 0.00, df = 0 ($p$ = NA)

008

| Design / Study | Events | Total | | Proportion | 95%-CI |
|---|---|---|---|---|---|
| **Non-randomised comparative study** | | | | | |
| Valim 2020 | 125 | 160 | 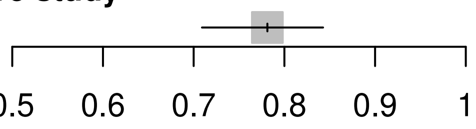 | 0.78 | [0.71; 0.84] |

009

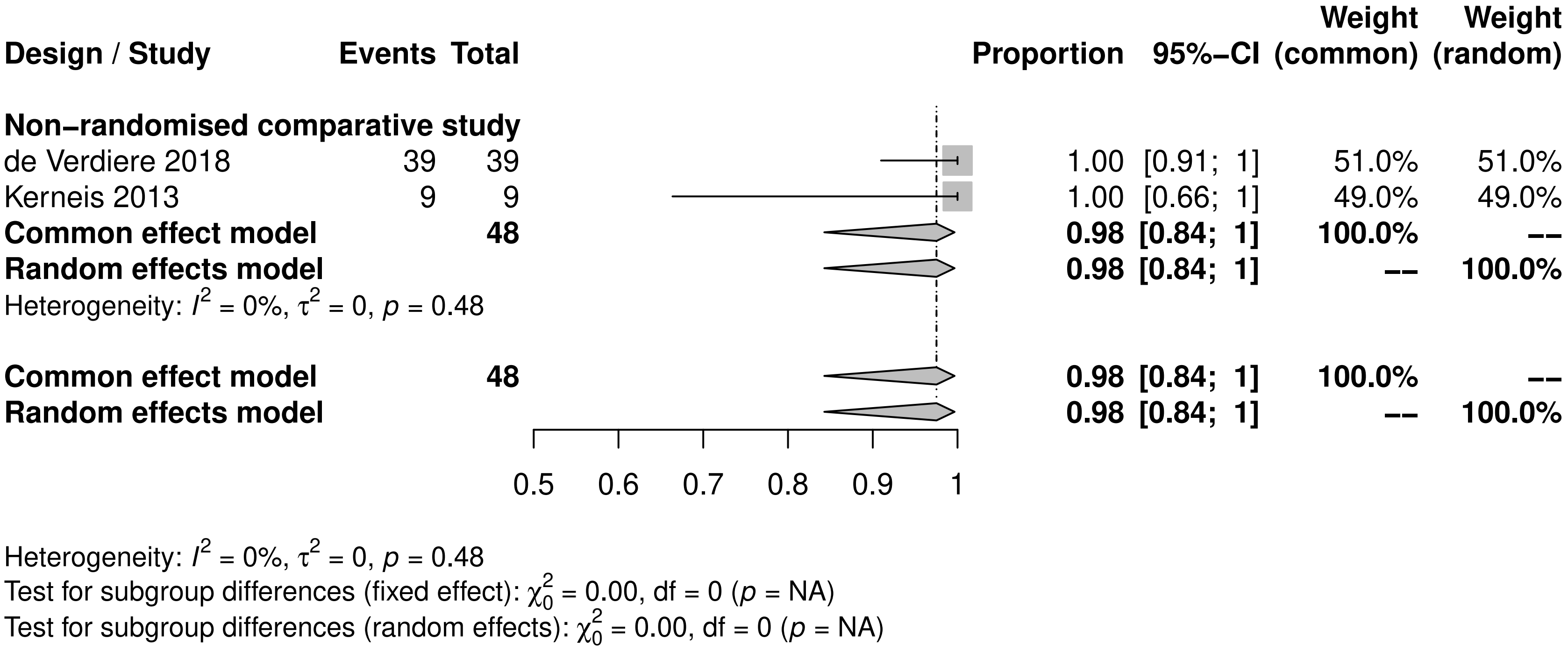
Design / Study
Events
Total
Proportion
95%-CI
Weight (common)
Weight (random)
Non-randomised comparative study
de Verdiere 2018 39 39 1.00 [0.91; 1] 51.0% 51.0%
Kerneis 2013 9 9 1.00 [0.66; 1] 49.0% 49.0%
Common effect model 48 0.98 [0.84; 1] 100.0% --
Random effects model 0.98 [0.84; 1] -- 100.0%
Heterogeneity: $I^2 = 0\%$, $\tau^2 = 0$, $p = 0.48$
Common effect model 48 0.98 [0.84; 1] 100.0% --
Random effects model 0.98 [0.84; 1] -- 100.0%
0.5
0.6
0.7
0.8
0.9
1
Heterogeneity: $I^2 = 0\%$, $\tau^2 = 0$, $p = 0.48$
Test for subgroup differences (fixed effect): $\chi^2_0 = 0.00$, df = 0 ($p$ = NA)
Test for subgroup differences (random effects): $\chi^2_0 = 0.00$, df = 0 ($p$ = NA)

010

| Design / Study | Events | Total | Proportion | 95%–CI | Weight (common) | Weight (random) |
|---|---|---|---|---|---|---|
| **RCT** | | | | | | |
| Campi–Azevedo 2014 | 152 | 157 | 0.97 | [0.93; 0.99] | 31.4% | 31.4% |
| Guirakhoo 2006 | 12 | 13 | 0.92 | [0.64; 1.00] | 6.0% | 6.0% |
| Juan–Giner 2021 | 110 | 111 | 0.99 | [0.95; 1.00] | 6.4% | 6.4% |
| Juan–Giner 2021 | 112 | 112 | 1.00 | [0.97; 1.00] | 3.2% | 3.2% |
| Juan–Giner 2021 | 105 | 106 | 0.99 | [0.95; 1.00] | 6.4% | 6.4% |
| Juan–Giner 2021 | 109 | 109 | 1.00 | [0.97; 1.00] | 3.2% | 3.2% |
| Nasveld 2010 | 17 | 17 | 1.00 | [0.80; 1.00] | 3.2% | 3.2% |
| Nasveld 2010 | 23 | 23 | 1.00 | [0.85; 1.00] | 3.2% | 3.2% |
| Nasveld 2010 | 23 | 23 | 1.00 | [0.85; 1.00] | 3.2% | 3.2% |
| **Common effect model** | | **671** | **0.98** | **[0.96; 0.99]** | **66.2%** | **--** |
| **Random effects model** | | | **0.98** | **[0.96; 0.99]** | **--** | **66.2%** |
| Heterogeneity: $I^2 = 0\%$, $\tau^2 = 0$, $p = 0.60$ | | | | | | |
| **Non–randomised comparative study** | | | | | | |
| Campi–Azevedo 2016 | 34 | 36 | 0.94 | [0.81; 0.99] | 12.3% | 12.3% |
| Collaborative Group 2019b | 38 | 40 | 0.95 | [0.83; 0.99] | 12.3% | 12.3% |
| de Verdiere 2018 | 30 | 30 | 1.00 | [0.88; 1.00] | 3.2% | 3.2% |
| **Common effect model** | | **106** | **0.95** | **[0.89; 0.98]** | **27.8%** | **--** |
| **Random effects model** | | | **0.95** | **[0.89; 0.98]** | **--** | **27.8%** |
| Heterogeneity: $I^2 = 0\%$, $\tau^2 = 0$, $p = 0.72$ | | | | | | |
| **Single arm** | | | | | | |
| Kareko 2020 | 12 | 13 | 0.92 | [0.64; 1.00] | 6.0% | 6.0% |
| **Common effect model** | | **13** | **0.92** | **[0.61; 0.99]** | **6.0%** | **--** |
| **Random effects model** | | | **0.92** | **[0.61; 0.99]** | **--** | **6.0%** |
| Heterogeneity: not applicable | | | | | | |
| **Common effect model** | | **790** | **0.97** | **[0.95; 0.98]** | **100.0%** | **--** |
| **Random effects model** | | | **0.97** | **[0.95; 0.98]** | **--** | **100.0%** |

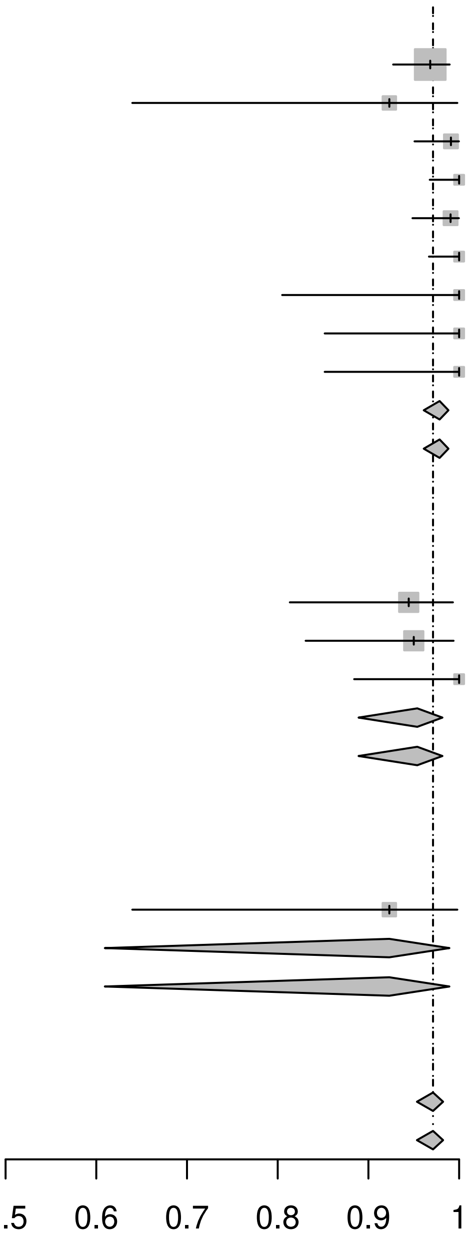

Heterogeneity: $I^2 = 0\%$, $\tau^2 = 0$, $p = 0.61$
Test for subgroup differences (fixed effect): $\chi^2_2 = 2.93$, df = 2 ($p = 0.23$)
Test for subgroup differences (random effects): $\chi^2_2 = 2.93$, df = 2 ($p = 0.23$)

011

| Design / Study | Events | Total | Proportion | 95%-CI | Weight (common) | Weight (random) |
|---|---|---|---|---|---|---|
| **RCT** | | | | | | |
| Juan–Giner 2021 | 110 | 111 | 0.99 | [0.95; 1.00] | 14.6% | 16.1% |
| Juan–Giner 2021 | 112 | 112 | 1.00 | [0.97; 1.00] | 7.4% | 9.4% |
| Juan–Giner 2021 | 105 | 106 | 0.99 | [0.95; 1.00] | 14.6% | 16.1% |
| Juan–Giner 2021 | 109 | 109 | 1.00 | [0.97; 1.00] | 7.4% | 9.4% |
| **Common effect model** | | **438** | **0.99** | **[0.98; 1.00]** | **44.0%** | **--** |
| **Random effects model** | | | **0.99** | **[0.98; 1.00]** | **--** | **50.9%** |
| Heterogeneity: $I^2 = 0\%$, $\tau^2 = 0$, $p = 0.95$ | | | | | | |
| **Non–randomised comparative study** | | | | | | |
| Campi–Azevedo 2016 | 34 | 36 | 0.94 | [0.81; 0.99] | 27.9% | 24.5% |
| Collaborative Group 2019b | 38 | 40 | 0.95 | [0.83; 0.99] | 28.1% | 24.6% |
| **Common effect model** | | **76** | **0.95** | **[0.87; 0.98]** | **56.0%** | **--** |
| **Random effects model** | | | **0.95** | **[0.87; 0.98]** | **--** | **49.1%** |
| Heterogeneity: $I^2 = 0\%$, $\tau^2 = 0$, $p = 0.91$ | | | | | | |
| **Common effect model** | | **514** | **0.98** | **[0.95; 0.99]** | **100.0%** | **--** |
| **Random effects model** | | | **0.98** | **[0.95; 0.99]** | **--** | **100.0%** |

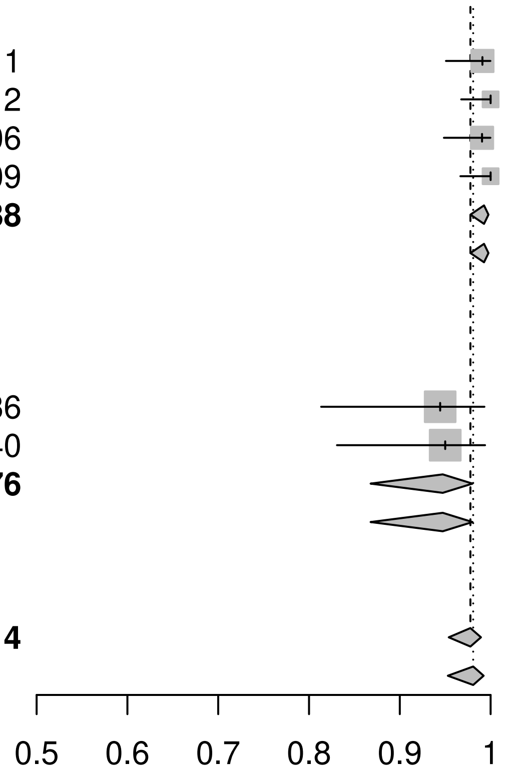

Heterogeneity: $I^2 = 31\%$, $\tau^2 = 0.3837$, $p = 0.20$
Test for subgroup differences (fixed effect): $\chi^2_1 = 6.88$, df = 1 ($p < 0.01$)
Test for subgroup differences (random effects): $\chi^2_1 = 6.88$, df = 1 ($p < 0.01$)

012

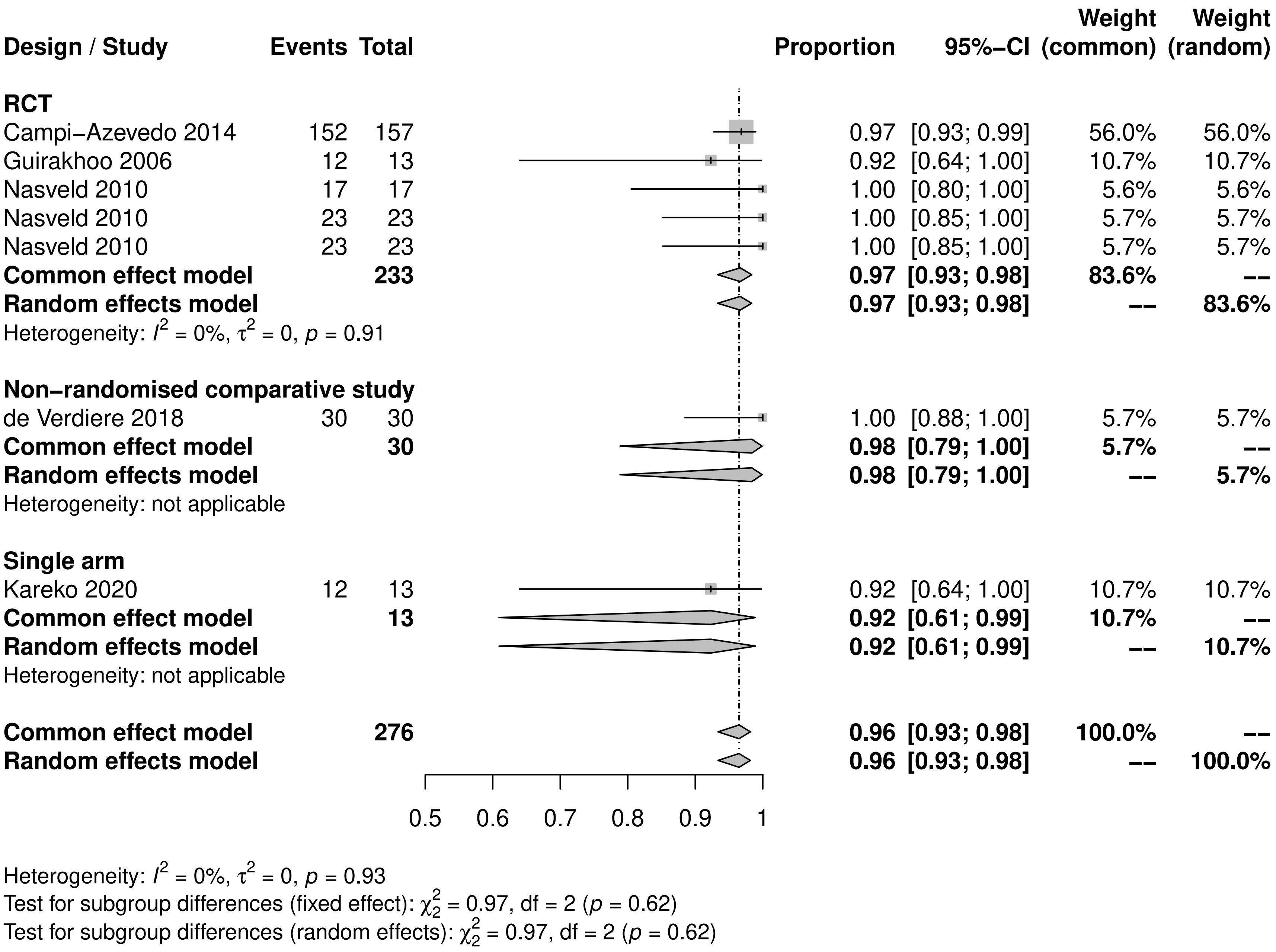

Design / Study
Events
Total
Proportion
95%−CI
Weight (common)
Weight (random)
RCT
Campi−Azevedo 2014 152 157 0.97 [0.93; 0.99] 56.0% 56.0%
Guirakhoo 2006 12 13 0.92 [0.64; 1.00] 10.7% 10.7%
Nasveld 2010 17 17 1.00 [0.80; 1.00] 5.6% 5.6%
Nasveld 2010 23 23 1.00 [0.85; 1.00] 5.7% 5.7%
Nasveld 2010 23 23 1.00 [0.85; 1.00] 5.7% 5.7%
Common effect model 233 0.97 [0.93; 0.98] 83.6% --
Random effects model 0.97 [0.93; 0.98] -- 83.6%
Heterogeneity: $I^2$ = 0%, $\tau^2$ = 0, $p$ = 0.91
Non−randomised comparative study
de Verdiere 2018 30 30 1.00 [0.88; 1.00] 5.7% 5.7%
Common effect model 30 0.98 [0.79; 1.00] 5.7% --
Random effects model 0.98 [0.79; 1.00] -- 5.7%
Heterogeneity: not applicable
Single arm
Kareko 2020 12 13 0.92 [0.64; 1.00] 10.7% 10.7%
Common effect model 13 0.92 [0.61; 0.99] 10.7% --
Random effects model 0.92 [0.61; 0.99] -- 10.7%
Heterogeneity: not applicable
Common effect model 276 0.96 [0.93; 0.98] 100.0% --
Random effects model 0.96 [0.93; 0.98] -- 100.0%
0.5
0.6
0.7
0.8
0.9
1
Heterogeneity: $I^2$ = 0%, $\tau^2$ = 0, $p$ = 0.93
Test for subgroup differences (fixed effect): $\chi^2_2$ = 0.97, df = 2 ($p$ = 0.62)
Test for subgroup differences (random effects): $\chi^2_2$ = 0.97, df = 2 ($p$ = 0.62)

013

| Design / Study | Events | Total | Proportion | 95%-CI | Weight (common) | Weight (random) |
|---|---|---|---|---|---|---|
| **Non–randomised comparative study** | | | | | | |
| Project RETRO–CI | 42 | 57 | 0.74 | [0.60; 0.84] | 4.0% | 23.1% |
| **Common effect model** | | **57** | **0.74** | **[0.61; 0.83]** | **4.0%** | **--** |
| **Random effects model** | | | **0.74** | **[0.61; 0.83]** | **--** | **23.1%** |
| Heterogeneity: not applicable | | | | | | |
| **Single arm** | | | | | | |
| Campi–Azevedo 2019 | 76 | 128 | 0.59 | [0.50; 0.68] | 11.2% | 25.0% |
| Domingo 2019 | 296 | 587 | 0.50 | [0.46; 0.55] | 53.1% | 26.0% |
| Domingo 2019 | 121 | 436 | 0.28 | [0.24; 0.32] | 31.7% | 25.8% |
| **Common effect model** | | **1151** | **0.43** | **[0.41; 0.46]** | **96.0%** | **--** |
| **Random effects model** | | | **0.45** | **[0.27; 0.64]** | **--** | **76.9%** |
| Heterogeneity: $I^2 = 97\%$, $\tau^2 = 0.4594$, $p < 0.01$ | | | | | | |
| **Common effect model** | | **1208** | **0.45** | **[0.42; 0.48]** | **100.0%** | **--** |
| **Random effects model** | | | **0.52** | **[0.33; 0.71]** | **--** | **100.0%** |

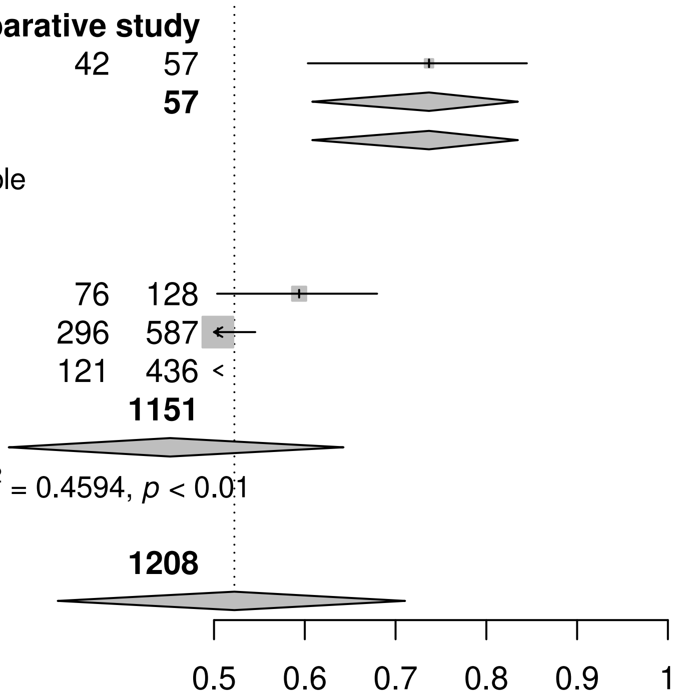

Heterogeneity: $I^2 = 96\%$, $\tau^2 = 0.6454$, $p < 0.01$
Test for subgroup differences (fixed effect): $\chi^2_1 = 17.70$, df = 1 ($p < 0.01$)
Test for subgroup differences (random effects): $\chi^2_1 = 6.01$, df = 1 ($p = 0.01$)

014

| Design / Study | Events | Total | Proportion | 95%-CI | Weight (common) | Weight (random) |
|---|---|---|---|---|---|---|
| **Non-randomised comparative study** | | | | | | |
| Project RETRO-CI | 42 | 57 | 0.74 | [0.60; 0.84] | 4.0% | 23.1% |
| **Common effect model** | | **57** | **0.74** | **[0.61; 0.83]** | **4.0%** | **--** |
| **Random effects model** | | | **0.74** | **[0.61; 0.83]** | **--** | **23.1%** |
| Heterogeneity: not applicable | | | | | | |
| **Single arm** | | | | | | |
| Campi-Azevedo 2019 | 76 | 128 | 0.59 | [0.50; 0.68] | 11.2% | 25.0% |
| Domingo 2019 | 296 | 587 | 0.50 | [0.46; 0.55] | 53.1% | 26.0% |
| Domingo 2019 | 121 | 436 | 0.28 | [0.24; 0.32] | 31.7% | 25.8% |
| **Common effect model** | | **1151** | **0.43** | **[0.41; 0.46]** | **96.0%** | **--** |
| **Random effects model** | | | **0.45** | **[0.27; 0.64]** | **--** | **76.9%** |
| Heterogeneity: $I^2 = 97\%$, $\tau^2 = 0.4594$, $p < 0.01$ | | | | | | |
| **Common effect model** | | **1208** | **0.45** | **[0.42; 0.48]** | **100.0%** | **--** |
| **Random effects model** | | | **0.52** | **[0.33; 0.71]** | **--** | **100.0%** |

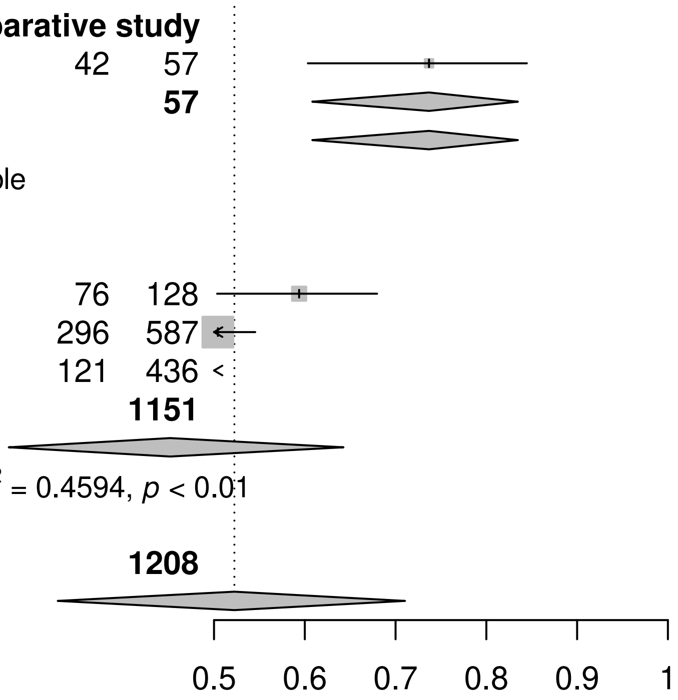

Heterogeneity: $I^2 = 96\%$, $\tau^2 = 0.6454$, $p < 0.01$
Test for subgroup differences (fixed effect): $\chi^2_1 = 17.70$, df = 1 ($p < 0.01$)
Test for subgroup differences (random effects): $\chi^2_1 = 6.01$, df = 1 ($p = 0.01$)

016

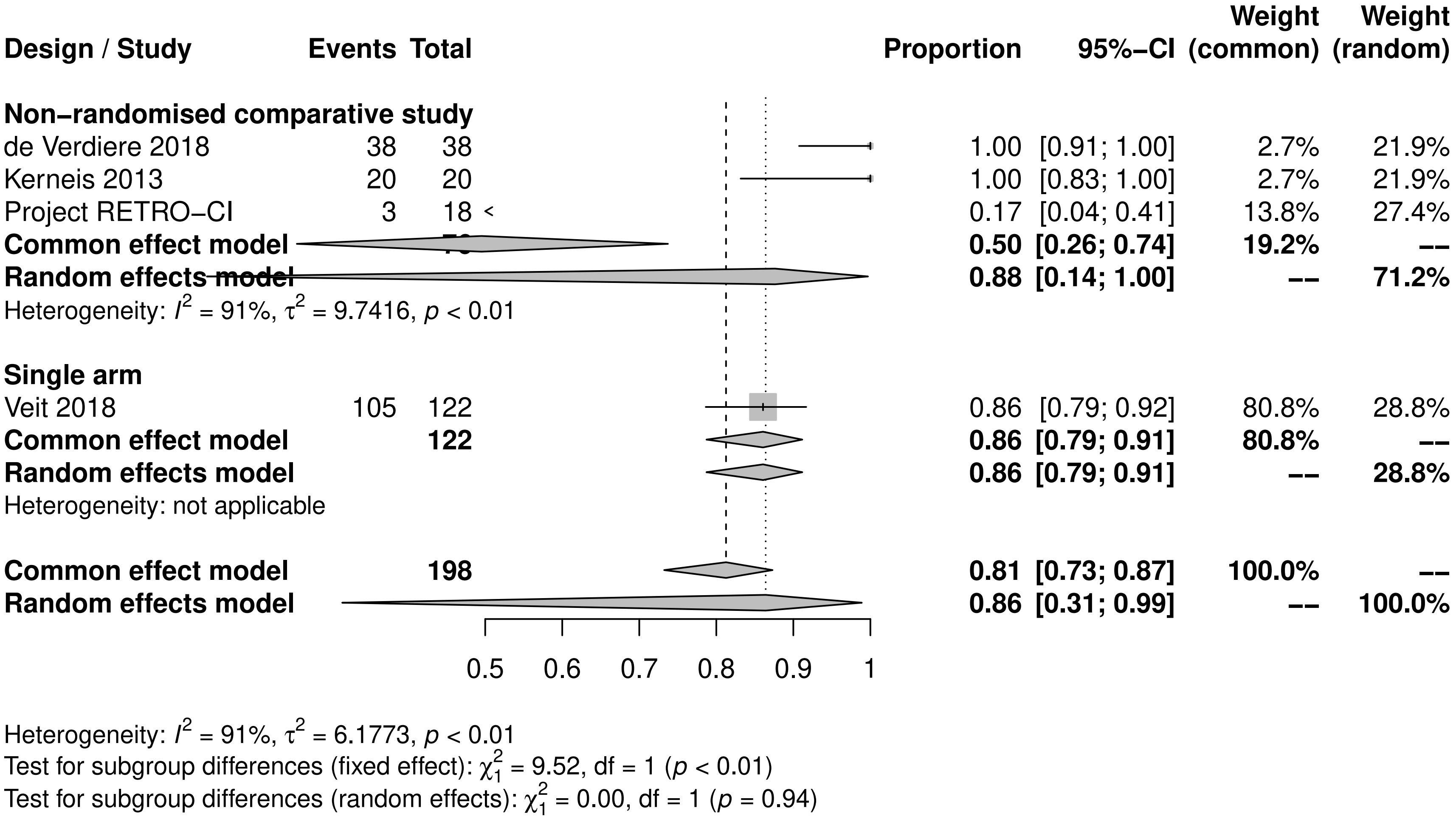

Design / Study
Events
Total
Proportion
95%−CI
Weight (common)
Weight (random)
Non−randomised comparative study
de Verdiere 2018 38 38 1.00 [0.91; 1.00] 2.7% 21.9%
Kerneis 2013 20 20 1.00 [0.83; 1.00] 2.7% 21.9%
Project RETRO−CI 3 18 0.17 [0.04; 0.41] 13.8% 27.4%
Common effect model 0.50 [0.26; 0.74] 19.2% --
Random effects model 0.88 [0.14; 1.00] -- 71.2%
Heterogeneity: $I^2$ = 91%, $\tau^2$ = 9.7416, $p$ < 0.01
Single arm
Veit 2018 105 122 0.86 [0.79; 0.92] 80.8% 28.8%
Common effect model 122 0.86 [0.79; 0.91] 80.8% --
Random effects model 0.86 [0.79; 0.91] -- 28.8%
Heterogeneity: not applicable
Common effect model 198 0.81 [0.73; 0.87] 100.0% --
Random effects model 0.86 [0.31; 0.99] -- 100.0%
0.5 0.6 0.7 0.8 0.9 1
Heterogeneity: $I^2$ = 91%, $\tau^2$ = 6.1773, $p$ < 0.01
Test for subgroup differences (fixed effect): $\chi^2_1$ = 9.52, df = 1 ($p$ < 0.01)
Test for subgroup differences (random effects): $\chi^2_1$ = 0.00, df = 1 ($p$ = 0.94)

017

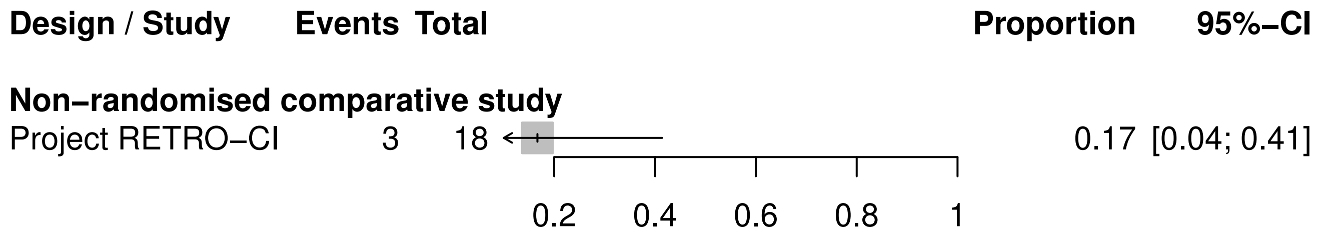
Design / Study
Events
Total
Proportion
95%−CI
Non−randomised comparative study
Project RETRO−CI
3
18
0.17
[0.04; 0.41]
0.2
0.4
0.6
0.8
1

018

| Design / Study | Events | Total | Proportion | 95%-CI | Weight (common) | Weight (random) |
|---|---|---|---|---|---|---|
| **Non-randomised comparative study** | | | | | | |
| de Verdiere 2018 | 38 | 38 | 1.00 | [0.91; 1.00] | 3.2% | 20.4% |
| Kerneis 2013 | 20 | 20 | 1.00 | [0.83; 1.00] | 3.1% | 20.3% |
| **Common effect model** | | **58** | **0.98** | **[0.89; 1.00]** | **6.3%** | **--** |
| **Random effects model** | | | **0.98** | **[0.89; 1.00]** | **--** | **40.7%** |
| Heterogeneity: $I^2 = 0\%$, $\tau^2 = 0$, $p = 0.75$ | | | | | | |
| **Single arm** | | | | | | |
| Veit 2018 | 105 | 122 | 0.86 | [0.79; 0.92] | 93.7% | 59.3% |
| **Common effect model** | | **122** | **0.86** | **[0.79; 0.91]** | **93.7%** | **--** |
| **Random effects model** | | | **0.86** | **[0.79; 0.91]** | **--** | **59.3%** |
| Heterogeneity: not applicable | | | | | | |
| **Common effect model** | | **180** | **0.88** | **[0.81; 0.92]** | **100.0%** | **--** |
| **Random effects model** | | | **0.94** | **[0.77; 0.99]** | **--** | **100.0%** |

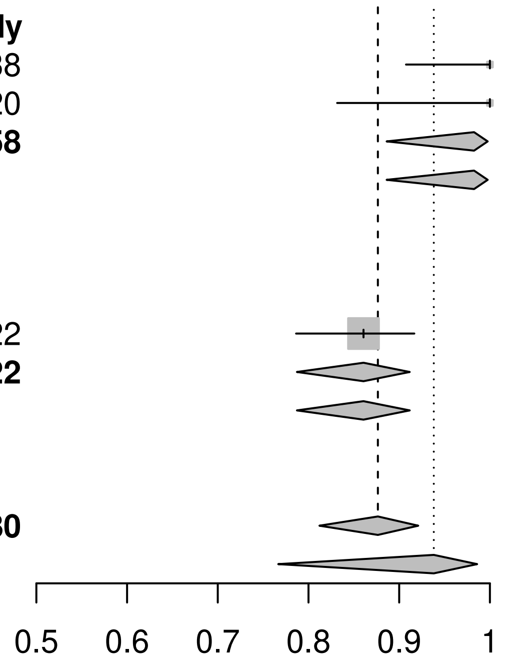

Heterogeneity: $I^2 = 56\%$, $\tau^2 = 0.9579$, $p = 0.10$
Test for subgroup differences (fixed effect): $\chi^2_1 = 4.49$, df = 1 ($p = 0.03$)
Test for subgroup differences (random effects): $\chi^2_1 = 4.49$, df = 1 ($p = 0.03$)

019

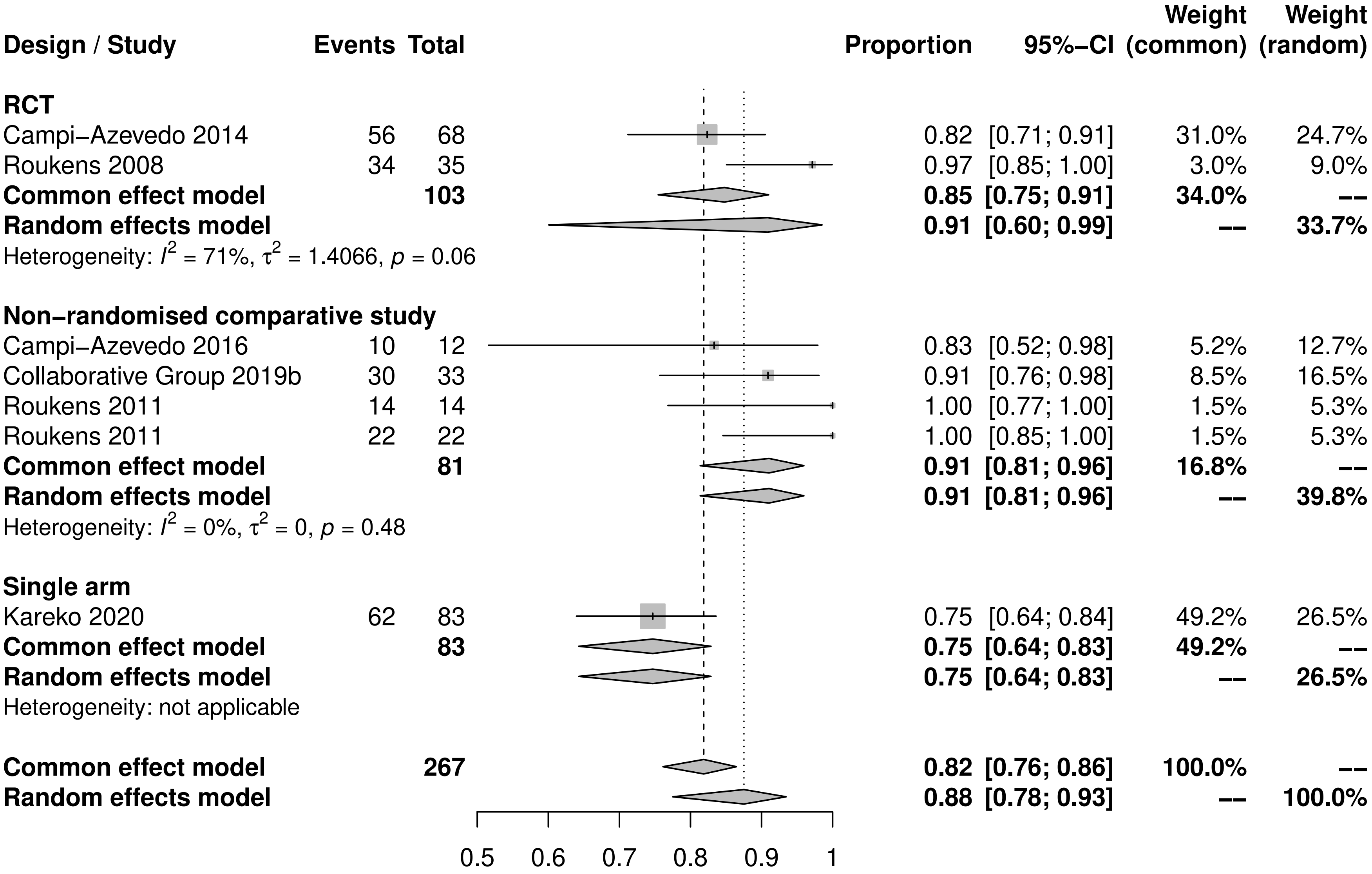

Heterogeneity: $I^2$ = 53%, $\tau^2$ = 0.4313, $p$ = 0.05
Test for subgroup differences (fixed effect): $\chi^2_2$ = 6.86, df = 2 ($p$ = 0.03)
Test for subgroup differences (random effects): $\chi^2_2$ = 6.96, df = 2 ($p$ = 0.03)

020

| Design / Study | Events | Total | Proportion | 95%-CI | Weight (common) | Weight (random) |
|---|---|---|---|---|---|---|
| **Non-randomised comparative study** | | | | | | |
| Campi-Azevedo 2016 | 10 | 12 | 0.83 | [0.52; 0.98] | 37.9% | 37.9% |
| Collaborative Group 2019b | 30 | 33 | 0.91 | [0.76; 0.98] | 62.1% | 62.1% |
| **Common effect model** | | **45** | **0.88** | **[0.75; 0.95]** | **100.0%** | **--** |
| **Random effects model** | | | **0.88** | **[0.75; 0.95]** | **--** | **100.0%** |
| Heterogeneity: $I^2 = 0\%$, $\tau^2 = 0$, $p = 0.48$ | | | | | | |
| **Common effect model** | | **45** | **0.88** | **[0.75; 0.95]** | **100.0%** | **--** |
| **Random effects model** | | | **0.88** | **[0.75; 0.95]** | **--** | **100.0%** |

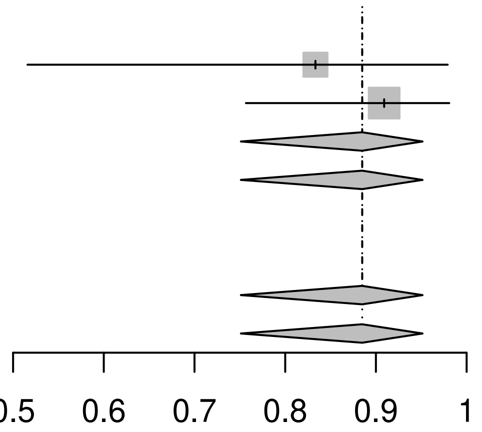

Heterogeneity: $I^2 = 0\%$, $\tau^2 = 0$, $p = 0.48$
Test for subgroup differences (fixed effect): $\chi^2_0 = 0.00$, df = 0 ($p$ = NA)
Test for subgroup differences (random effects): $\chi^2_0 = 0.00$, df = 0 ($p$ = NA)

021

| Design / Study | Events | Total | Proportion | 95%-CI | Weight (common) | Weight (random) |
|---|---|---|---|---|---|---|
| **RCT** | | | | | | |
| Campi−Azevedo 2014 | 56 | 68 | 0.82 | [0.71; 0.91] | 35.9% | 31.0% |
| Roukens 2008 | 34 | 35 | 0.97 | [0.85; 1.00] | 3.5% | 16.0% |
| **Common effect model** | | **103** | **0.85** | **[0.75; 0.91]** | **39.4%** | **--** |
| **Random effects model** | | | **0.91** | **[0.60; 0.99]** | **--** | **47.0%** |
| Heterogeneity: $I^2 = 71\%$, $\tau^2 = 1.4066$, $p = 0.06$ | | | | | | |
| **Non−randomised comparative study** | | | | | | |
| Roukens 2011 | 14 | 14 | 1.00 | [0.77; 1.00] | 1.8% | 10.4% |
| Roukens 2011 | 22 | 22 | 1.00 | [0.85; 1.00] | 1.8% | 10.4% |
| **Common effect model** | | **36** | **0.97** | **[0.83; 1.00]** | **3.5%** | **--** |
| **Random effects model** | | | **0.97** | **[0.83; 1.00]** | **--** | **20.8%** |
| Heterogeneity: $I^2 = 0\%$, $\tau^2 = 0$, $p = 0.83$ | | | | | | |
| **Single arm** | | | | | | |
| Kareko 2020 | 62 | 83 | 0.75 | [0.64; 0.84] | 57.0% | 32.2% |
| **Common effect model** | | **83** | **0.75** | **[0.64; 0.83]** | **57.0%** | **--** |
| **Random effects model** | | | **0.75** | **[0.64; 0.83]** | **--** | **32.2%** |
| Heterogeneity: not applicable | | | | | | |
| **Common effect model** | | **222** | **0.81** | **[0.74; 0.86]** | **100.0%** | **--** |
| **Random effects model** | | | **0.89** | **[0.74; 0.96]** | **--** | **100.0%** |

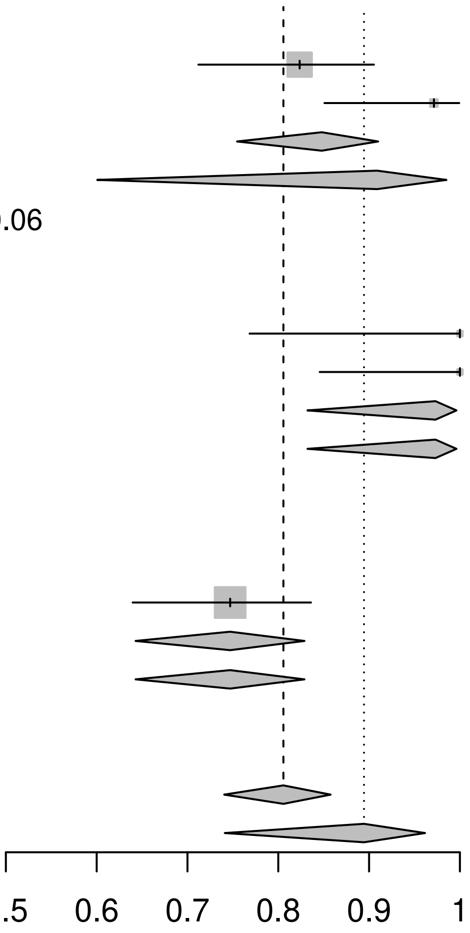

Heterogeneity: $I^2 = 63\%$, $\tau^2 = 0.8869$, $p = 0.03$
Test for subgroup differences (fixed effect): $\chi^2_2 = 7.32$, df = 2 ($p = 0.03$)
Test for subgroup differences (random effects): $\chi^2_2 = 6.91$, df = 2 ($p = 0.03$)

022

| Design / Study | Events | Total | Proportion | 95%-CI | Weight (common) | Weight (random) |
|---|---|---|---|---|---|---|
| **Single arm** | | | | | | |
| Campi–Azevedo 2019 | 61 | 127 | 0.48 | [0.39; 0.57] | 13.3% | 32.1% |
| Domingo 2019 | 188 | 436 | 0.43 | [0.38; 0.48] | 44.7% | 34.0% |
| Idoko 2020 | 338 | 481 | 0.70 | [0.66; 0.74] | 42.0% | 33.9% |
| **Common effect model** | | **1044** | **0.56** | **[0.53; 0.59]** | **100.0%** | **--** |
| **Random effects model** | | | **0.54** | **[0.37; 0.70]** | **--** | **100.0%** |
| Heterogeneity: $I^2 = 97\%$, $\tau^2 = 0.3575$, $p < 0.01$ | | | | | | |
| **Common effect model** | | **1044** | **0.56** | **[0.53; 0.59]** | **100.0%** | **--** |
| **Random effects model** | | | **0.54** | **[0.37; 0.70]** | **--** | **100.0%** |

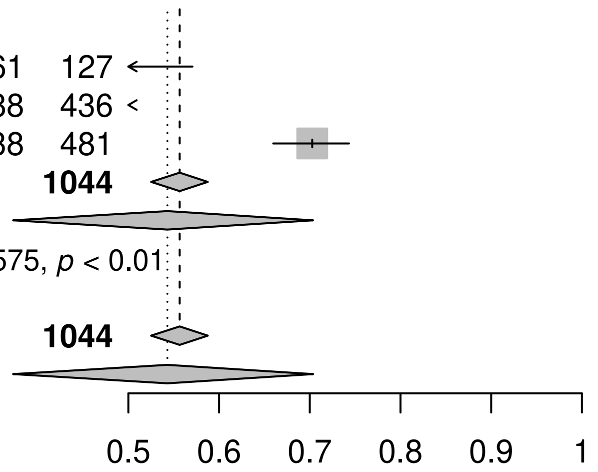

Heterogeneity: $I^2 = 97\%$, $\tau^2 = 0.3575$, $p < 0.01$
Test for subgroup differences (fixed effect): $\chi^2_0 = 0.00$, df = 0 ($p$ = NA)
Test for subgroup differences (random effects): $\chi^2_0 = 0.00$, df = 0 ($p$ = NA)

023

| Design / Study | Events | Total | Proportion | 95%-CI | Weight (common) | Weight (random) |
|---|---|---|---|---|---|---|
| **Single arm** | | | | | | |
| Campi–Azevedo 2019 | 61 | 127 | 0.48 | [0.39; 0.57] | 13.3% | 32.1% |
| Domingo 2019 | 188 | 436 | 0.43 | [0.38; 0.48] | 44.7% | 34.0% |
| Idoko 2020 | 338 | 481 | 0.70 | [0.66; 0.74] | 42.0% | 33.9% |
| **Common effect model** | | **1044** | **0.56** | **[0.53; 0.59]** | **100.0%** | **--** |
| **Random effects model** | | | **0.54** | **[0.37; 0.70]** | **--** | **100.0%** |
| Heterogeneity: $I^2 = 97\%$, $\tau^2 = 0.3575$, $p < 0.01$ | | | | | | |
| **Common effect model** | | **1044** | **0.56** | **[0.53; 0.59]** | **100.0%** | **--** |
| **Random effects model** | | | **0.54** | **[0.37; 0.70]** | **--** | **100.0%** |

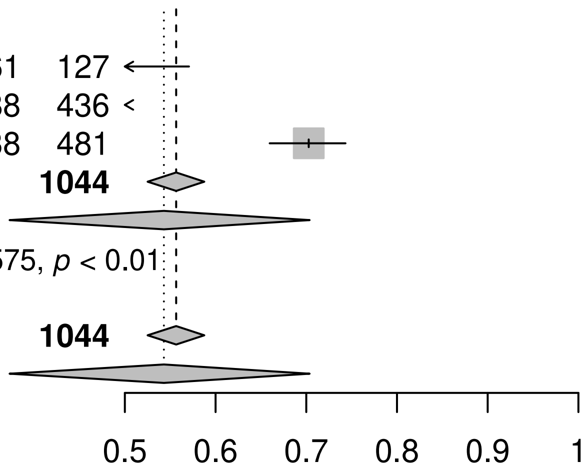

Heterogeneity: $I^2 = 97\%$, $\tau^2 = 0.3575$, $p < 0.01$
Test for subgroup differences (fixed effect): $\chi^2_0 = 0.00$, df = 0 ($p$ = NA)
Test for subgroup differences (random effects): $\chi^2_0 = 0.00$, df = 0 ($p$ = NA)

025

| Design / Study | Events | Total | Proportion | 95%-CI | Weight (common) | Weight (random) |
|---|---|---|---|---|---|---|
| **Non-randomised comparative study** | | | | | | |
| Burkhard 2020 | 4 | 4 | 1.00 | [0.40; 1.00] | 3.6% | 3.6% |
| **Common effect model** | | **4** | **0.90** | **[0.33; 0.99]** | **3.6%** | **--** |
| **Random effects model** | | | **0.90** | **[0.33; 0.99]** | **--** | **3.6%** |
| Heterogeneity: not applicable | | | | | | |
| **Single arm** | | | | | | |
| Veit 2018 | 47 | 63 | 0.75 | [0.62; 0.85] | 96.4% | 96.4% |
| **Common effect model** | | **63** | **0.75** | **[0.62; 0.84]** | **96.4%** | **--** |
| **Random effects model** | | | **0.75** | **[0.62; 0.84]** | **--** | **96.4%** |
| Heterogeneity: not applicable | | | | | | |
| **Common effect model** | | **67** | **0.75** | **[0.64; 0.84]** | **100.0%** | **--** |
| **Random effects model** | | | **0.75** | **[0.64; 0.84]** | **--** | **100.0%** |

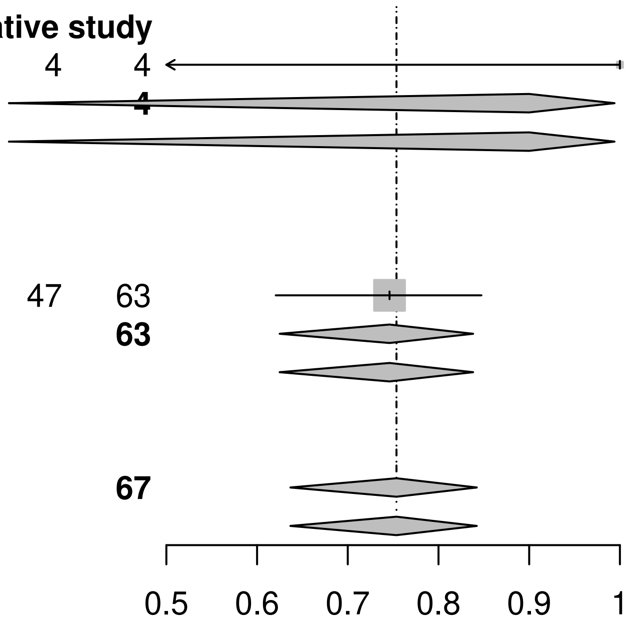

Heterogeneity: $I^2 = 0\%$, $\tau^2 = 0$, $p = 0.46$
Test for subgroup differences (fixed effect): $\chi^2_1 = 0.54$, df = 1 ($p = 0.46$)
Test for subgroup differences (random effects): $\chi^2_1 = 0.54$, df = 1 ($p = 0.46$)

027

| Design / Study | Events | Total | Proportion | 95%-CI | Weight (common) | Weight (random) |
|---|---|---|---|---|---|---|
| **Non-randomised comparative study** | | | | | | |
| Burkhard 2020 | 4 | 4 | 1.00 | [0.40; 1.00] | 3.6% | 3.6% |
| **Common effect model** | | **4** | **0.90** | **[0.33; 0.99]** | **3.6%** | **--** |
| **Random effects model** | | | **0.90** | **[0.33; 0.99]** | **--** | **3.6%** |
| Heterogeneity: not applicable | | | | | | |
| **Single arm** | | | | | | |
| Veit 2018 | 47 | 63 | 0.75 | [0.62; 0.85] | 96.4% | 96.4% |
| **Common effect model** | | **63** | **0.75** | **[0.62; 0.84]** | **96.4%** | **--** |
| **Random effects model** | | | **0.75** | **[0.62; 0.84]** | **--** | **96.4%** |
| Heterogeneity: not applicable | | | | | | |
| **Common effect model** | | **67** | **0.75** | **[0.64; 0.84]** | **100.0%** | **--** |
| **Random effects model** | | | **0.75** | **[0.64; 0.84]** | **--** | **100.0%** |

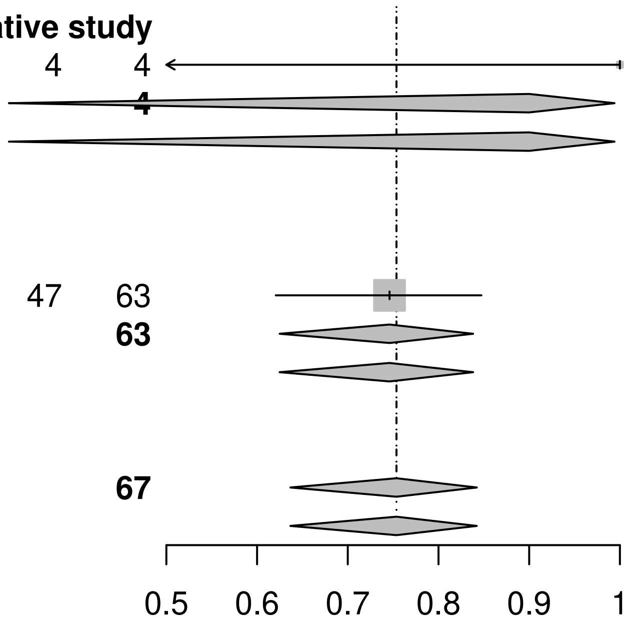

Heterogeneity: $I^2 = 0\%$, $\tau^2 = 0$, $p = 0.46$
Test for subgroup differences (fixed effect): $\chi^2_1 = 0.54$, df = 1 ($p = 0.46$)
Test for subgroup differences (random effects): $\chi^2_1 = 0.54$, df = 1 ($p = 0.46$)

028

| Design / Study | Events | Total | Proportion | 95%−CI | Weight (common) | Weight (random) |
|---|---|---|---|---|---|---|
| **Non−randomised comparative study** | | | | | | |
| Campi−Azevedo 2016 | 33 | 39 | 0.85 | [0.69; 0.94] | 10.9% | 16.3% |
| Collaborative Group 2019a | 68 | 99 | 0.69 | [0.59; 0.78] | 45.7% | 34.3% |
| Collaborative Group 2019b | 39 | 55 | 0.71 | [0.57; 0.82] | 24.4% | 26.3% |
| **Common effect model** | | **193** | **0.72** | **[0.65; 0.78]** | **81.0%** | **−−** |
| **Random effects model** | | | **0.74** | **[0.63; 0.82]** | **−−** | **76.9%** |
| Heterogeneity: $I^2$ = 43%, $\tau^2$ = 0.1051, $p$ = 0.17 | | | | | | |
| **Single arm** | | | | | | |
| Kareko 2020 | 24 | 38 | 0.63 | [0.46; 0.78] | 19.0% | 23.1% |
| **Common effect model** | | **38** | **0.63** | **[0.47; 0.77]** | **19.0%** | **−−** |
| **Random effects model** | | | **0.63** | **[0.47; 0.77]** | **−−** | **23.1%** |
| Heterogeneity: not applicable | | | | | | |
| **Common effect model** | | **231** | **0.70** | **[0.64; 0.76]** | **100.0%** | **−−** |
| **Random effects model** | | | **0.71** | **[0.62; 0.79]** | **−−** | **100.0%** |

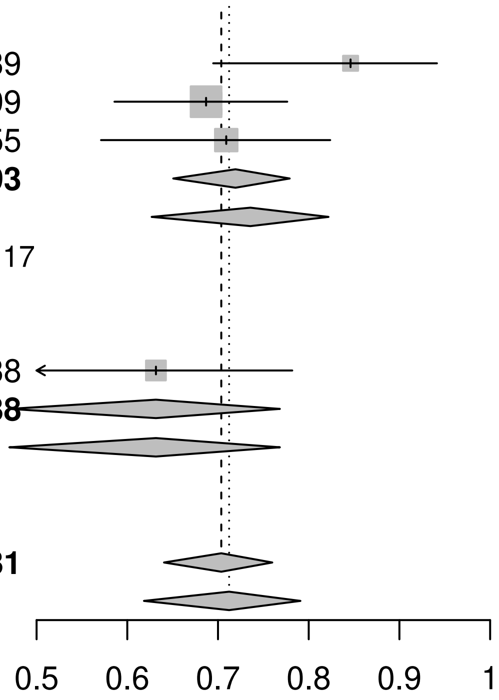

Heterogeneity: $I^2$ = 36%, $\tau^2$ = 0.0895, $p$ = 0.20
Test for subgroup differences (fixed effect): $\chi^2_1$ = 1.16, df = 1 ($p$ = 0.28)
Test for subgroup differences (random effects): $\chi^2_1$ = 1.31, df = 1 ($p$ = 0.25)

029

| Design / Study | Events | Total | Proportion | 95%-CI | Weight (common) | Weight (random) |
|---|---|---|---|---|---|---|
| **Non-randomised comparative study** | | | | | | |
| Campi-Azevedo 2016 | 33 | 39 | 0.85 | [0.69; 0.94] | 13.5% | 22.0% |
| Collaborative Group 2019a | 68 | 99 | 0.69 | [0.59; 0.78] | 56.5% | 43.7% |
| Collaborative Group 2019b | 39 | 55 | 0.71 | [0.57; 0.82] | 30.1% | 34.4% |
| **Common effect model** | | **193** | **0.72** | **[0.65; 0.78]** | **100.0%** | **--** |
| **Random effects model** | | | **0.74** | **[0.63; 0.82]** | **--** | **100.0%** |
| Heterogeneity: $I^2 = 43\%$, $\tau^2 = 0.1051$, $p = 0.17$ | | | | | | |
| **Common effect model** | | **193** | **0.72** | **[0.65; 0.78]** | **100.0%** | **--** |
| **Random effects model** | | | **0.74** | **[0.63; 0.82]** | **--** | **100.0%** |

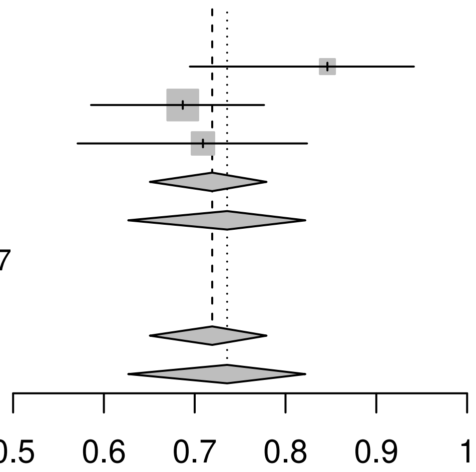

Heterogeneity: $I^2 = 43\%$, $\tau^2 = 0.1051$, $p = 0.17$
Test for subgroup differences (fixed effect): $\chi^2_0 = 0.00$, df = 0 ($p$ = NA)
Test for subgroup differences (random effects): $\chi^2_0 = 0.00$, df = 0 ($p$ = NA)

030

| Design / Study | Events | Total | Proportion | 95%−CI |
|---|---|---|---|---|
| **Single arm** | | | | |
| Kareko 2020 | 24 | 38 | 0.63 | [0.46; 0.78] |

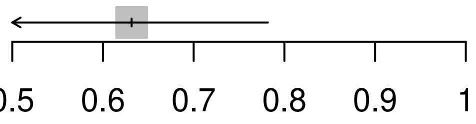

034

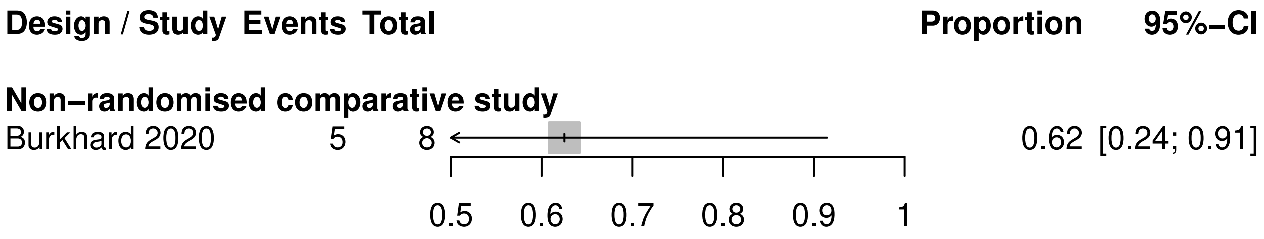

Design / Study
Events
Total
Proportion
95%−CI
Non−randomised comparative study
Burkhard 2020
5
8
0.62
[0.24; 0.91]
0.5
0.6
0.7
0.8
0.9
1

036

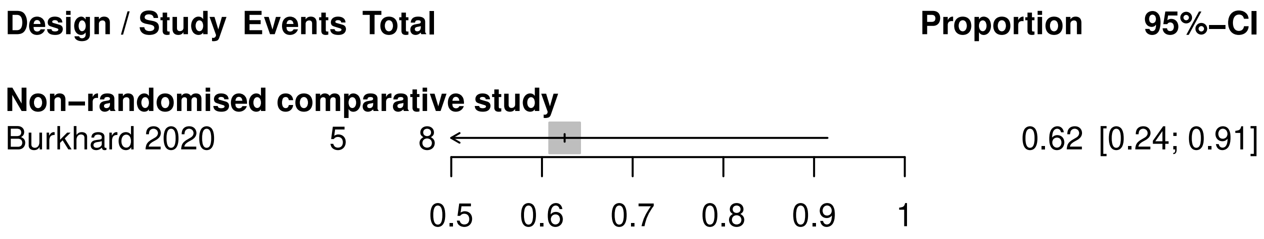
Design / Study
Events
Total
Proportion
95%−CI
Non−randomised comparative study
Burkhard 2020
5
8
0.62
[0.24; 0.91]
0.5
0.6
0.7
0.8
0.9
1

037

| Design / Study | Events | Total | Proportion | 95%−CI |
|---|---|---|---|---|
| **Non−randomised comparative study** | | | | |
| Burkhard 2020 | 0 | 1 | 0.00 | [ 0; 0.98] |

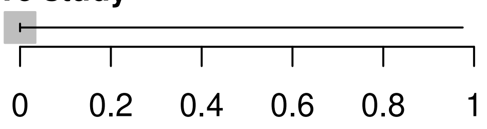

039

| Design / Study | Events | Total | Proportion | 95%-CI |
|---|---|---|---|---|
| **Non-randomised comparative study** | | | | |
| Burkhard 2020 | 0 | 1 | 0.00 | [ 0; 0.98] |

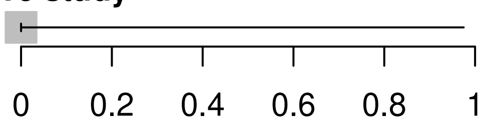

043

| Design / Study | Events | Total | Proportion | 95%−CI |
|---|---|---|---|---|
| **Non−randomised comparative study** | | | | |
| Burkhard 2020 | 16 | 17 | 0.94 | [0.71; 1] |

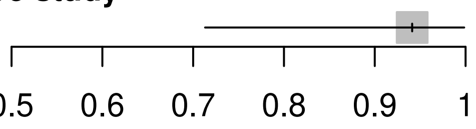

045

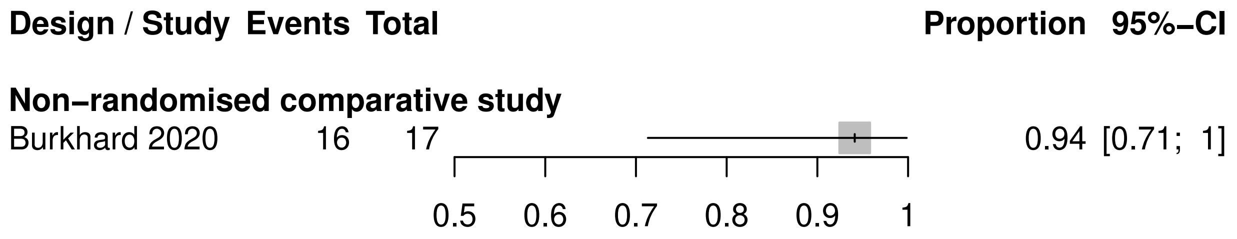

Design / Study
Events
Total
Proportion
95%−CI
Non−randomised comparative study
Burkhard 2020
16
17
0.94
[0.71; 1]
0.5
0.6
0.7
0.8
0.9
1

046

| Design / Study | Events | Total | Proportion | 95%-CI | Weight (common) | Weight (random) |
|---|---|---|---|---|---|---|
| **RCT** | | | | | | |
| Roukens 2008 | 19 | 19 | 1.00 | [0.82; 1] | 49.6% | 49.6% |
| **Common effect model** | | **19** | **0.98** | **[0.70; 1]** | **49.6%** | **--** |
| **Random effects model** | | | **0.98** | **[0.70; 1]** | **--** | **49.6%** |
| Heterogeneity: not applicable | | | | | | |
| **Non-randomised comparative study** | | | | | | |
| Collaborative Group 2019b | 45 | 45 | 1.00 | [0.92; 1] | 50.4% | 50.4% |
| **Common effect model** | | **45** | **0.99** | **[0.85; 1]** | **50.4%** | **--** |
| **Random effects model** | | | **0.99** | **[0.85; 1]** | **--** | **50.4%** |
| Heterogeneity: not applicable | | | | | | |
| **Common effect model** | | **64** | **0.98** | **[0.89; 1]** | **100.0%** | **--** |
| **Random effects model** | | | **0.98** | **[0.89; 1]** | **--** | **100.0%** |

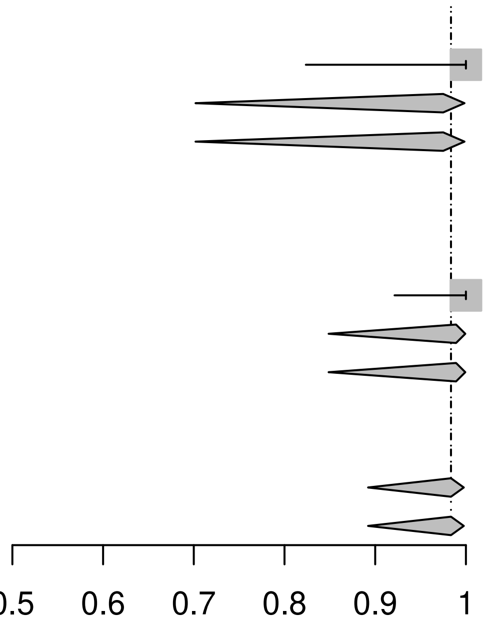

Heterogeneity: $I^2 = 0\%$, $\tau^2 = 0$, $p = 0.67$
Test for subgroup differences (fixed effect): $\chi^2_1 = 0.18$, df = 1 ($p = 0.67$)
Test for subgroup differences (random effects): $\chi^2_1 = 0.18$, df = 1 ($p = 0.67$)

047

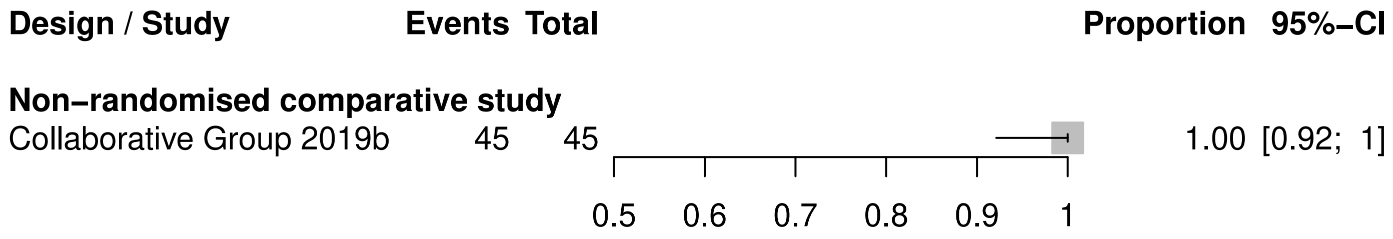
Design / Study
Events
Total
Proportion
95%−CI
Non−randomised comparative study
Collaborative Group 2019b
45
45
1.00 [0.92; 1]
0.5
0.6
0.7
0.8
0.9
1

048

| Design / Study | Events | Total | Proportion | 95%-CI |
|---|---|---|---|---|
| **RCT** | | | | |
| Roukens 2008 | 19 | 19 | 1.00 | [0.82; 1] |

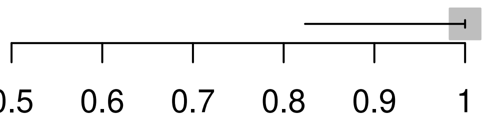

052

| Design / Study | Events | Total | Proportion | 95%−CI |
|---|---|---|---|---|
| **Non−randomised comparative study** | | | | |
| Kerneis 2013 | 11 | 11 | 1.00 | [0.72; 1] |

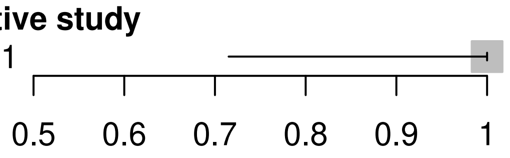

054

| Design / Study | Events | Total | | Proportion | 95%−CI |
|---|---|---|---|---|---|
| **Non−randomised comparative study** | | | | | |
| Kerneis 2013 | 11 | 11 | 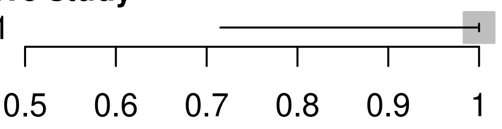 | 1.00 | [0.72; 1] |

055

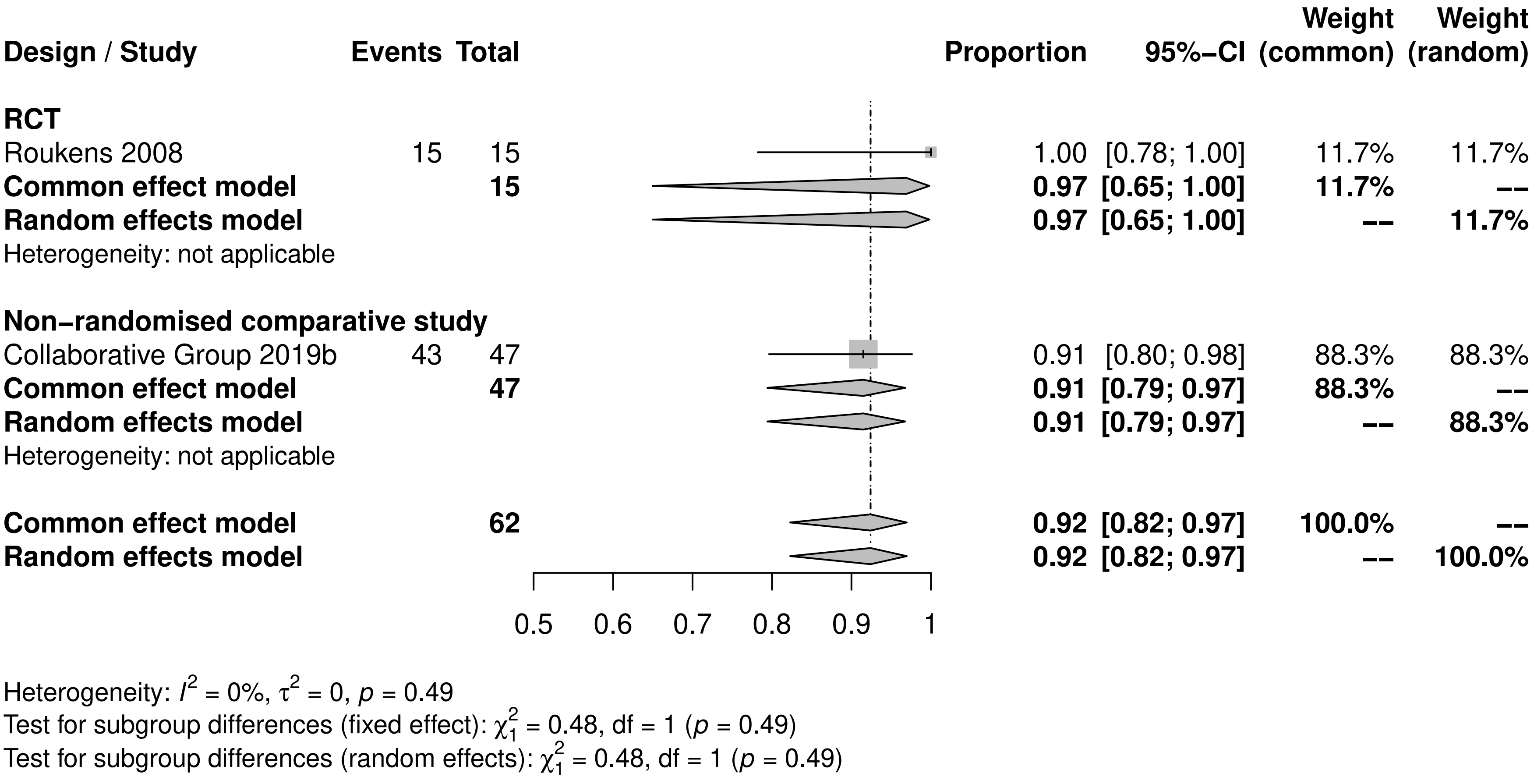
Design / Study
Events
Total
Proportion
95%−CI
Weight (common)
Weight (random)
RCT
Roukens 2008 15 15 1.00 [0.78; 1.00] 11.7% 11.7%
Common effect model 15 0.97 [0.65; 1.00] 11.7% --
Random effects model 0.97 [0.65; 1.00] -- 11.7%
Heterogeneity: not applicable
Non−randomised comparative study
Collaborative Group 2019b 43 47 0.91 [0.80; 0.98] 88.3% 88.3%
Common effect model 47 0.91 [0.79; 0.97] 88.3% --
Random effects model 0.91 [0.79; 0.97] -- 88.3%
Heterogeneity: not applicable
Common effect model 62 0.92 [0.82; 0.97] 100.0% --
Random effects model 0.92 [0.82; 0.97] -- 100.0%
0.5
0.6
0.7
0.8
0.9
1
Heterogeneity: $I^2$ = 0%, $\tau^2$ = 0, $p$ = 0.49
Test for subgroup differences (fixed effect): $\chi^2_1$ = 0.48, df = 1 ($p$ = 0.49)
Test for subgroup differences (random effects): $\chi^2_1$ = 0.48, df = 1 ($p$ = 0.49)

056

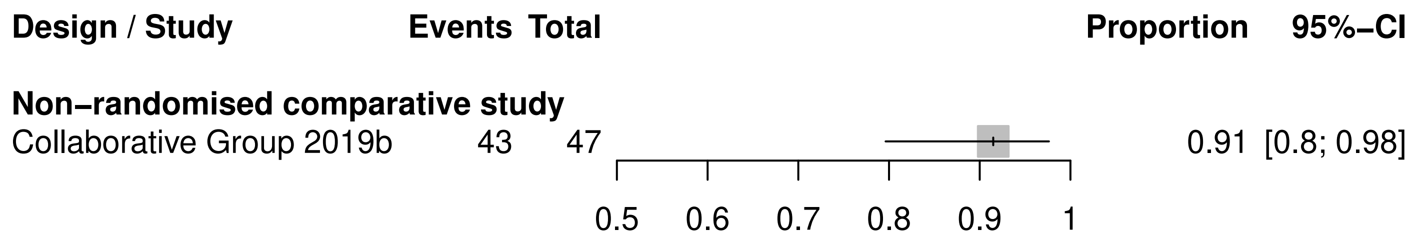

Design / Study
Events
Total
Proportion
95%−CI
Non−randomised comparative study
Collaborative Group 2019b
43
47
0.91
[0.8; 0.98]
0.5
0.6
0.7
0.8
0.9
1

057

| Design / Study | Events | Total | Proportion | 95%−CI |
|---|---|---|---|---|
| **RCT** | | | | |
| Roukens 2008 | 15 | 15 | 1.00 | [0.78; 1] |

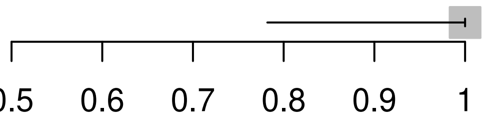

064

| Design / Study | Events | Total | Proportion | 95%−CI | Weight (common) | Weight (random) |
|---|---|---|---|---|---|---|
| **Non−randomised comparative study** | | | | | | |
| Collaborative Group 2019a | 189 | 215 | 0.88 | [0.83; 0.92] | 85.1% | 85.1% |
| Collaborative Group 2019b | 30 | 34 | 0.88 | [0.73; 0.97] | 13.1% | 13.1% |
| **Common effect model** | | **249** | **0.88** | **[0.83; 0.91]** | **98.2%** | **--** |
| **Random effects model** | | | **0.88** | **[0.83; 0.91]** | **--** | **98.2%** |
| Heterogeneity: $I^2 = 0\%$, $\tau^2 = 0$, $p = 0.96$ | | | | | | |
| **Single arm** | | | | | | |
| Kareko 2020 | 9 | 9 | 1.00 | [0.66; 1.00] | 1.8% | 1.8% |
| **Common effect model** | | **9** | **0.95** | **[0.53; 1.00]** | **1.8%** | **--** |
| **Random effects model** | | | **0.95** | **[0.53; 1.00]** | **--** | **1.8%** |
| Heterogeneity: not applicable | | | | | | |
| **Common effect model** | | **258** | **0.88** | **[0.84; 0.92]** | **100.0%** | **--** |
| **Random effects model** | | | **0.88** | **[0.84; 0.92]** | **--** | **100.0%** |

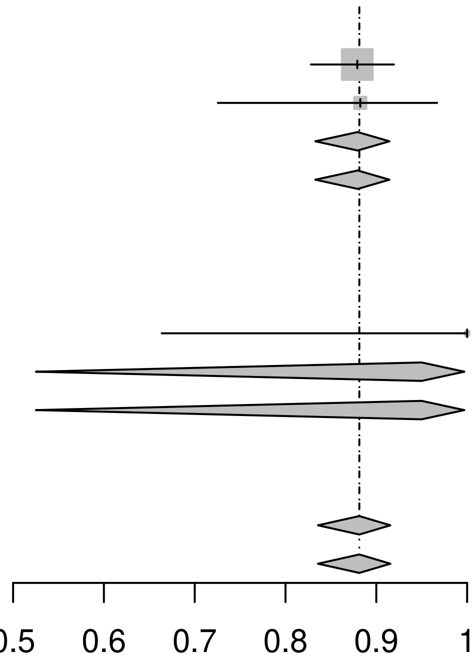

Heterogeneity: $I^2 = 0\%$, $\tau^2 = 0$, $p = 0.81$
Test for subgroup differences (fixed effect): $\chi^2_1 = 0.43$, df = 1 ($p = 0.51$)
Test for subgroup differences (random effects): $\chi^2_1 = 0.43$, df = 1 ($p = 0.51$)

065

| Design / Study | Events | Total | Proportion | 95%−CI | Weight (common) | Weight (random) |
|---|---|---|---|---|---|---|
| **Non−randomised comparative study** | | | | | | |
| Collaborative Group 2019a | 189 | 215 | 0.88 | [0.83; 0.92] | 86.6% | 86.6% |
| Collaborative Group 2019b | 30 | 34 | 0.88 | [0.73; 0.97] | 13.4% | 13.4% |
| **Common effect model** | | **249** | **0.88** | **[0.83; 0.91]** | **100.0%** | **--** |
| **Random effects model** | | | **0.88** | **[0.83; 0.91]** | **--** | **100.0%** |
| Heterogeneity: $I^2 = 0\%$, $\tau^2 = 0$, $p = 0.96$ | | | | | | |
| **Common effect model** | | **249** | **0.88** | **[0.83; 0.91]** | **100.0%** | **--** |
| **Random effects model** | | | **0.88** | **[0.83; 0.91]** | **--** | **100.0%** |

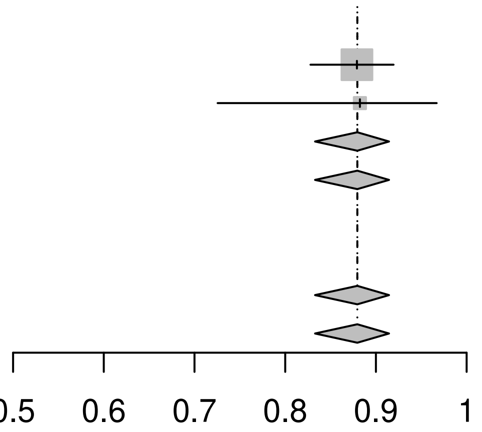

Heterogeneity: $I^2 = 0\%$, $\tau^2 = 0$, $p = 0.96$
Test for subgroup differences (fixed effect): $\chi^2_0 = 0.00$, df = 0 ($p$ = NA)
Test for subgroup differences (random effects): $\chi^2_0 = 0.00$, df = 0 ($p$ = NA)

066

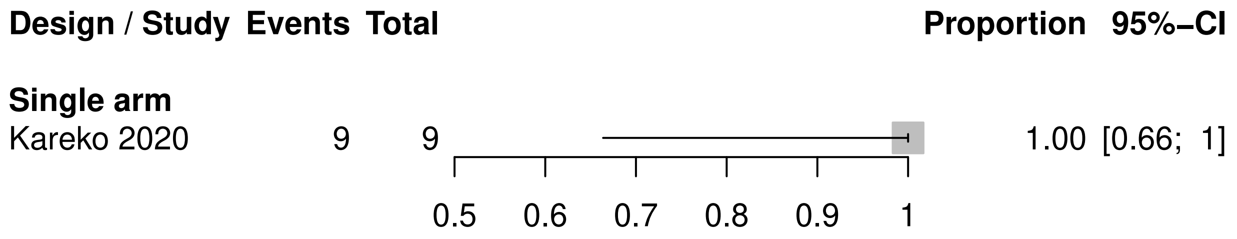
Design / Study Events Total
Proportion 95%−CI
Single arm
Kareko 2020 9 9
1.00 [0.66; 1]
0.5 0.6 0.7 0.8 0.9 1

073

| Design / Study | Events | Total | Proportion | 95%−CI | Weight (common) | Weight (random) |
|---|---|---|---|---|---|---|
| **Non−randomised comparative study** | | | | | | |
| Collaborative Group 2019b | 10 | 12 | 0.83 | [0.52; 0.98] | 78.4% | 78.4% |
| **Common effect model** | | **12** | **0.83** | **[0.52; 0.96]** | **78.4%** | **--** |
| **Random effects model** | | | **0.83** | **[0.52; 0.96]** | **--** | **78.4%** |
| Heterogeneity: not applicable | | | | | | |
| **Single arm** | | | | | | |
| Kareko 2020 | 5 | 5 | 1.00 | [0.48; 1.00] | 21.6% | 21.6% |
| **Common effect model** | | **5** | **0.92** | **[0.38; 0.99]** | **21.6%** | **--** |
| **Random effects model** | | | **0.92** | **[0.38; 0.99]** | **--** | **21.6%** |
| Heterogeneity: not applicable | | | | | | |
| **Common effect model** | | **17** | **0.86** | **[0.61; 0.96]** | **100.0%** | **--** |
| **Random effects model** | | | **0.86** | **[0.61; 0.96]** | **--** | **100.0%** |

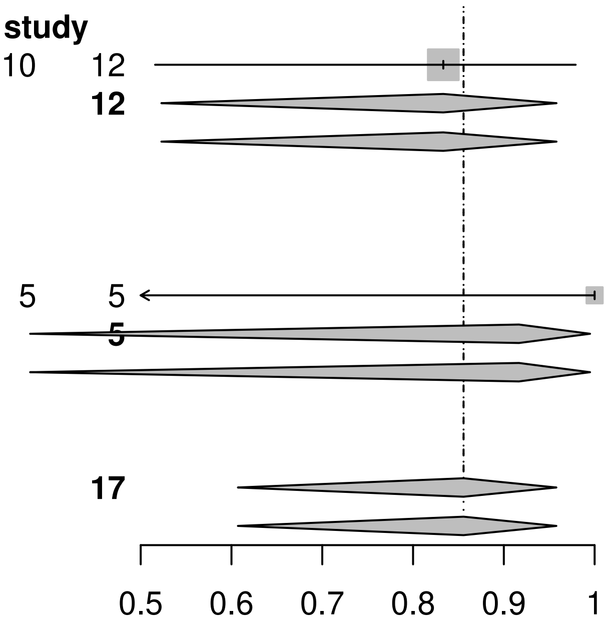

Heterogeneity: $I^2 = 0\%$, $\tau^2 = 0$, $p = 0.64$
Test for subgroup differences (fixed effect): $\chi^2_1 = 0.22$, df = 1 ($p = 0.64$)
Test for subgroup differences (random effects): $\chi^2_1 = 0.22$, df = 1 ($p = 0.64$)

074

| Design / Study | Events | Total | Proportion | 95%-CI |
|---|---|---|---|---|
| **Non-randomised comparative study** | | | | |
| Collaborative Group 2019b | 10 | 12 | 0.83 | [0.52; 0.98] |

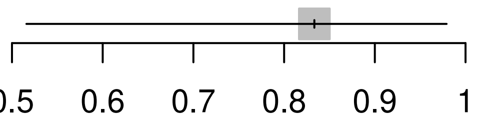

075

| Design / Study | Events | Total | | Proportion | 95%-CI |
|---|---|---|---|---|---|
| **Single arm** | | | | | |
| Kareko 2020 | 5 | 5 | 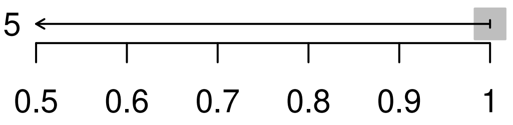 | 1.00 | [0.48; 1] |

088

| Design / Study | Events | Total | Proportion | 95%-CI |
|---|---|---|---|---|
| **Non-randomised comparative study** | | | | |
| Burkhard 2020 | 35 | 40 | 0.88 | [0.73; 0.96] |

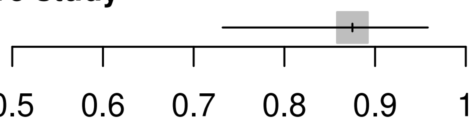

090

| Design / Study | Events | Total | Proportion | 95%-CI |
|---|---|---|---|---|
| **Non-randomised comparative study** | | | | |
| Burkhard 2020 | 35 | 40 | 0.88 | [0.73; 0.96] |

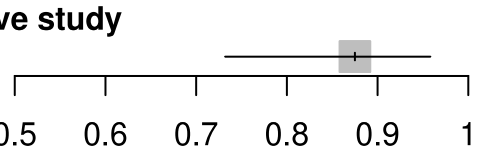

100

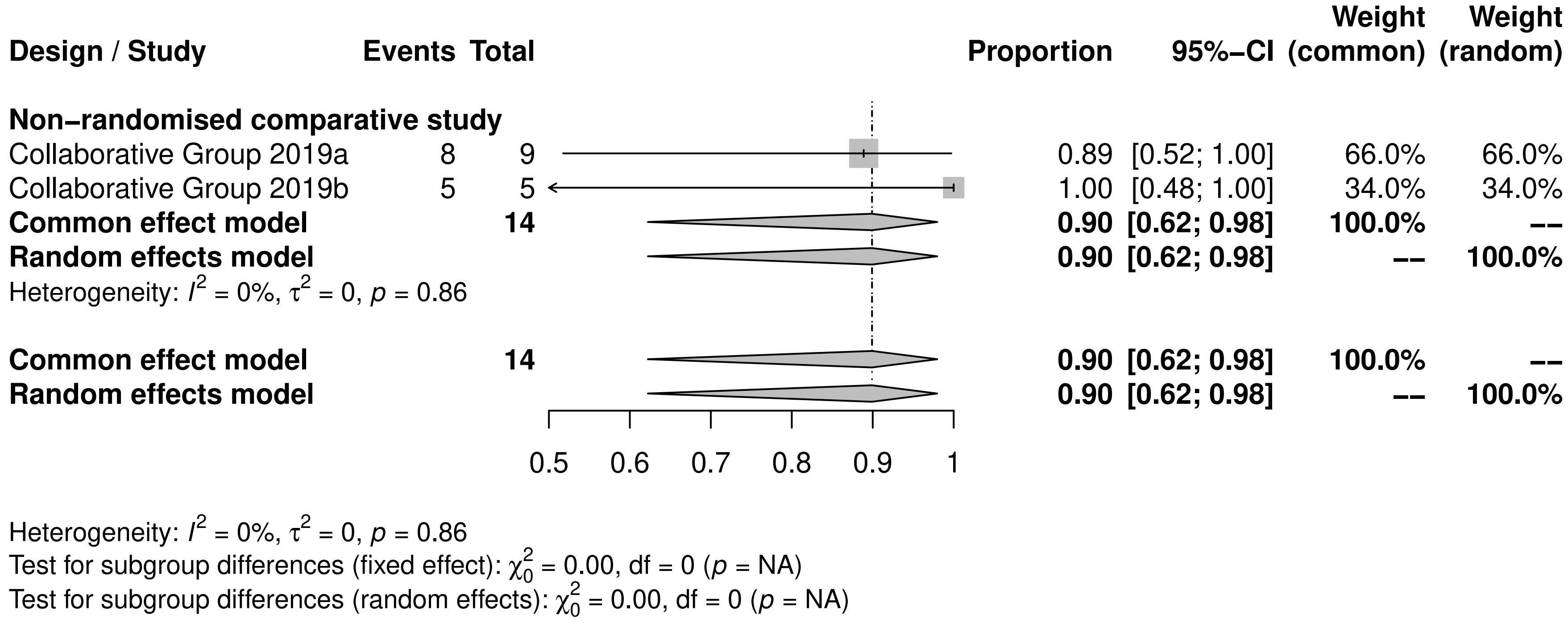
Design / Study
Events
Total
Proportion
95%-CI
Weight (common)
Weight (random)
Non-randomised comparative study
Collaborative Group 2019a 8 9 0.89 [0.52; 1.00] 66.0% 66.0%
Collaborative Group 2019b 5 5 1.00 [0.48; 1.00] 34.0% 34.0%
Common effect model 14 0.90 [0.62; 0.98] 100.0% --
Random effects model 0.90 [0.62; 0.98] -- 100.0%
Heterogeneity: $I^2$ = 0%, $\tau^2$ = 0, $p$ = 0.86
Common effect model 14 0.90 [0.62; 0.98] 100.0% --
Random effects model 0.90 [0.62; 0.98] -- 100.0%
0.5
0.6
0.7
0.8
0.9
1
Heterogeneity: $I^2$ = 0%, $\tau^2$ = 0, $p$ = 0.86
Test for subgroup differences (fixed effect): $\chi^2_0$ = 0.00, df = 0 ($p$ = NA)
Test for subgroup differences (random effects): $\chi^2_0$ = 0.00, df = 0 ($p$ = NA)

101

| Design / Study | Events | Total | Proportion | 95%-CI | Weight (common) | Weight (random) |
|---|---|---|---|---|---|---|
| **Non-randomised comparative study** | | | | | | |
| Collaborative Group 2019a | 8 | 9 | 0.89 | [0.52; 1.00] | 66.0% | 66.0% |
| Collaborative Group 2019b | 5 | 5 | 1.00 | [0.48; 1.00] | 34.0% | 34.0% |
| **Common effect model** | | **14** | **0.90** | **[0.62; 0.98]** | **100.0%** | **--** |
| **Random effects model** | | | **0.90** | **[0.62; 0.98]** | **--** | **100.0%** |
| Heterogeneity: $I^2 = 0\%$, $\tau^2 = 0$, $p = 0.86$ | | | | | | |
| **Common effect model** | | **14** | **0.90** | **[0.62; 0.98]** | **100.0%** | **--** |
| **Random effects model** | | | **0.90** | **[0.62; 0.98]** | **--** | **100.0%** |

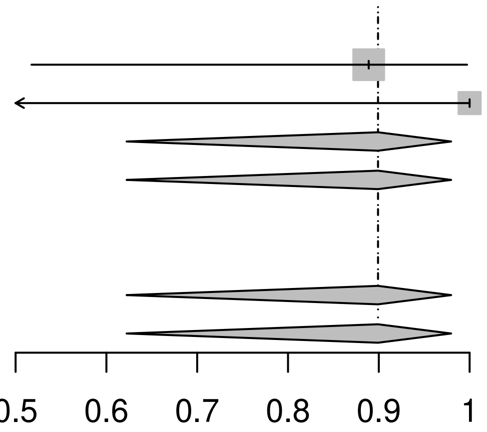

Heterogeneity: $I^2 = 0\%$, $\tau^2 = 0$, $p = 0.86$
Test for subgroup differences (fixed effect): $\chi^2_0 = 0.00$, df = 0 ($p$ = NA)
Test for subgroup differences (random effects): $\chi^2_0 = 0.00$, df = 0 ($p$ = NA)

124

| Design / Study | Events | Total | | Proportion | 95%−CI |
|---|---|---|---|---|---|
| **Non−randomised comparative study** | | | | | |
| Burkhard 2020 | 2 | 2 | 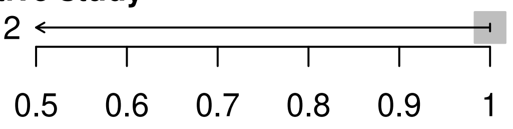 | 1.00 | [0.16; 1] |

126

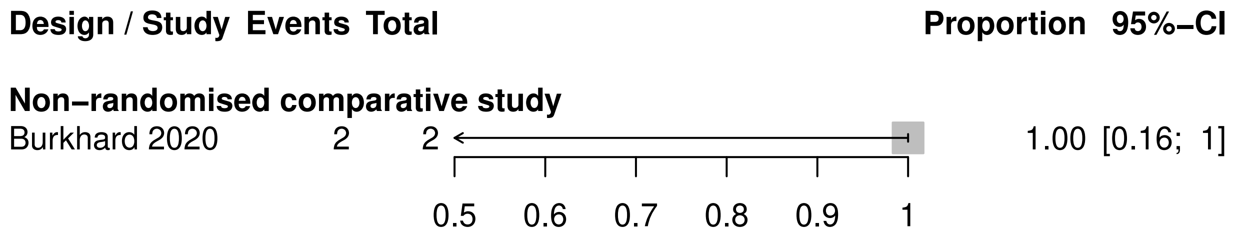

Design / Study Events Total
Proportion 95%−CI
Non−randomised comparative study
Burkhard 2020 2 2
1.00 [0.16; 1]
0.5 0.6 0.7 0.8 0.9 1

133

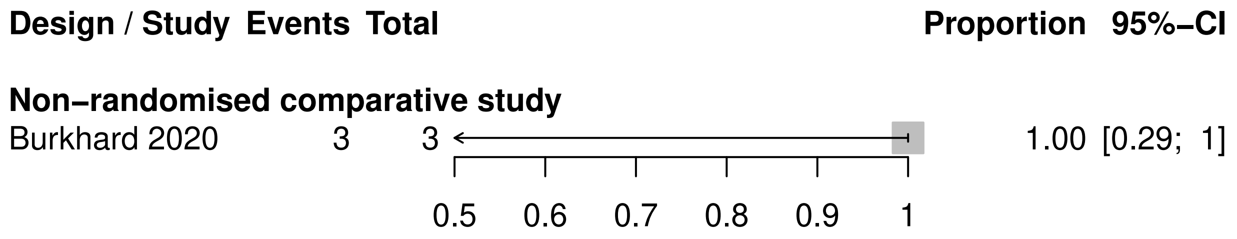

Design / Study
Events
Total
Proportion
95%−CI
Non−randomised comparative study
Burkhard 2020
3
3
1.00 [0.29; 1]
0.5
0.6
0.7
0.8
0.9
1

135

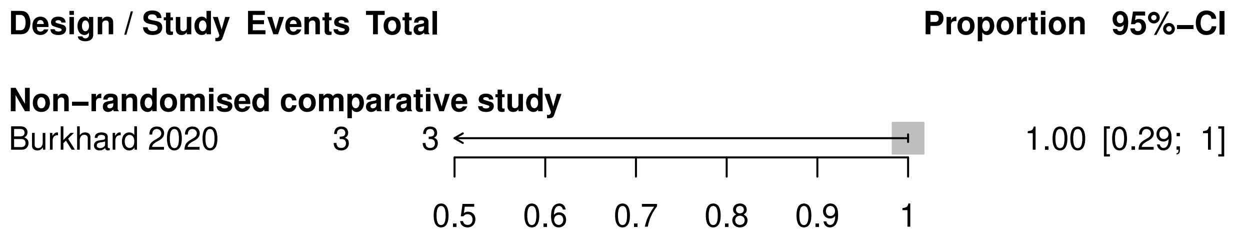

Design / Study
Events
Total
Proportion
95%−CI
Non−randomised comparative study
Burkhard 2020
3
3
1.00 [0.29; 1]
0.5
0.6
0.7
0.8
0.9
1
